# Health-economic impacts of age- and sex-targeted Lassa fever vaccination in endemic regions of Nigeria, Guinea, Liberia and Sierra Leone: a modelling study

**DOI:** 10.1101/2025.03.31.25324924

**Authors:** David R M Smith, Mary Chriselda Antony Oliver, Katherine M Holohan, Harry R Street, Andrew A Torkelson, Danny Asogun, Oladele Oluwafemi Ayodeji, Benedict N Azuogu, Anton Camacho, William A Fischer, Donald S Grant, Kamji Jan, Sylvanus A Okogbenin, Jefferson Sibley, David A Wohl, T Déirdre Hollingsworth, Koen B Pouwels

**Author notes:** contributed equally (T Déirdre Hollingsworth, Koen B Pouwels).

## Abstract

**Introduction:** Lassa fever is an emerging zoonotic disease endemic to West Africa. Several vaccines aimed at preventing Lassa fever are currently under development, creating a need to assess how best to administer them once licensed for human use.

**Methods:** We developed a mathematical model using a zoonosis risk map and epidemiological data from recent and ongoing cohort studies to predict the health-economic burden of Lassa fever across age and sex groups in endemic regions. We simulated vaccination campaigns targeting different risk groups to estimate the health-economic benefits of various scenarios for Lassa vaccine administration. Threshold vaccine costs (TVCs) were estimated in International dollars (I$ 2023), accounting for healthcare costs, productivity losses and monetised disability-adjusted life-years (DALYs) averted due to vaccination.

**Results:** The most efficient strategy for preventing symptomatic Lassa fever was targeting children aged 2-14, for preventing hospitalisation was targeting adolescents and adults aged 15-49, for preventing death was targeting older adults aged 50+, and for preventing DALYs was targeting women of childbearing age (WCBA) aged 15-49. Under base case assumptions, the highest TVC for a single-dose vaccine was estimated at I$7.39 (95% uncertainty interval (UI): I$4.33-I$11.60) when targeting adolescents and adults, followed by I$6.69 (95%UI: I$4.17-I$9.85) when targeting older adults, I$6.10 (95%UI: I$3.56-I$9.74) when targeting WCBA and I$1.94 (95%UI: I$1.10-I$3.10) when targeting children.

**Conclusion:** These results suggest that the most cost-efficient use of limited doses of Lassa fever vaccine is to target adolescents and adults followed by the elderly.

## INTRODUCTION

Lassa fever (LF) is an emerging viral haemorrhagic disease caused by *Lassa mammarenavirus* (LASV). LASV is endemic to West Africa and phylogenetic data support that most human infections result from zoonotic transmission (“spillover”) from the Natal multimammate mouse, *Mastomys natalensis*.^1–3^ Millions of LASV spillovers are estimated to occur annually throughout West Africa,^4,5^ and although most human infections remain asymptomatic or cause only mild febrile illness,^6^ recent burden estimates suggest that LF causes approximately 24,000 hospitalisations and 4,000 deaths annually throughout the region.^7^

While no vaccines against LF are currently licensed, enrolment in the first Phase 2 clinical trial of a Lassa vaccine began in 2024, and several other vaccines are under development,^8–11^ highlighting a pressing need to assess how best to use a Lassa vaccine once licensed for human use. A recent study estimated the potential impacts of population-wide preventive Lassa vaccination campaigns.^7^ However, such whole-community, non-targeted vaccine administration is unlikely to be the most efficient strategy, and was estimated to require a low vaccine price to be cost-effective at cost-effectiveness thresholds that reflect healthcare spending opportunity costs in endemic areas.^12^

Due to such efficiency concerns, most vaccines included in national immunisation programmes preferentially target higher-risk groups. However, while children appear to be at highest risk of LASV infection,^13^ most hospitalised patients are middle-aged adults,^14^ mortality risk is estimated to increase with age,^15^ and pregnant women with LF bear particularly high risks of mortality and foetal and neonatal demise.^16^ Such risk heterogeneity makes it unclear who to target for greatest returns on vaccine programme investment, and no studies to date have assessed how Lassa vaccine impact could vary across risk groups—a critical consideration for forthcoming Lassa vaccination programmes.

In this vaccine impact modelling study, we aimed (i) to project the health-economic burden of LF from 2025-2037 across age and sex groups in subnational administrative divisions of West Africa with endemic LASV transmission and (ii) to estimate the health-economic impact and cost-efficiency of targeting Lassa vaccination to different risk groups, including children aged 2-14, women of childbearing age (WCBA) aged 15-49, all adolescents and adults aged 15-49 and older adults aged 50+.

## METHODS

### Ethics and inclusion

Ethical approval was not necessary as this model-based analysis relied only on publicly available data. The study team included LF researchers from endemic countries with whom regular virtual meetings were held. They contributed expert opinion on the value and interpretation of data sources, feedback on interim results and insights into vaccine scenarios.

### Model overview

We previously developed a mathematical model that predicts human LF burden throughout West Africa.^7^ Here, this model was extended using data from several recent and ongoing epidemiological studies to characterise LF risk across age and sex groups, and to estimate the health-economic benefits of conducting risk-targeted Lassa vaccination campaigns in areas with endemic LASV transmission. The spillover incidence map underlying the model’s infection projections is shown in **Figure 1** and an illustrated model schematic is provided in **Figure 2**.

**Figure 1.**
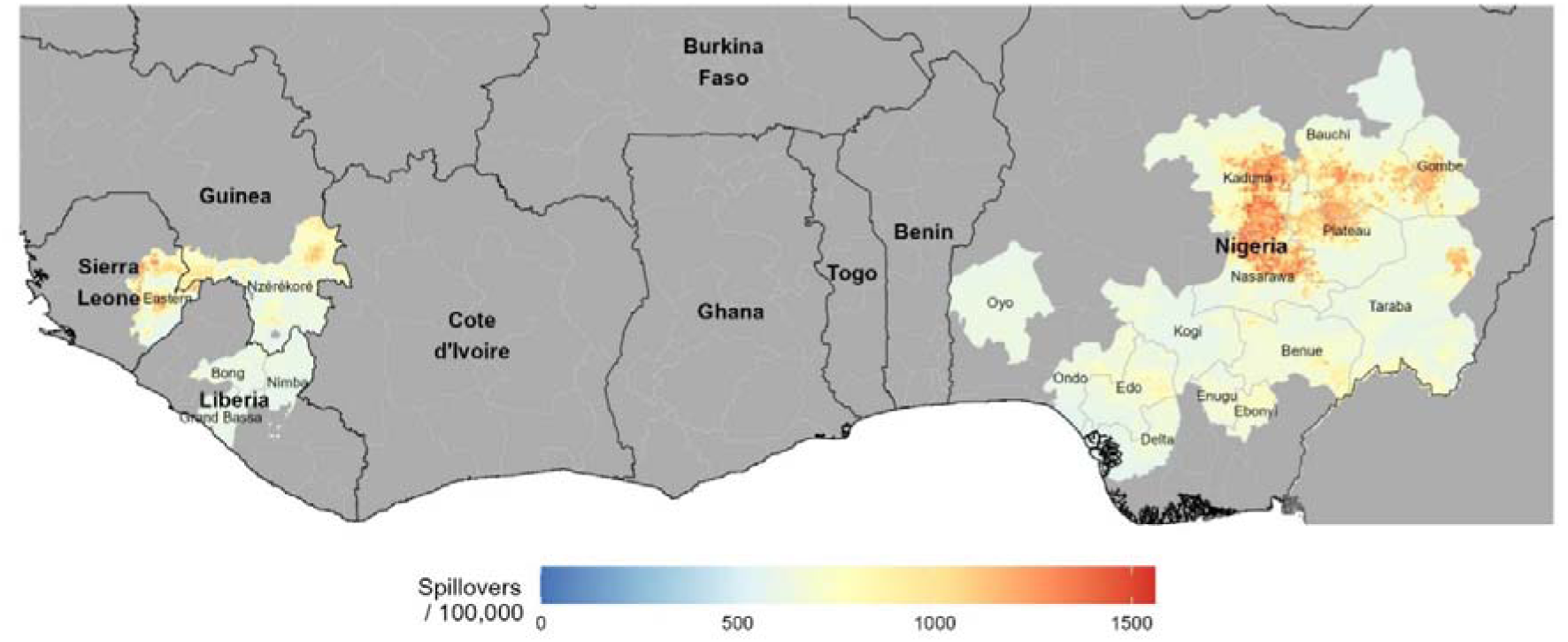
Estimated incidence of zoonotic Lassa virus infection (spillover) per 100,000 population in 2019 at the level of 0.05° by 0.05° grid cells in the 19 areas classified as endemic using WHO’s 2024 Lassa fever risk map. Zoonosis incidence was predicted in Smith et al., ^7^ building on a geospatial risk map from Basinski et al.^5^ Although the incidence estimates shown here exclude areas not classified as endemic, they result from region-wide geospatial estimation that borrows data across all West Africa.

**Figure 2.**
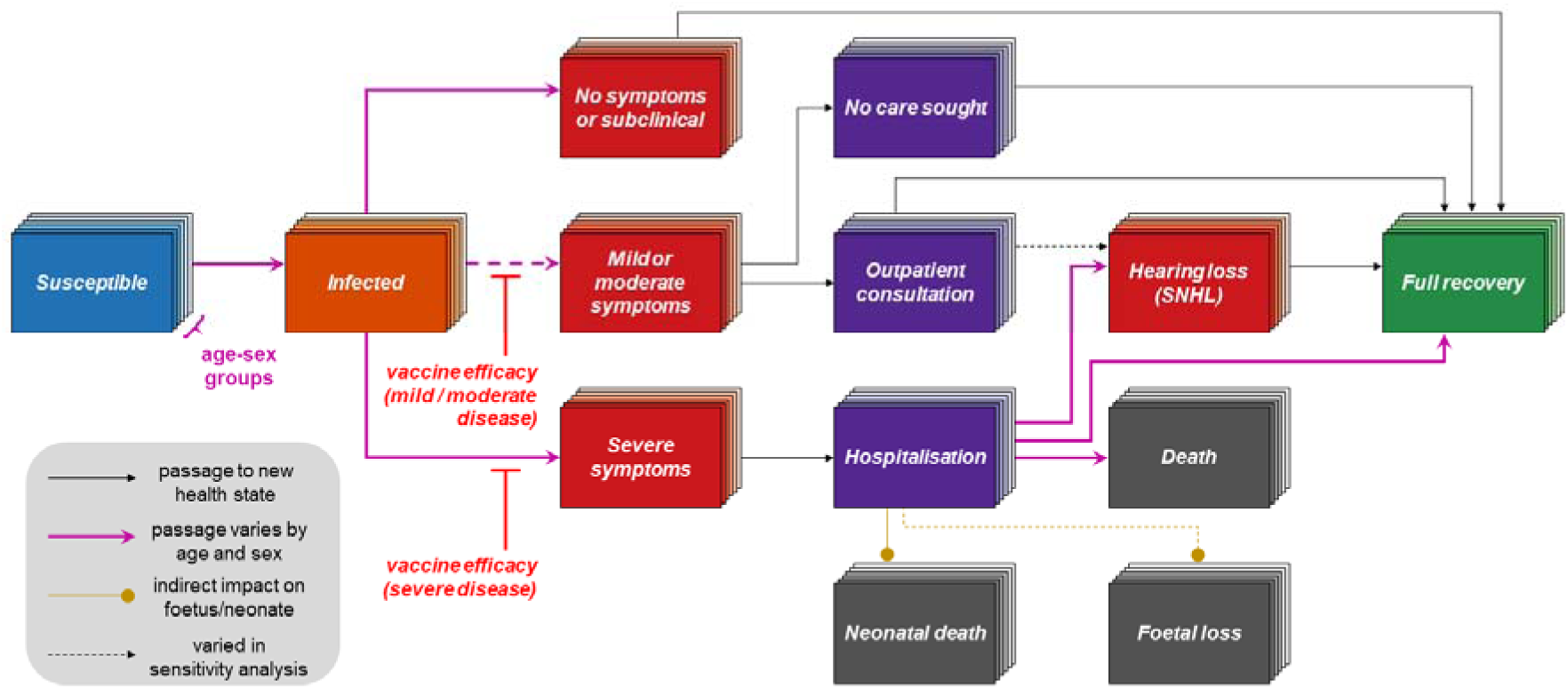
Model schematic. Flow diagram showing how individuals of different ages and sexes with Lassa virus infection move through the health states included in the model (see methods and supplementary appendix for technical details regarding model structure, parameterisation, simulation and outcome calculation). Economic outcomes associated with different health states are not shown. SNHL = sensorineural hearing loss.

### Setting and demography

Following WHO’s LF risk map from March 2024, endemic *areas* were defined as the 19 first-level subnational administrative units (e.g. states in Nigeria) reporting ≥5 LF cases annually.^17^ Population sizes and their distributions by age, sex and pregnancy status from 2025-2037 were defined using UN World Population Prospects and geospatial population estimates from WorldPop (**Figures S1, S2**).^18–20^

### Projecting infections through time

Our previous work estimated 710 (95%UI: 671-750) LASV spillover infections/100,000 population across these 19 areas in 2019 (**Table S1** ).^7^ Here, data from the ongoing Enable cohort study were harnessed to estimate age-specific spillover risk.^21^ A Bayesian serocatalytic model was fitted to age-stratified seroprevalence data from Enable’s interim results to estimate the force of infection (FOI) for human LASV infection across age groups (**Table S2** ).^13^ We assumed no LASV seroreversion, consistent with the underlying zoonosis model. Given that Enable mainly targets sites with high expected incidence, to remain conservative, age-specific incidence predictions derived from FOI estimates were scaled to match previous incidence estimates for each area from Smith *et al*.^7^

Age-specific LASV infection incidence in 2019 was multiplied by annual population projections to project forward in time. To account for seasonal infection dynamics, national-level Nigeria Centre for Disease Control and Prevention (NCDC) case data were used to distribute LASV infections at the weekly level, accounting for estimates of LF’s incubation period and lags to case reporting (**Figure S3**).

### Disease risk

A previous estimate that 19.3% (95%UI: 10.2%-32.7%) of infections coincide with symptoms (of any severity) was used, assuming no association with age or sex.^6,7^ In a sensitivity analysis, symptom risk was assumed to increase with age (**Figure S4** ). Age- and sex-specific risk of severe LF and hospitalisation was calculated by scaling a previous infection-hospitalisation risk (IHR) estimate using laboratory-confirmed hospital case data from the NCDC (**Figure S5**).^7,14^

Among hospitalised cases, age-specific case-fatality risk (CFR) was estimated using data from two arms of the LASCOPE study (**Figure S6** ).^15,22^ Increased risk of death in pregnant women relative to non-pregnant women was accounted for,^16^ while maintaining stable overall age-specific CFR in men and women, as reported in LASCOPE (**Figure S7**).^15^ Previous meta-analyses were augmented with data from LASCOPE to estimate the proportion of pregnant women hospitalised with LF who lose their foetus or experience neonatal loss (**Figures S8, S9** ).^15,16,22^ Final estimates of risks of symptomatic disease, hospitalisation and death stratified by age and sex are provided in **Table S3**.

Sensorineural hearing loss (SNHL) is a common sequela of LF that tends to occur suddenly during convalescence.^23^ Individual patient-level data from a prospective cohort study were used to quantify SNHL risk, duration and associated disability.^24^ Audiometry data from patients’ better ear at baseline and 1-year follow-up were bootstrapped and fitted to exponential decay models to estimate the average time to disability resolution (**Figures S10, S11** ). Using disability weights from the Global Burden of Disease study,^25^ a Gamma-distributed generalised additive model was used to simulate disability associated with the severity of each patient’s SNHL at baseline (**Figure S12** ). Available literature suggests that post-acute SNHL is not limited to severe cases or strongly associated with age or sex,^23,24^ so SNHL risk was applied to anyone surviving symptomatic disease, but in sensitivity analysis was limited only to those surviving hospitalisation.

### Vaccination campaigns

A range of preventive vaccination campaigns were simulated in four target groups living in Lassa-endemic areas: (i) children aged 2-14, (ii) WCBA aged 15-49, (iii) all adolescents and adults aged 15-49 and (iv) older adults aged 50+ (**Table 1** ). Untargeted vaccination administered to all individuals aged 2+ was also evaluated. Campaigns were designed as 3-year mass vaccination programmes with doses administered seasonally to align with periods of reduced LASV transmission (1 April to 31 October). Each year, campaigns covered 25% of the target group, reaching 75% coverage after 3 years with an excess 10% of doses added annually to account for wastage (**Table S4**).

**Table 1.** Total vaccine demand for target age groups in endemic areas grouped by country. The target group “all” refers to untargeted vaccination to all individuals aged 2+. The demand figures assume 75% coverage achieved over a 3-year campaign, a 1-dose schedule and 10% wastage, and account for projected population change over the vaccination period. WCBA = women of childbearing age; K = thousand; M = million.

| Target group<br>(age in years) | Guinea | Liberia | Nigeria | Sierra Leone | Total |
| --- | --- | --- | --- | --- | --- |
| Children (2-14) | 601 K | 411 K | 23.7 M | 513 K | 25.2 M |
| WCBA (15-49) | 436 K | 312 K | 16.9 M | 414 K | 18.1 M |
| Adolescents and adults<br>(15-49) | 868 K | 625 K | 34.4 M | 831 K | 36.7 M |
| Older adults (50+) | 184 K | 140 K | 7.27 M | 184 K | 7.78 M |
| All (2+) | 1.65 M | 1.18 M | 65.3 M | 1.53 M | 69.7 M |

The vaccine’s characteristics followed WHO’s target product profile for a Lassa vaccine.^26^ These include a 1-dose schedule and a 2-week delay to immunological response, after which the recipient acquires 10 years of partial protection from disease and its downstream health-economic consequences (**Figure S13** ). No estimates of vaccine efficacy are available and no correlates of protection have been identified among Lassa vaccines that have undergone assessment in clinical trials, so hypothetical vaccine efficacy assumptions were made. In the base case analysis, vaccine was 70% effective against all symptomatic disease, but vaccine efficacies of 50%, 70% and 90% were considered and scenarios with greater vaccine efficacy against severe disease than mild/moderate disease were included. There was assumed to be no association between vaccination uptake and infection risk or serostatus.

### Model outcomes

Our model was used to predict baseline health-economic outcomes in the absence of vaccination, as well as outcomes occurring in vaccinated individuals and hence averted proportionally to vaccine efficacy. Final outcomes in each area are aggregated at the level of sex, pregnancy status and age group, and are reported both annually and as cumulative totals over the full model horizon from 2025-2037. Remaining model parameters are detailed in **Tables S5 and S6.**

Health outcomes include: (i) zoonotic LASV infections, (ii) cases of LF, defined as reported or unreported symptomatic LASV infections and stratified into mild/moderate cases or severe cases, (iii) LF hospitalisations, (iv) LF deaths (including neonatal deaths), (v) foetal losses, (vi) cases of SNHL among LF survivors, and (vii) disability-adjusted life-years (DALYs).

Economic outcomes include: (i) healthcare costs, stratified by setting (outpatient/inpatient) and payer (government-reimbursed/out-of-pocket (OOP)); (ii) instances of catastrophic healthcare expenditure or impoverishing healthcare expenditure resulting from OOP healthcare costs; (iii) productivity losses due to reduced labour force participation because of acute symptomatic disease, SNHL or death (see **Figures S14 to S17** ); (iv) monetised DALYs, quantified using country-specific health opportunity costs;^12^ (v) the value of statistical life (VSL) and value of statistical life-years (VSLY) lost due to LF mortality; and (vi) threshold vaccine costs (TVCs), representing the price per vaccine dose at which the benefit-to-cost ratio equals 1. Societal costs are presented as the sum of all healthcare costs, productivity losses and monetised DALYs, and in the base case analysis TVCs are calculated using societal costs, but VSLY is used instead in sensitivity analysis. Costs beyond the first year of the study horizon (2025) are discounted at 3.5% annually, or 0% in sensitivity analysis. Monetary outcomes are reported in International dollars (I$) 2023.

### Simulation and statistical reporting

Monte Carlo simulations were conducted by randomly drawing input parameters from estimated distributions for 500 model runs. Parameters drawn from distributions include: (i) area-level zoonosis incidence rates; (ii) age-stratified LASV seroprevalence and FOI; and (iii) the probabilities, durations and disability weights associated with different health states. Final health-economic outcomes, as well as outcomes averted by vaccination, are reported as means and 95% uncertainty intervals (95%UIs) of outcome distributions across all model runs. Partial rank correlation coefficients were calculated to assess impacts of input parameter uncertainty on outcome uncertainty. Burden estimates are reported following the GATHER statement (**Table S7**).

### Code and data sharing

All analyses were run using R version 4.3.0. The code and minimum dataset required to reproduce results are available at www.github.com/drmsmith/lassaRiskVac.

## RESULTS

### LF burden without vaccination

From 2025-2037 in the 19 included endemic areas, our model predicted a cumulative 8.68 million (95%UI: 8.19 million-9.15 million) human LASV infections. At the annual level, growing populations in endemic regions coupled with our assumption of stable annual transmission risk from the rodent host translated to gradually increasing LASV infections year-over-year, from 609,000 (95%UI: 575,000-643,000) in 2025 to 724,000 (95%UI: 683,000-763,000) in 2037 (**Figure 3**). In the absence of vaccination, these infections led to a substantial burden of hospitalisations, deaths and sequelae, resulting in a cumulative 401,000 (95%UI: 228,000-672,000) DALYs (**Table 2**).

**Figure 3.**
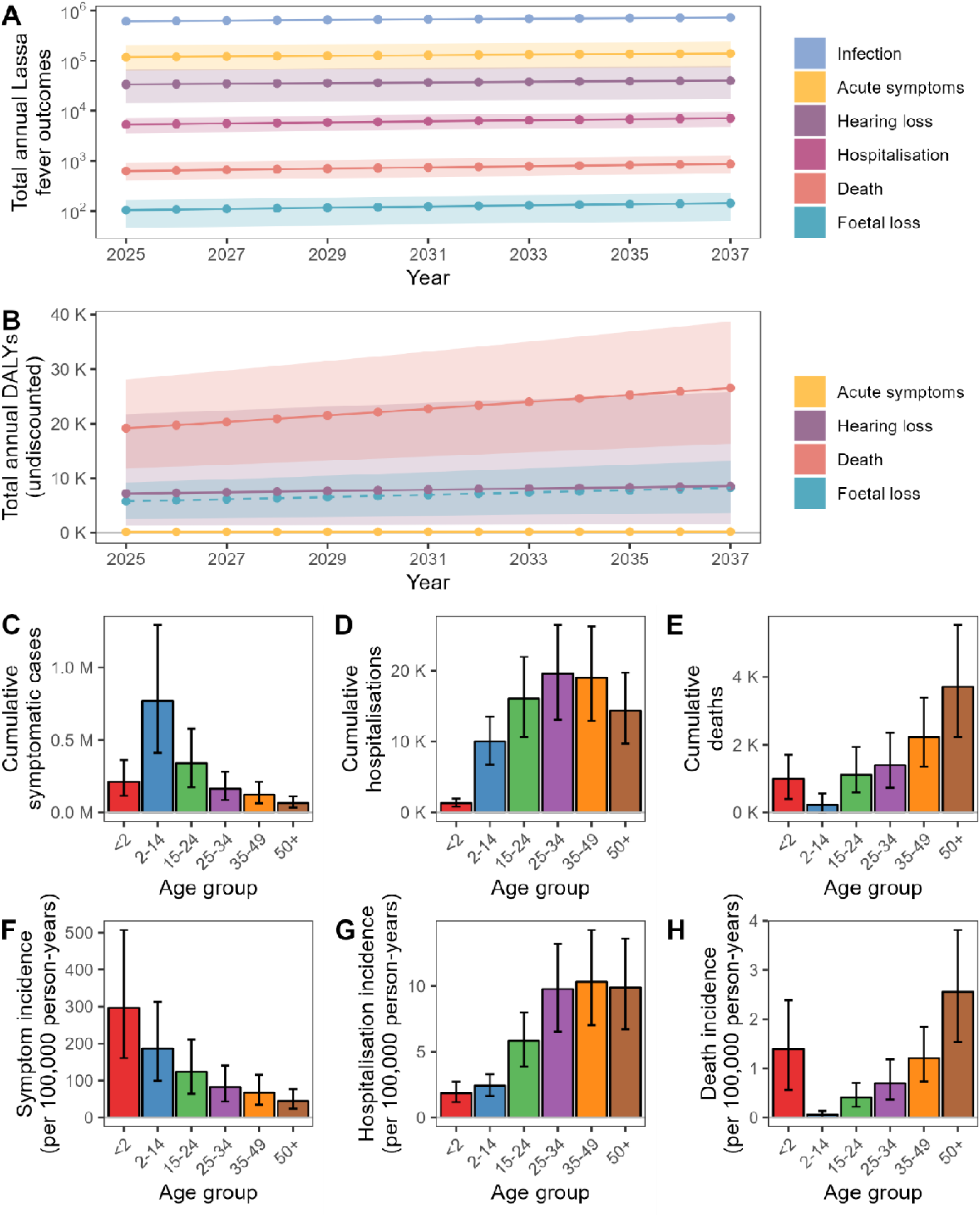
Projected burden of Lassa fever from 2025 to 2037 in the absence of vaccination and associations with age. (**A**) Change over time in the annual number of infection outcomes (log scale). Acute symptoms refers to all symptomatic cases. (**B**) Change over time in annual DALYs associated with Lassa fever. DALYs due to acute symptoms combine those due to mild/moderate disease and severe disease. DALYs due to foetal loss (dashed line) are only included in sensitivity analysis. (**C**) The cumulative total number of symptomatic cases, stratified by age group. (**D**) The cumulative total number of Lassa fever hospitalisations, stratified by age group. (**E**) The cumulative total number of Lassa fever deaths, stratified by age group. (**F**) The cumulative incidence of symptomatic cases per 100,000 person-years, stratified by age group. (**G**) The cumulative incidence of Lassa fever hospitalisation per 100,000 person-years, stratified by age group. (**H**) The cumulative incidence of Lassa fever death per 100,000 person-years, stratified by age group. In panels A and B, lines represent means and shading represents 95% uncertainty intervals. In panels C through G, bar heights represent means and error bars represent 95% uncertainty intervals. Neonatal deaths occurring subsequent to maternal Lassa fever are counted as deaths in children aged <2. Foetal losses are not counted as deaths. DALY = disability-adjusted life-year.

**Table 2.** Projected cumulative total health and economic burden of Lassa fever from 2025 to 2037 in the absence of vaccination. DALYs exclude DALYs due to foetal loss. Future costs and life-years are discounted annually at 3.5%/year. Symptomatic cases include both mild/moderate and severe symptomatic infections. DALY = disability-adjusted life-year, VSL = value of statistical life, VSLY = value of statistical life-years, I$ = International dollar, K = thousand, M = million, B = billion.

| Outcome | Mean (95% uncertainty interval)<br>cumulative totals | Mean (95% uncertainty interval)<br>cumulative totals per 100,000<br>person-years |
| --- | --- | --- |
| *Health burden* |  |  |
| Lassa virus infection (n) | 8.68 M (8.19 M - 9.15 M) | 674 (636 - 711) |
| Symptomatic cases (n) | 1.67 M (884 K - 2.86 M) | 130 (68.6 - 222) |
| Hospitalisation (n) | 80.3 K (54.2 K - 108 K) | 6.23 (4.21 - 8.42) |
| Sensorineural hearing loss (n) | 477 K (204 K - 933 K) | 37.1 (15.8 - 72.4) |
| Death (n) | 9.66 K (6.22 K - 14.2 K) | 0.75 (0.48 - 1.10) |
| Foetal loss (n) | 1.60 K (710 - 2.56 K) | 0.12 (0.06 - 0.20) |
| DALYs (n) | 401 K (228 K - 672 K) | 31.1 (17.7 - 52.2) |
| *Economic burden* |  |  |
| Healthcare costs<br>(I\$ 2023) | 177 M (116 M - 247 M) | 13.7 K (8.99 K - 19.2 K) |
| Catastrophic healthcare<br>expenditure (n) | 80.1 K (54.1 K - 108 K) | 6.22 (4.20 - 8.40) |
| Impoverishing healthcare<br>expenditure (n) | 47.4 K (32.0 K - 64.0 K) | 3.68 (2.49 - 4.97) |
| Productivity losses<br>(I\$ 2023) | 706 M (402 M - 1.14 B) | 54.9 K (31.2 K - 88.4 K) |
| Monetised DALYs<br>(I\$ 2023) | 39.9 M (21.9 M - 74.2 M) | 3.10 K (1.70 K - 5.76 K) |
| VSL (I\$ 2023) | 2.53 B (1.63 B - 3.74 B) | 197 K (127 K - 291 K) |
| VSLY (I\$ 2023) | 1.38 B (852 M - 2.03 B) | 107 K (66.2 K - 158 K) |

The cumulative economic burden of LF in terms of societal costs—the sum of healthcare costs, productivity losses and monetised DALYs—was estimated at I$923 million (95%UI: I$555 million-I$1.42 billion). An alternative measure of LF’s economic burden, VSLY, was estimated at I$1.38 billion (95%UI: I$852 million-I$2.03 billion). Inpatient treatment costs far outnumbered outpatient treatment costs, and LF deaths were responsible for the majority of productivity losses and monetised DALYs (**Table S9**). However, hearing loss, which in our base case analysis was assumed to result from on average 29% of symptomatic cases, made major contributions to productivity losses (25%) and monetised DALYs (37%).

The projected burden of LF varied greatly across age groups (**Figure 3** ). Annual LASV infection incidence per 100,000 population decreased with age, from 1,540 (95%UI: 1,320-1,770) in children aged <2 years to 232 (95%UI: 207-255) in those aged 50+ (**Table S8**). Children aged 2-14 experienced nearly half (46%) of all infections, although the proportion of infections occurring in different age groups varied slightly through time (**Figure S18**). In contrast to highest infection burden in youngest age groups, middle-aged adults accounted for the most hospitalisations, while older adults aged 50+ accounted for the most deaths. Pregnant women accounted for 2.1% of all symptomatic cases but a disproportionally large share (5.6%) of deaths (**Figure S19** ). Children <2 were second only to adults aged 50+ in terms of mortality risk per person-year, due to both a high estimated CFR and a mean estimated 38% of live births in pregnant women hospitalised with LF resulting in neonatal death. Total DALYs were similar across age groups but DALYs per 100,000 person-years were greatest in children <2 (**Figure S20**).

### Vaccine impact

Modelled vaccination campaigns were implemented seasonally over a 3-year period with immunity assumed to last for 10 years, resulting in heterogeneous vaccine impacts through time (**Figure 4, Figures S21 to S24**). Overall, targeting children aged 2-14 averted the most symptomatic cases, while targeting adolescents and adults aged 15-49 averted the most hospitalisations, deaths and DALYs (**Figure 5, Figure S25** ). However, these two target groups also required the most vaccine doses (**Table 1** ). When accounting for vaccine benefits relative to the number of doses administered, the most efficient strategy depended on the health outcome considered (**Figure 5**). The most efficient strategy for preventing symptomatic disease was targeting children aged 2-14, for preventing hospitalisations was targeting adolescents and adults aged 15-49, for preventing deaths was targeting older adults aged 50+, and for preventing DALYs was targeting WCBA. In contrast to targeted vaccination (**Figure S26** ), for untargeted vaccination the age distributions of vaccine benefits resembled the underlying age distributions of disease (**Figure S27** ). Projected vaccine impacts depended importantly on vaccine efficacy assumptions (**Table S10**).

**Figure 4.**
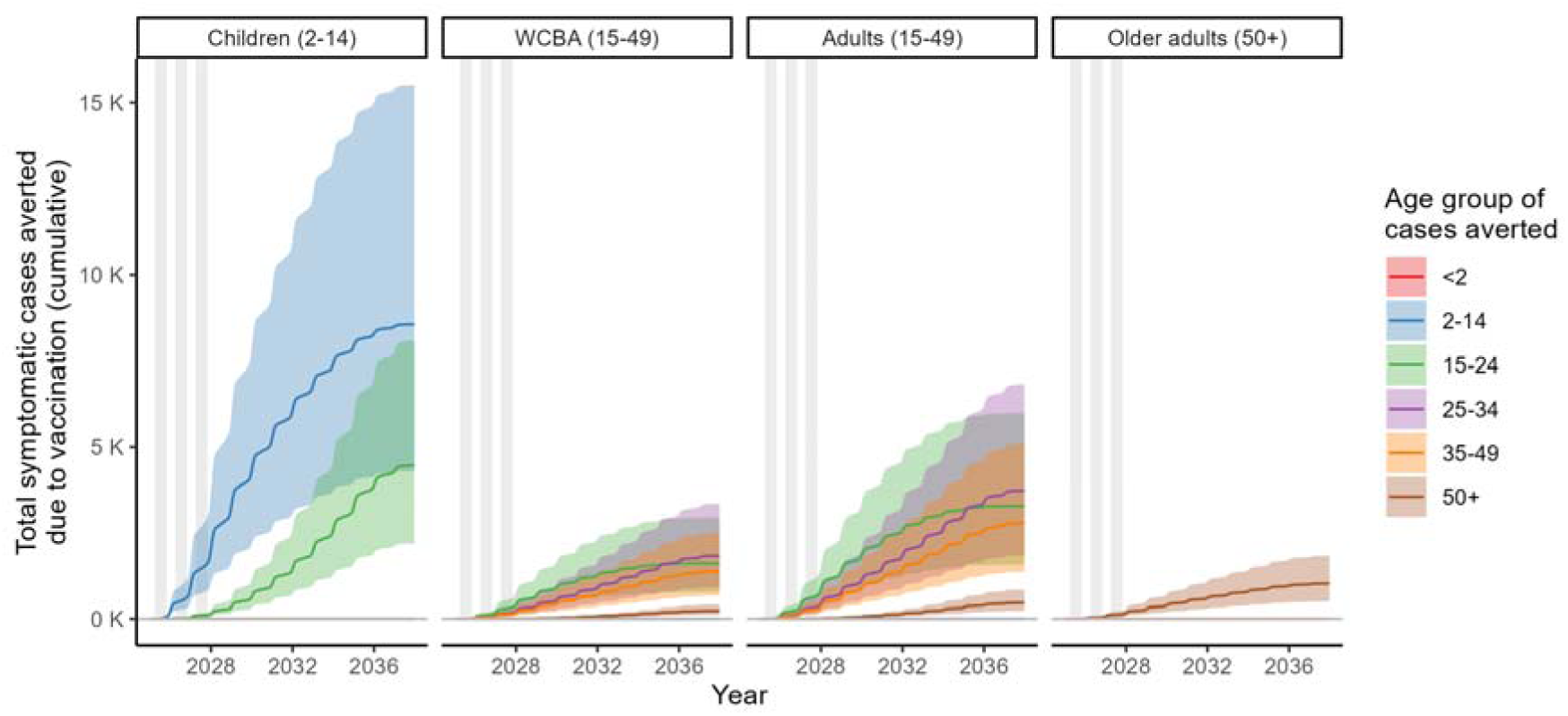
Evolution of age-specific vaccine impacts through time. The weekly total cumulative number of symptomatic Lassa fever cases in different age groups (colours) averted due to vaccination in one exemplar area (Edo, Nigeria) from 1 January 2025 to 31 December 2037. Grey vertical bars indicate periods of Lassa vaccine distribution and panels compare results when targeting different groups for vaccination. Thick lines and shaded areas represent means and 95% uncertainty intervals, respectively, for a 1-dose vaccine 70% effective against all symptomatic disease and an immunological response lasting 10 years. The target group “Adults” includes adolescents and adults. WCBA = women of childbearing age.

**Figure 5.**
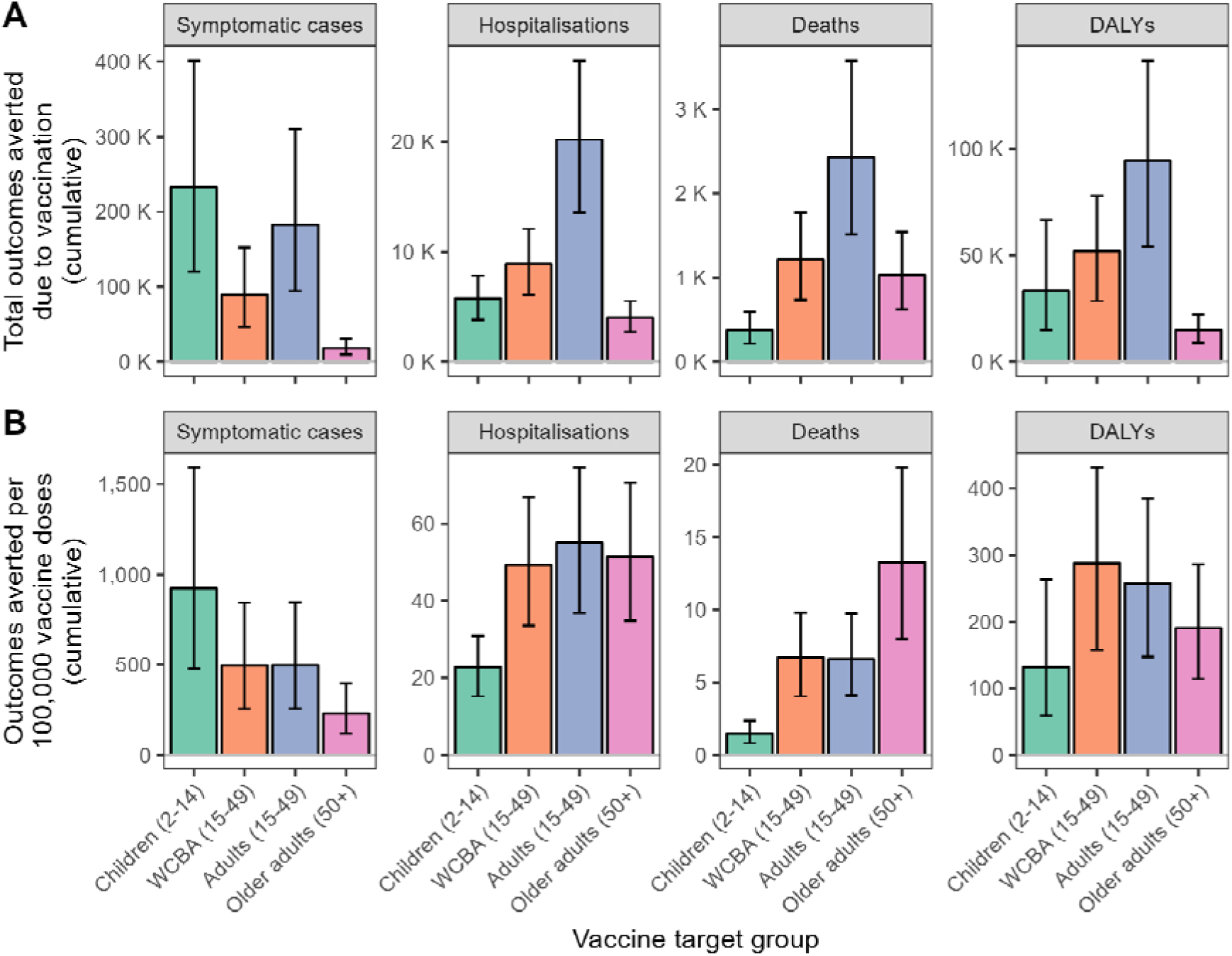
Comparing the cumulative health impacts and efficiencies of targeting different risk groups for vaccination with a 1-dose Lassa fever vaccine having 70% efficacy against all symptomatic disease. (**A**) Impact: cumulative total health outcomes averted due to vaccination across all 19 areas from 2025 through to 2037. (**B**) Efficiency: cumulative total health outcomes averted per 100,000 doses of vaccine administered . exclude DALYs due to foetal loss. Error bars represent 95% uncertainty intervals. The target group “Adults” includes adolescents and adults. WCBA = women of childbearing age.

Economic benefits of targeted vaccination campaigns were always greatest when targeting adolescents and adults aged 15-49, under base case assumptions preventing I$253 million (95%UI: I$148 million-I$396 million) in societal costs, compared to I$102 million (95%UI: I$59.4 million-I$163 million) when targeting WCBA, I$48.5 million (95%UI: I$30.3 million-I$71.5 million) when targeting older adults and I$43.8 million (95%UI: I$25.0 million-I$69.7 million) when targeting children (**Table 3**). However, the most cost-efficient strategy in terms of economic benefits of vaccination again depended on the outcome considered (**Figures S28, S29**). Targeting adolescents and adults was most efficient in terms of healthcare costs and productivity losses, while targeting WCBA was most efficient in terms of monetised DALYs and VSLY. Targeting children was by a considerable margin the least efficient strategy across all economic outcomes considered (**Tables S11, S12** ). As expected, untargeted vaccination led to the greatest overall economic benefits, equivalent to the sum of benefits from vaccinating children aged 2-14, adolescents and adults aged 15-49 and older adults aged 50+, but with intermediate efficiency relative to targeted strategies.

**Table 3.**
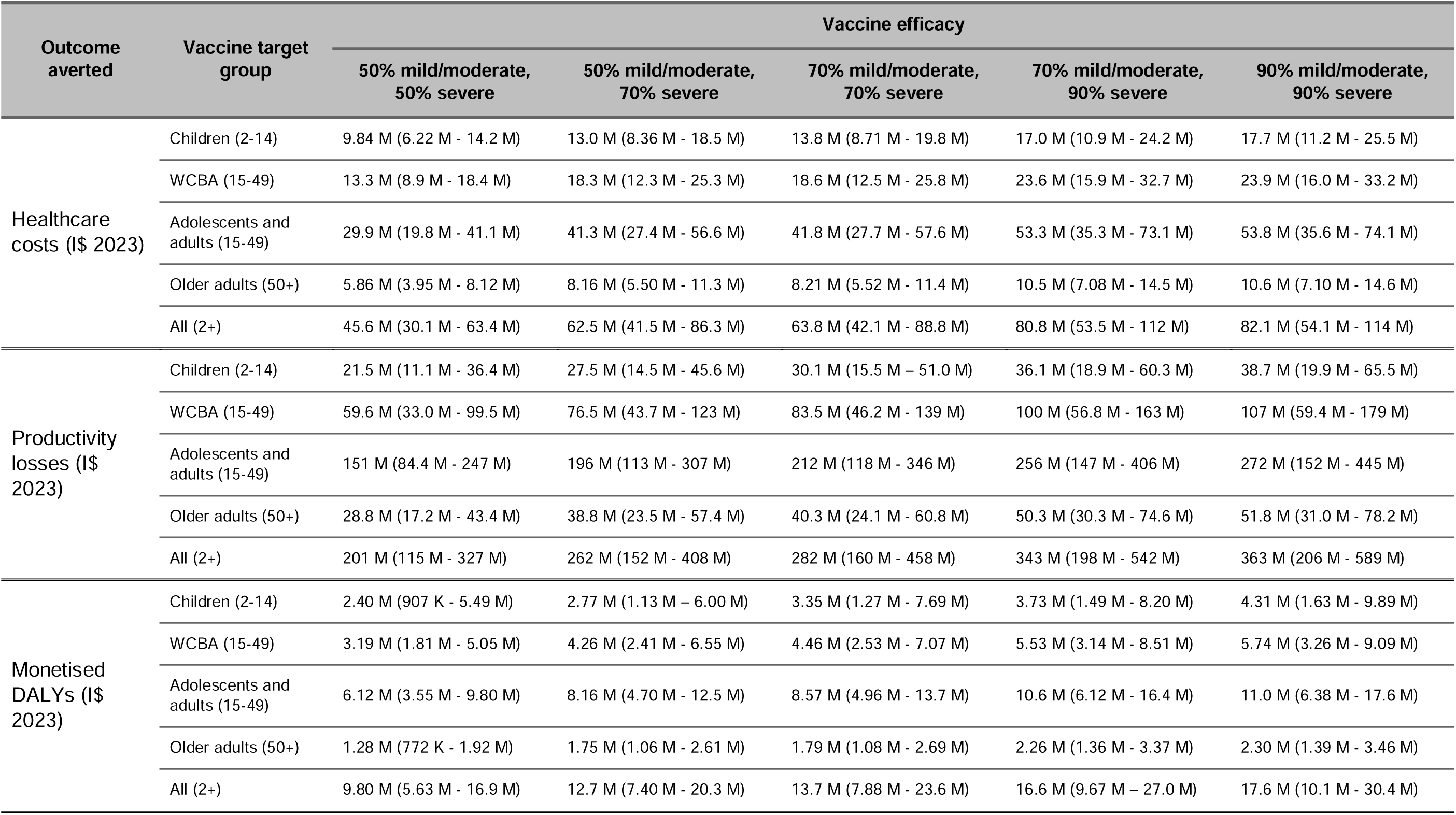
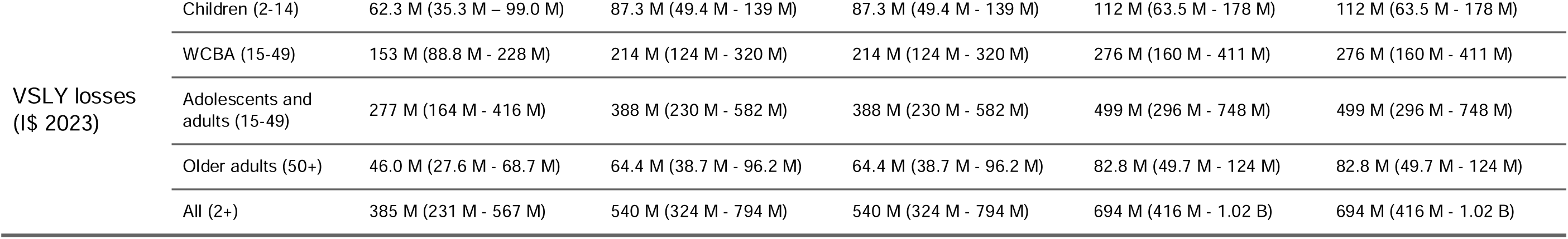
Projected cumulative total economic costs averted due to Lassa vaccination from 2025 to 2037, depending on the group targeted for vaccination and the vaccine’s efficacy against mild/moderate and severe disease. Monetised DALYs exclude DALYs due to foetal loss. Future costs and life-years are discounted annually at 3.5%/year. DALY = disability-adjusted life-year, VSLY = value of statistical life-years, WCBA = women of childbearing age, I$ = International dollar, K = thousand, M = million, B = billion.

Under base case assumptions, the highest TVC for a single-dose vaccine was estimated at I$7.39 (95%UI: I$4.33-I$11.60) when targeting adolescents and adults, followed by I$6.69 (95%UI: I$4.17-I$9.85) when targeting older adults, I$6.10 (95%UI: I$3.56-I$9.74) when targeting WCBA, I$5.34 (95%UI: I$3.18-I$8.23) for untargeted vaccination and I$1.94 (95%UI: I$1.10-I$3.10) when targeting children (**Table 4**). This order of vaccine target groups by TVC was robust to sensitivity analyses with three exceptions: (i) older adults aged 50+ yielded highest TVCs if SNHL is assumed only to occur after severe disease, (ii) WCBA yielded second-instead of third-highest TVCs if not discounting future costs, and (iii) if considering averted VSLY losses instead of averted societal costs as the metric of vaccine benefit, TVCs were highest when targeting WCBA and lowest when targeting older adults.

**Table 4.**
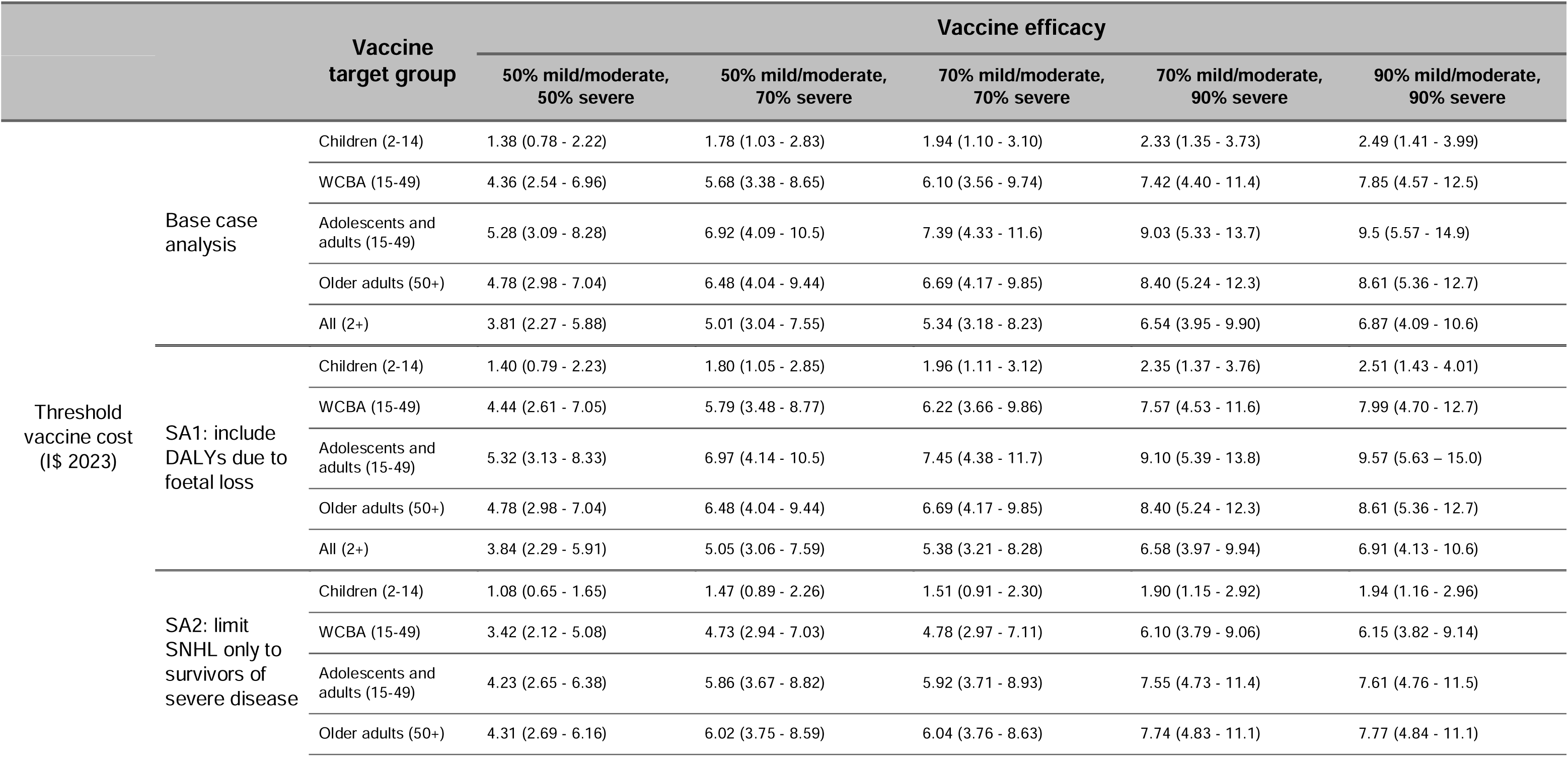
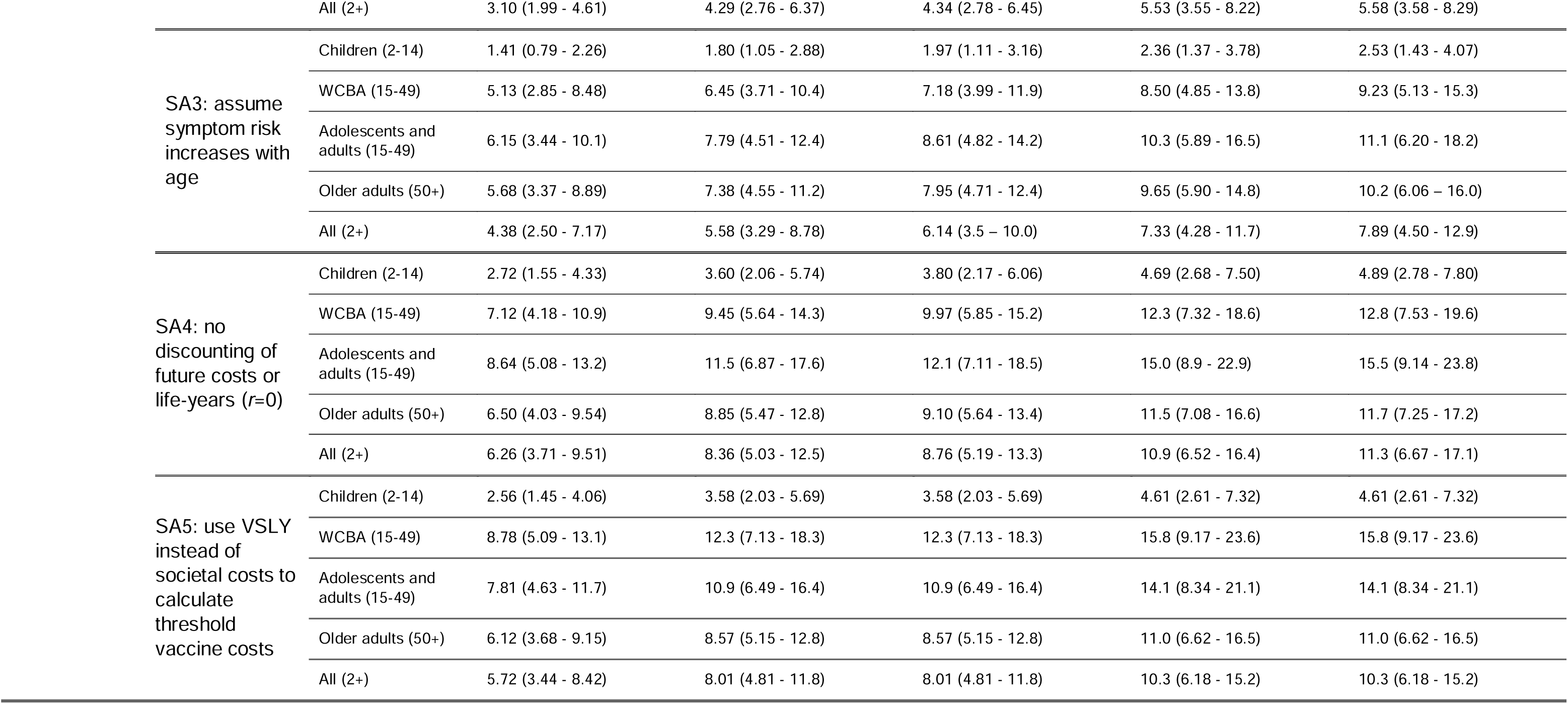
Threshold vaccine costs estimated by dividing the cumulative total societal costs averted by vaccination by the number of doses administered to different target groups, in the base case analysis compared to various sensitivity analyses. DALY = disability-adjusted life-year, VSLY = value of statistical life-years, WCBA = women of childbearing age, SNHL = sensorineural hearing loss, I$ = International dollar, SA = sensitivity analysis.

## DISCUSSION

This study projected the burden of LF in endemic regions of West Africa from 2025-2037 and estimated the health-economic benefits of targeting Lassa vaccination to different groups based on age and sex. Benefits of vaccination varied greatly across target groups, reflecting heterogeneity in underlying risk. Targeting adolescents and adults regardless of sex was the most efficient strategy for preventing healthcare costs and productivity losses, while targeting WCBA was most efficient for preventing DALYs and VSLY losses. Children bore the greatest infection burden but were by far the least cost-efficient target for limited doses due to their low estimated risks of severe disease and death upon infection. Across a range of scenarios and sensitivity analyses, TVCs were highest when targeting adolescents and adults aged 15-49, followed by older adults aged 50+, suggesting that prioritising these groups would lead to greatest health-economic returns on vaccine investment from a societal perspective. However, the most efficient target group could vary if vaccine efficacy were to vary across risk groups, for instance if vaccine-induced immune responses vary with age or provide protection against particular outcomes, such as hearing loss or neonatal demise, more than others. Monitoring vaccine efficacy against different outcomes across population groups in clinical trials will therefore be crucial to inform future decision-making around target prioritisation.

We synthesised data from several recent and ongoing prospective cohort studies to estimate LF risk across age and sex groups. Interim results from the Enable study—the largest prospective community-based multi-country LF cohort study—provided an opportunity to estimate age-specific infection risk,^13,21^ while data from the LASCOPE study in Nigeria,^15^ including new results from the paediatric cohort,^22^ provided further opportunities to characterise age- and sex-specific risks. We also accounted for aspects of LF’s health-economic burden not considered previously, including data-driven characterisation of the risk, duration, quality-of-life impacts and economic consequences of post-acute SNHL.^24,25^

LF is concentrated in areas that face a challenging burden of many diseases,^27^ and there are opportunity costs to investing in Lassa vaccination in lieu of alternative interventions. Estimates of vaccine programme cost-effectiveness are therefore critical to guide Lassa vaccine investment decisions. Under base case assumptions, TVC estimates ranged from I$1.94/dose when targeting children to I$7.39/dose when targeting adolescents and adults. The TVC for untargeted vaccination (I$5.34/dose) was about 32% higher than our previous estimate for a population-wide campaign in endemic areas under similar vaccine efficacy assumptions (I$2.03 for a 2-dose schedule, translating to approximately I$4.06 for a 1-dose schedule), which had a shorter time horizon and did not account for future productivity losses due to SNHL.^7^ Across 18 vaccines in 73 GAVI-eligible countries, Portnoy et al. estimated that the highest price for any vaccine (pneumococcal conjugate) was $6.79/dose in US$2010.^28^ While the future prices of any forthcoming Lassa vaccines are not known, our results highlight why Lassa vaccination may only be cost-effective if targeted appropriately to groups yielding the greatest health-economic returns.

Some elements of LF’s burden remain poorly characterised. Data describing mild disease and post-acute sequelae are particularly scarce.^29^ Three parameters that contributed most to economic outcome uncertainty were symptomatic disease risk, SNHL risk and SNHL duration (Figure S30). We conducted sensitivity analyses around these parameters. Accounting for an alternative age structure in mild/moderate disease risk did not affect which vaccination strategies were most cost-efficient. However, when SNHL risk was limited only to severe cases, targeting older adults aged 50+ was the most cost-efficient strategy. Further, average DALY incidence increased from 31 DALYs/100,000 person-years to 38/100,000 in a sensitivity analysis including foetal loss DALYs, which assumed each lost foetus would have lived to average life expectancy, thereby representing an upper bound to potential foetal loss DALYs.^30^ Despite these uncertainties, our main conclusions regarding vaccine efficiency and prioritisation were robust to sensitivity analyses.

Two remaining uncertainties relate to the previously reported zoonosis risk map used to estimate LASV incidence.^5,7^ First, distinct LASV lineages were not explicitly accounted for, although there is suggestion of variable disease severity across regions,^31^ and vaccine efficacy could vary across populations if any vaccine-mediated immune responses are lineage-specific.^32,33^ Similarly, it is unclear whether age- and sex-specific risks of hospitalisation and death, which were quantified using Nigerian data, differ in Guinea, Liberia or Sierra Leone. While potential differences are unlikely to influence our conclusions, as >90% of predicted infections occurred in Nigeria, vaccine target prioritisation decisions in the Mano River region should nonetheless take local epidemiological patterns into account. Second, whilst we propagate incidence uncertainty throughout our simulations, the map’s predictive accuracy is difficult to validate, as few representative population-based studies have quantified infection risk throughout LASV’s endemic range.^34^ Another, more recent bottom-up geospatial modelling study provides an opportunity for comparison. Across 19 endemic areas, we estimated 130 (95%UI: 69-222) symptomatic cases/100,000 person-years, while Moore et al. estimated 80 to 390 cases/100,000 across the 20 highest incidence areas (when also assuming no seroreversion and full immunity among seropositives).^35^ Although some degree of LASV seroreversion does occur, seroreverted individuals are believed to have reduced LF risk relative to the serologically naïve, so to remain conservative we only considered a model without seroreversion, therefore likely underestimating infection incidence. However, by fitting IHRs and CFRs to laboratory-confirmed hospital case data from Nigerian states with extensive LF surveillance programmes in place, we ensured that our model faithfully reproduces best-available prospectively collected age- and sex-stratified estimates of annual hospitalisations and deaths—the main predicted drivers of LF’s health-economic burden.

In conclusion, our analysis has provided detailed LF burden estimates stratified by age and sex, allowing us for the first time to evaluate the potential impacts and cost-effectiveness of Lassa vaccination strategies targeting different population groups. These results may inform prioritisation strategies for forthcoming Lassa vaccines.

## Acknowledgments

This work was conducted by the OxLiv Consortium and funded by the Coalition for Epidemic Preparedness Innovations (CEPI) through Vaccine Impact Assessment project funding. We acknowledge the CEPI project team (project lead: Christinah Mukandavire) and the CEPI Lassa Disease Programme (lead: Katrin Ramsauer) for their continuous support and helpful discussions, and the project’s external advisory team for their invaluable feedback. KBP and DRMS are supported by the Medical Research Foundation (MRF-160-0017-ELP-POUW-C0909). TDH thanks the Li Ka Shing Foundation for institutional funding. DA, OOA, BNA, AC, WAF, DSG, KJ, SAO, JS and DAW would like to acknowledge CEPI for funding the Enable Lassa research programme in West Africa. We thank Paul Bessell for visualising Figure 1 and Emily Nixon for helpful conversations about this work. The views expressed are those of the authors and not necessarily those of the institutions with which they are affiliated.

## Data sharing

All data used in this study are publicly available. The code and minimum dataset required to reproduce results are available at www.github.com/drmsmith/lassaRiskVac.

## Contributions

KBP and TDH acquired funding and supervised the work. AAT administered the project. DRMS, MCAO, HRS and KMH reviewed the literature and synthesised data. DA, OOA, BNA, AC, WAF, DSG, KJ, SAO, JS and DAW provided expert input on Lassa fever epidemiology and the value and interpretation of data. DRMS and AAT developed the vaccine administration strategies. KBP conducted serocatalytic modelling. DRMS developed all other model components, conducted analyses and produced results. DRMS, KBP, TDH and AAT interpreted results. The underlying data were verified by DRMS, KBP and TDH, and all authors had full access to the study data and accept responsibility to submit for publication. DRMS wrote the first draft. The final version of this manuscript was reviewed and approved by all authors.

