## Supplementary appendix for "Health-economic impacts of age- and sex-targeted Lassa fever vaccination in endemic regions of Nigeria, Guinea, Liberia and Sierra Leone: a modelling study"

*by:* ***David R M Smith et al.***

**CONTENTS:**

1. **Model calibration and parameterisation**
   a. Setting and demography
   b. Age-specific infection risk
   c. Projecting infections through time
   d. Symptomatic Lassa fever risks stratified by age, sex and pregnancy status
   e. Lassa fever-induced sensorineural hearing loss
   f. Vaccination campaigns
   g. Other health state parameters
   h. Economic parameters
2. **Outcome calculations**
   a. Disease states
   b. Disability-adjusted life years (DALYs)
   c. Treatment costs
   d. Monetised DALYs
   e. Productivity losses
   f. Societal costs
   g. Value of statistical life-years (VSLY)
   h. Threshold vaccine costs (TVCs)
3. **GATHER checklist**
4. **Supplementary results**
   a. Projected Lassa fever burden in the absence of vaccination
   b. Vaccine impact
   c. Partial rank correlation coefficients

***APPENDIX 1: MODEL CALIBRATION AND PARAMETERISATION***

We extended a previously developed epidemiological model, which predicts human Lassa fever burden across West Africa,^1^ to estimate the health and economic impacts of a range of risk-targeted Lassa fever vaccination campaigns targeting different age and sex groups. We extended this model by: (i) focusing on areas of West Africa classified as endemic for Lassa fever in an updated risk map from the World Health Organization (WHO); (ii) accounting for the distributions of age, sex and pregnancy status in each area, projected forward through time from 2025 to 2037; (iii) using seroprevalence data from the Enable study to estimate age-specific infection risk; (iv) using case data from the Nigeria Centre for Disease Control and Prevention (NCDC) to predict infection seasonality and age- and sex-specific infection-hospitalisation risk (IHR); (v) using clinical outcome data from the *Lassa fever outcomes and prognostic factors in Nigeria* (LASCOPE) study to predict age- and sex-specific case-fatality risk (CFR); (vi) updating meta-analyses to quantify risks of mortality, foetal loss and neonatal death in pregnant women hospitalised with Lassa fever; (vii) analysing individual patient-level audiometry data to quantify the risk, duration and disability of Lassa fever-induced SNHL; (viii) accounting for age- and sex-specific productivity losses, including those resulting from SNHL; (ix) using age-stratified mortality projections to calculate the value of statistical life-years (VSLY) lost due to Lassa fever; and (x) developing updated risk-targeted vaccine rollout scenarios considering distinct estimates of vaccine efficacy against mild and severe disease.

We adapted the model to focus only on zoonotic infection, as human-to-human transmission is primarily associated with healthcare settings—although substantial nosocomial outbreaks are rare—and is estimated to account for only a small minority of total human LASV infections, even in geographically constrained outbreak contexts.^2-4^

***1a. Setting and demography***

The 19 *areas* classified as Lassa fever endemic include 14 states in Nigeria (Bauchi, Benue, Delta, Ebonyi, Edo, Enugu, Gombe, Kaduna, Kogi, Nasarawa, Ondo, Oyo, Plateau and Taraba), 3 counties in Liberia (Bong, Grand Bassa and Nimba), 1 region in Guinea (Nzérékoré) and 1 province in Sierra Leone (Eastern Province).^5^

The population sizes of each area and their distributions by age, sex and pregnancy status were projected from 2025 to 2037. First, UN World Population Prospects (2024 revision) by single year ages (medium variant) were used to define the number of males and females in each 1-year age-band living in each country from 2025 to 2037, where all individuals aged 80+ were binned together.^6^

To account for pregnancy, national-level UN estimates from 2023 of the number of women in different age groups giving birth annually were divided by the number of women in each corresponding country-age group in 2023.^7^ These proportions were assumed to be stable, whereby in population projections the number of women having a live birth from 2025 to 2037 was calculated by multiplying these annual age-specific live birth probabilities by the corresponding number of women in each country-age group.

Finally, the proportion of each country’s population living in each area was quantified by extracting rasterised UN-adjusted population estimates from WorldPop in 2020 for each country and dividing the number of individuals in each area by the national population size.^8^ National-level population projections were then multiplied by these proportions to arrive at area-level population projections, where each country’s distributions of age, sex and pregnancy status were assumed to be the same in each area within that country and stable through time.

All analyses were conducted at the level of single year age-bands before aggregation into age groups for final presentation of results. In all cases where model input data were stratified by age groups, data were translated to single-year estimates, accounting for the underlying population distributions in the corresponding age group in each area or country, depending on the level of reporting.

A cross-section of projected population pyramids for target age groups in each area in 2025 are shown in **Figure S1**, and estimates of the proportion of women of different ages having a live birth annually are shown in **Figure S2**.

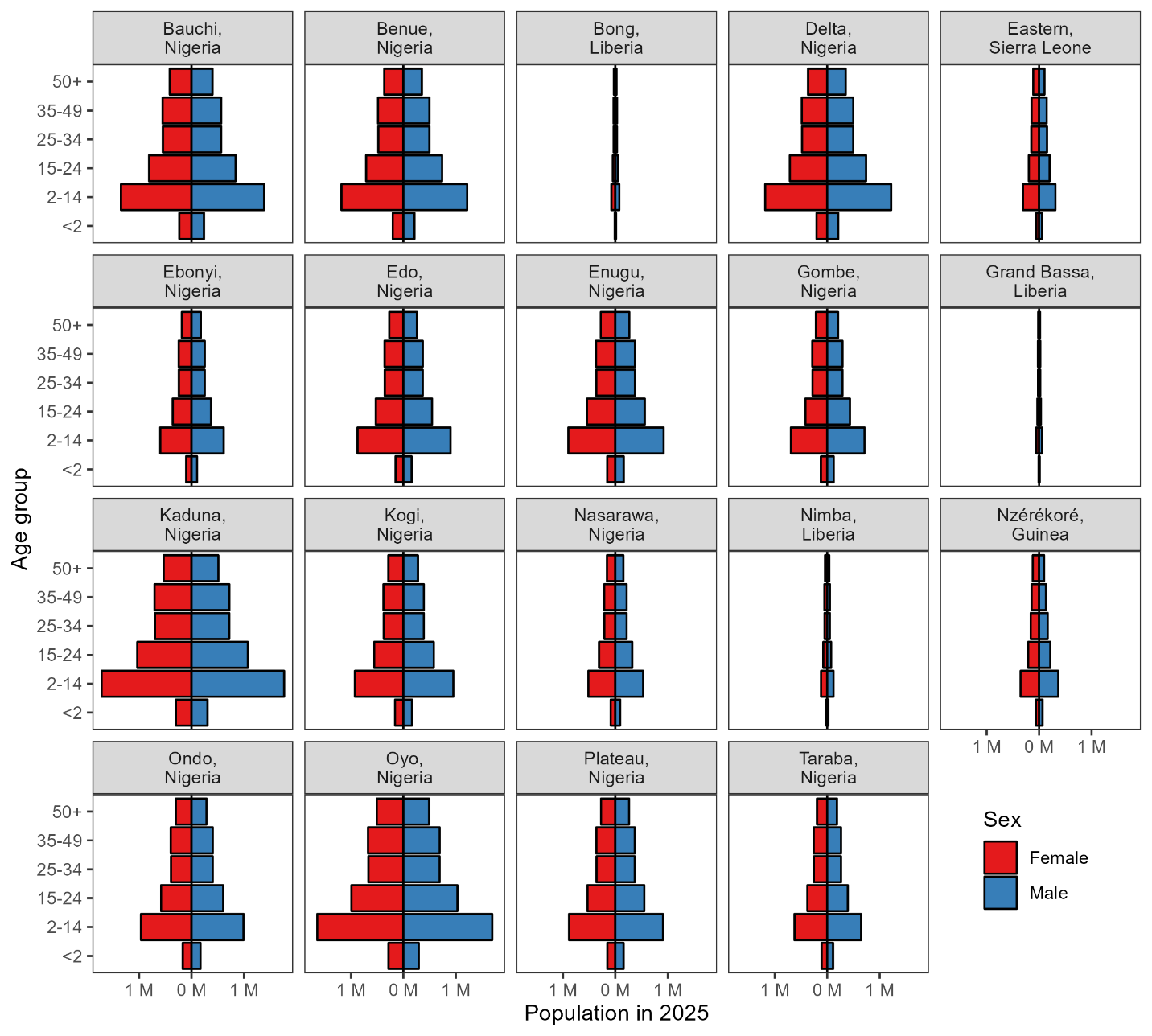

***Figure S1.*** *Population pyramids showing the projected number of males and females in each age group in 2025 across each of the included areas.*

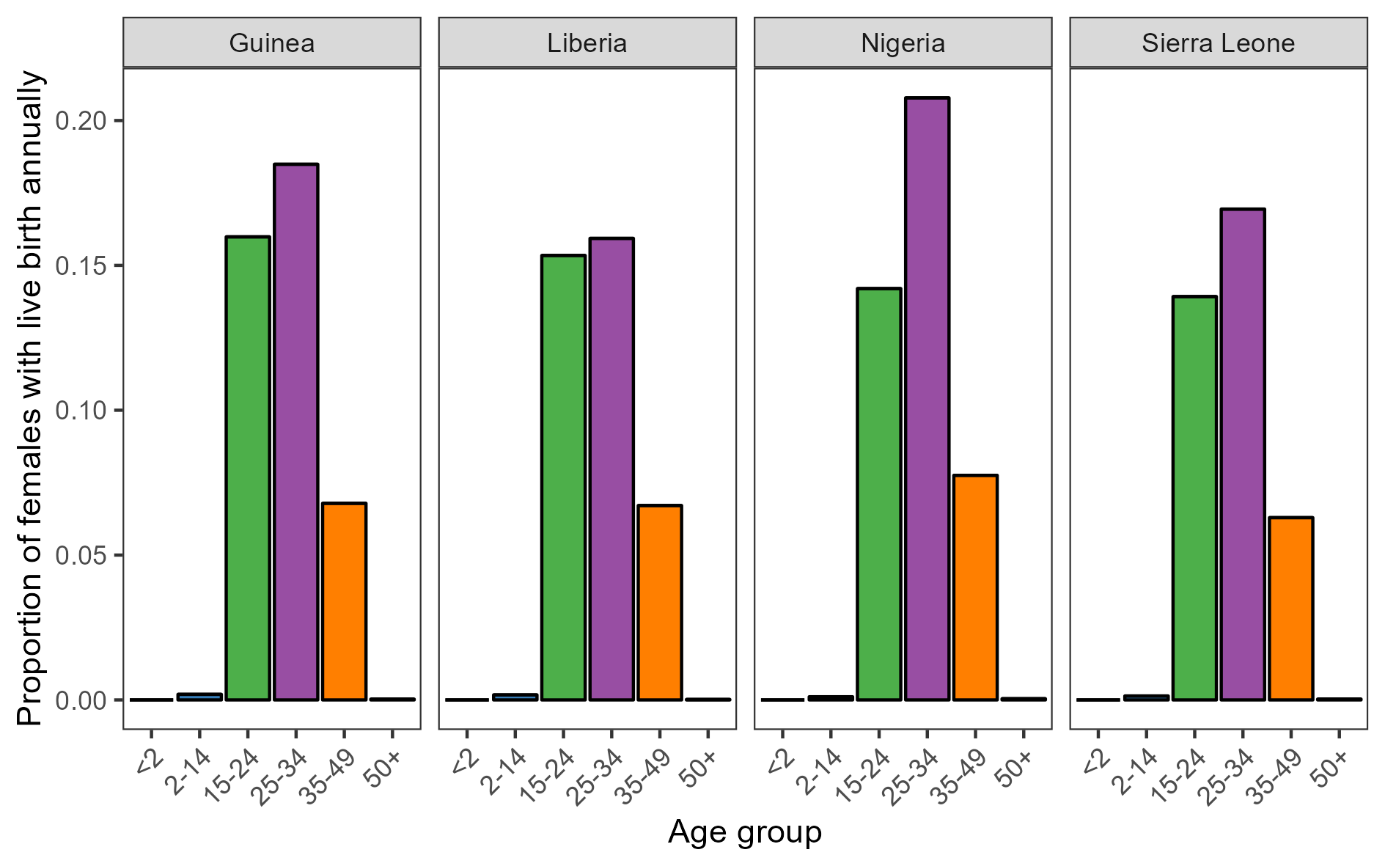

***Figure S2.*** *Estimates of the proportion of females in each age group in each country having a live birth annually.*

***1b. Age-specific infection risk***

Estimates of LASV zoonosis incidence used in this work are built upon a geospatial risk map developed by Basinski *et al*. that predicts zoonosis risk using *M. natalensis* occurrence data from 167 locations in 13 countries from 1977 to 2017, rodent LASV seroprevalence data from 13 studies in 6 countries from 1972 to 2014, human LASV seroprevalence data from 94 community-based serosurveys in 5 countries from 1970 to 2015, and rasterised estimates of human population size and 11 landcover features across continental West Africa.^9^ The Lassa fever model developed by Smith *et al.* used these rasterised spillover risk estimates to predict human LASV infection incidence at the pixel level.^1^ Resulting estimates of infection incidence and total infections in 2019 in the 19 areas included in the present study are shown in **Table S1**.

***Table S1****. Estimated annual zoonotic Lassa virus infections occurring in each area in 2019. These numbers correspond to estimates generated previously in Smith et al. (2024), for the areas highlighted as having endemic Lassa fever transmission in the 2024 WHO risk map.*

| **Country** | **Area** | **Infections per 100,000 population (2019)** | **Total infections (2019)** |
| --- | --- | --- | --- |
| Guinea | Nzérékoré | 695 (566 – 830) | 12,501 (10,184 – 14,941) |
| Liberia | Nimba | 536 (402 – 680) | 3,349 (2,511 – 4,249) |
|  | Bong | 575 (440 – 719) | 2,301 (1,760 – 2,879) |
|  | Grand Bassa | 501 (375 – 636) | 1,384 (1,036 – 1,757) |
| Nigeria | Delta | 623 (481 – 773) | 38,161 (29,484 – 47,405) |
|  | Ebonyi | 697 (566 – 834) | 21,520 (17,485 – 25,767) |
|  | Edo | 683 (540 – 834) | 30,894 (24,444 – 37,732) |
|  | Enugu | 676 (538 – 822) | 31,372 (24,964 – 38,148) |
|  | Gombe | 812 (668 – 963) | 29,092 (23,931 – 34,501) |
|  | Kaduna | 853 (702 – 1,012) | 75,833 (62,367 – 89,916) |
|  | Kogi | 635 (493 – 787) | 30,447 (23,611 – 37,721) |
|  | Nasarawa | 930 (766 – 1,102) | 24,800 (20,425 – 29,372) |
|  | Ondo | 601 (469 – 742) | 30,047 (23,450 – 37,058) |
|  | Oyo | 612 (471 – 762) | 52,141 (40,170 – 64,895) |
|  | Plateau | 846 (704 – 994) | 38,671 (32,195 – 45,435) |
|  | Taraba | 700 (560 – 848) | 22,712 (18,167 – 27,516) |
|  | Bauchi | 734 (590 – 887) | 51,121 (41,062 – 61,745) |
|  | Benue | 692 (558 – 834) | 42,343 (34,145 – 50,986) |
| Sierra Leone | Eastern | 726 (601 – 856) | 12,214 (10,121 – 14,406) |

Age-specific LASV seroprevalence estimates from the Enable study were used to estimate the age structure of infection incidence. Using Enable’s publicly available interim results,^10^ mean estimates and 95% confidence intervals (CI) of age-specific LASV seroprevalence pooled across the study’s seven sites (3 in Nigeria and 1 each in Benin, Guinea, Liberia and Sierra Leone) were digitised. These estimates range from 7.5% (95% CI: 5.7% – 9.5%) in children < 5 years old to 35.8% (95% CI: 33.8% – 37.9%) in those aged 50+. The sample size underlying seroprevalence in each age group was not reported, so these were estimated by assuming normally distributed confidence intervals and solving,

$$n_{a}=\frac{z^{2}\cdot p_{a}\cdot\left( 1-p_{a} \right)}{\left( p_{a}-l_{a} \right)^{2}}$$

where *p_a_* is the mean and *l_a_* is the lower bound of the CI for age group *a*, giving the squared margin of error in the denominator, and *z* is the z-value corresponding to the 95% CI (*z*=1.96). The resulting sample size pooled across all age groups (*n*=15,804) is within 4.8% of the reported number of individuals tested (*n*=16,604), noting that it is unclear whether all individuals tested are necessarily included in reported age-stratified pooled seroprevalence estimates (e.g., due to potential missing age or test result data).

A Bayesian serocatalytic model was fitted to these age-stratified pooled seroprevalence data to generate estimates of age-specific force of infection (FOI). As our study focuses on areas where Lassa fever is considered endemic, the model assumes that age-specific FOI is time-invariant, i.e. that there is a stable per-person risk of acquisition from the rodent host within each age group annually. Consistent with the underlying zoonosis risk map and overall analysis, there was assumed to be no LASV seroreversion, meaning that seroprevalence represents the proportion of individuals who have been infected with LASV by a certain age. The expected seroprevalence *S_a_* for an age group *a* is given by:

$$S_{a}=1 - e^{-\lambda_{a}\alpha_{a}}$$

where *λ_a_* denotes age-specfic FOI and *α_a_* denotes the midpoint age for age group *a*.

The model was implemented in R using the rstan package, which provides an interface to Stan modelling the likelihood of the seroprevalence data using a binomial distribution. Posterior distributions of age-specific FOI and the resulting seroprevalence for each group were obtained by running 4 Markov Chain Monte Carlo chains with 2,000 iterations each, discarding the first 1,000 iterations as burn-in (resulting in 4,000 posterior draws total, of which 500 were randomly sampled and carried forward in analysis). All *Rhat* values were close to 1, indicating good mixing and convergence.^11^ A summary of resulting FOI estimates is provided in **Table S2**.

The number of human LASV infections, *L*, can be estimated directly by multiplying age-specific FOI by the age-specific population at risk:

$$L_{d,a}=\lambda_{a}\times N_{d,a}\times\left( 1-S_{a} \right)$$

where *N_d,a_* is the total population size of each age group in each area. Applying *λ_a_* and *S_a_* estimates from Enable to the combined population size of all 19 areas in 2019 resulted in 1,042,989 (967,606 – 1,125,853) infections. However, Enable study sites were chosen because of a particularly high risk of LASV infection,^12^ which may lead to overestimation of incidence when aggregating over all 19 areas. To remain conservative, age-specific infections in each area, *L_d,a_*, were scaled to reproduce the total number of infections estimated previously in each area in 2019, thus conserving the area-level incidence estimated in Smith *et al.* (2024) while accounting for the relative age-specific risk of infection estimated from the age-stratified serocatalytic model. A summary of final age-specific incidence across all areas is provided in **Table S2**.

***Table S2.*** *Age-specific seroprevalence, estimated age-specific force of infection and total annual LASV spillover infections. Spillover infections were first calculated in the age groups reported in Enable then allocated to 1-year age bands, controlling for the number of individuals in each age band in each area, before regrouping to the final age groups considered in the model and summing across areas.*

| **Parameter (unit) *Symbol*** | **Age group** | **Mean (95% UI)** | **Source** |
| --- | --- | --- | --- |
| Seroprevalence (%)  $S_{a}$ | <5 | 7.5 (5.7 – 9.5) | ^10^ |
|  | 5-17 | 18.0 (17.0 – 19.0) |  |
|  | 18-24 | 26.8 (25.1 – 28.7) |  |
|  | 25-34 | 28.1 (26.2 – 30.0) |  |
|  | 35-49 | 34.3 (32.5 – 36.3) |  |
|  | 50+ | 35.8 (33.8 – 37.9) |  |
| Force of infection (%)  $\lambda_{a}$ | <5 | 3.14 (2.45 – 3.91) | Estimated |
|  | 5-17 | 1.80 (1.69 – 1.92) |  |
|  | 18-24 | 1.49 (1.37 – 1.61) |  |
|  | 25-34 | 1.12 (1.03 – 1.21) |  |
|  | 35-49 | 1.00 (0.93 – 1.07) |  |
|  | 50+ | 0.68 (0.63 – 0.73) |  |
| Annual LASV spillovers (n, 2019)  $L_{a}$ | <2 | 76.6 K (65.1 K – 88.7 K) | Estimated |
|  | 2-14 | 276.3 K (259.7 K – 292.7 K) |  |
|  | 15-24 | 97.8 K (88.7 K – 107.4 K) |  |
|  | 25-34 | 45.2 K (40.5 K – 50.2 K) |  |
|  | 35-49 | 37.0 K (33.5 K – 40.9 K) |  |
|  | 50+ | 17.7 K (15.9 K – 19.6 K) |  |

***1c. Projecting infections through time***

National case data from the NCDC were used to distribute LASV infections seasonally. Weekly counts of the number of laboratory-confirmed Lassa fever cases occurring each year from 2019 to 2023 were digitised from the Lassa Fever Situation Report corresponding to Epi Week 25 of 2024.^13^ For each year, the proportion of cases occurring in each epidemiological week was calculated and a rolling mean (k=7) was applied to define average seasonality (**Figure S3**). To estimate infection timing from reported cases, the rolling average was shifted by 3 weeks, the approximate sum of mean estimates of the incubation period (10.3 days) and the delay from symptom onset to hospital admission (9.3 days).^1,14^ From the 3-week shifted rolling mean, the proportions of cases occurring in each epidemiological week were applied to annual infection projections to simulate weekly infection incidence.

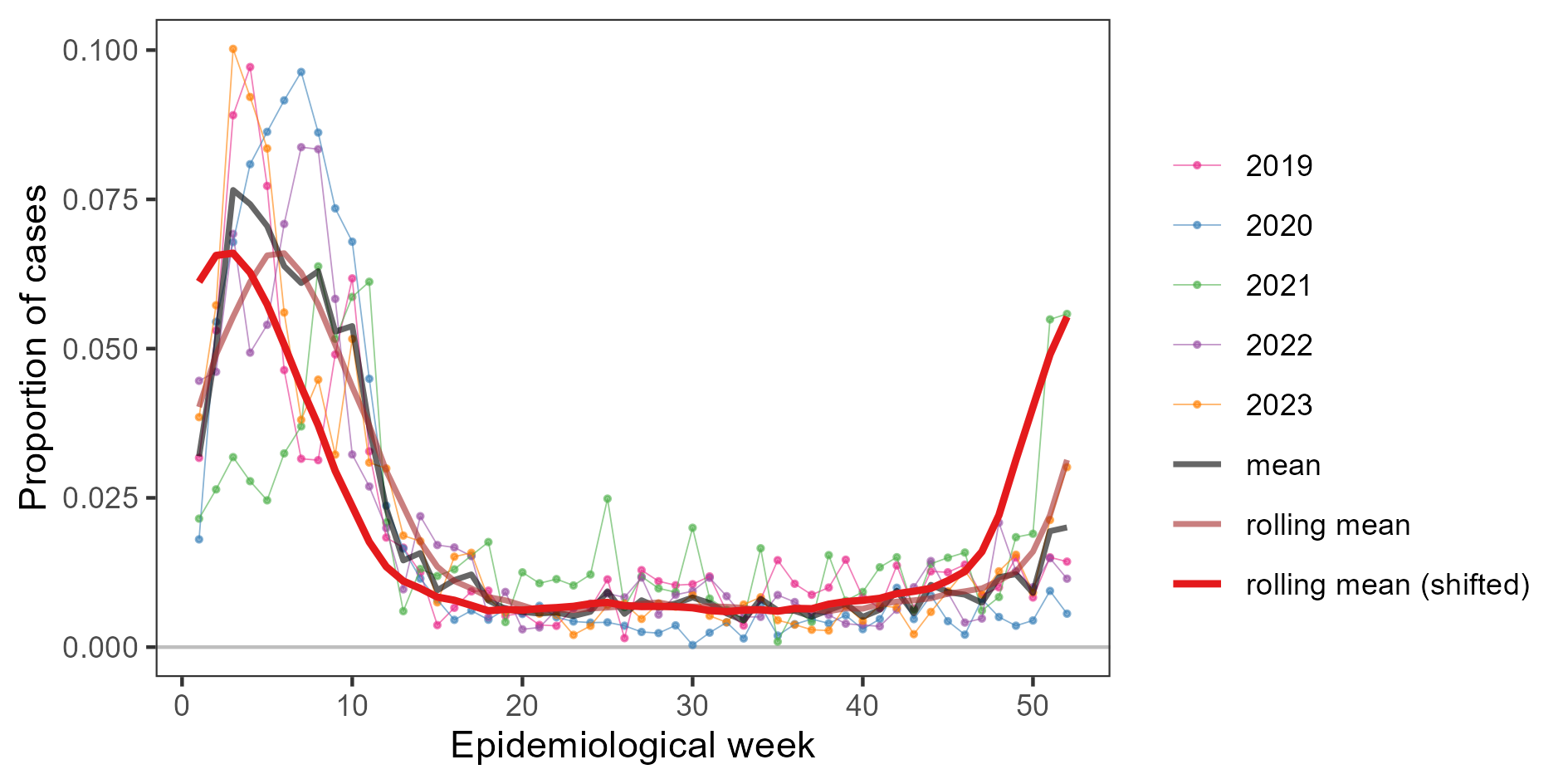

***Figure S3****. The proportion of Lassa fever cases occurring in each epidemiological week from 2019 to 2023. The shifted rolling mean represents estimated infection timing and was calculated assuming a 3-week delay from infection acquisition to case reporting.*

***1d. Symptomatic Lassa fever risks stratified by age, sex and pregnancy status***

In the base case analysis, the probability of infected individuals developing any symptoms, ℙ*(symptoms|infection)*, was assumed to be constant across age groups (**Figure S4 panel A**). However, a sensitivity analysis was included whereby infection-symptom risk increases across the six included age groups while maintaining the total number of symptomatic infections, *N^symptoms^*, from the population-wide estimate (**Figure S4 panel B**). This was calculated by scaling age-specific risk relative to baseline risk by a coefficient *ρ_a_* and minimising an objective function over each matched parameter draw by solving for the coefficient *φ*,

$$\varphi^{*}=\arg\min_{\varphi} \left( N^{symptoms}-\sum_{a} L_{a}\mathbb{\cdot P}\left( symptoms | infection \right)\cdot\rho_{a}\cdot\varphi\right), \rho_{a}=0.5,1,\ldots,3$$

where *ρ_a_* was increased by increments of 0.5 across the six included age groups, from 0.5 in children <2 to 3 in adults 50+, giving final age-specific estimates,

$$\mathbb{P}_{a}\left( symptoms | infection \right)\mathbb{=P(}symptoms|infection)\cdot\rho_{a}\cdot\varphi^{*}$$

In one simulation (0.2%), symptom risk in adults aged 50+ was >1 so was truncated to equal exactly 1.

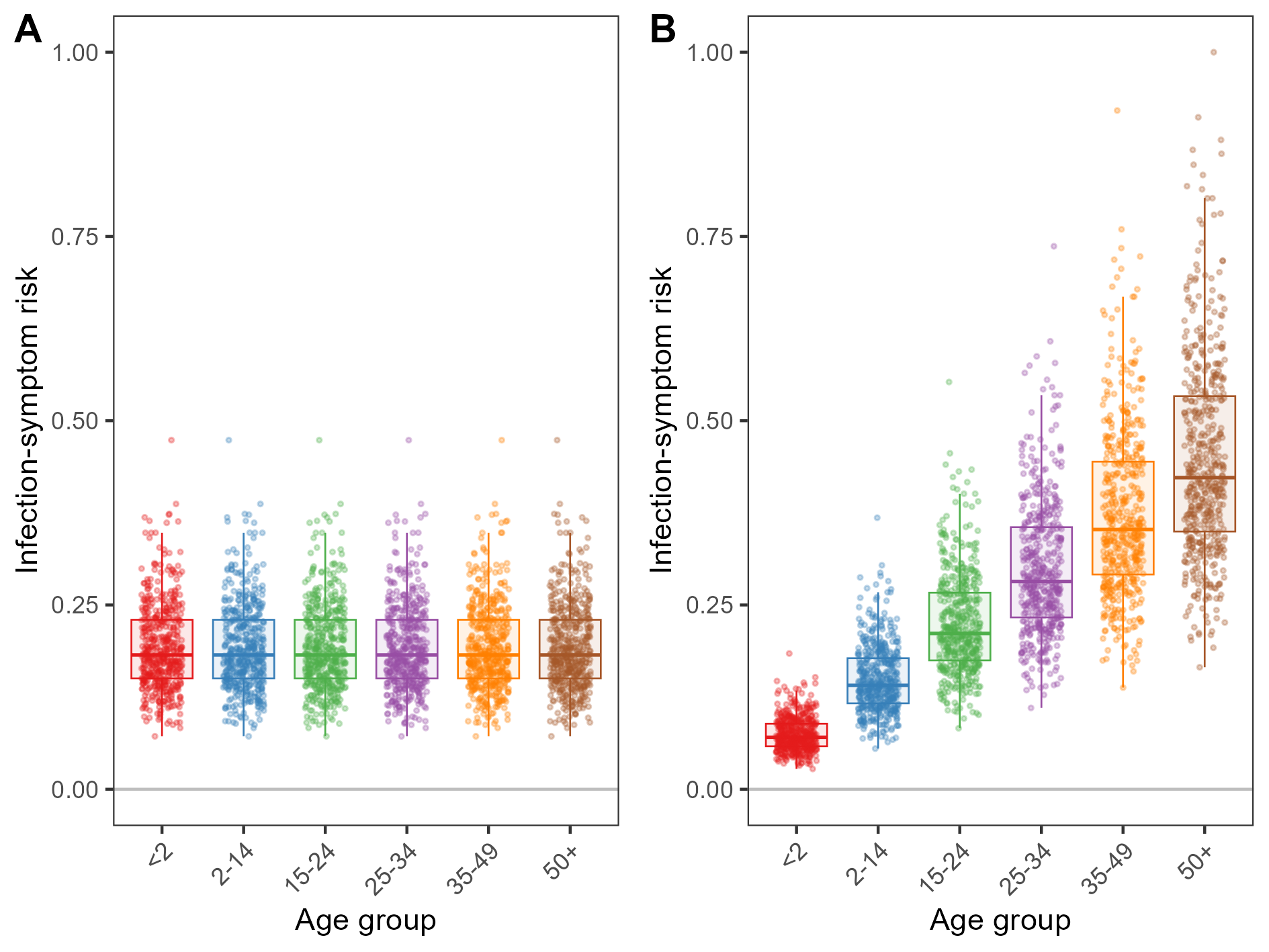

***Figure S4****. Infection-symptom risk model input parameters. (****A****) Each point is a draw (n=500) from an inverse-logit transformed Normal distribution from an inverse variance meta-analysis and generalised linear model fit to data describing the proportion of LASV infections associated with illness in a prospective cohort study in four villages in Sierra Leone.^15^ In our baseline analysis, infection-symptom risk is assumed to be identical across age groups. (****B****) Each point is a draw (n=500) of the data from panel A after rescaling by ρ_a_ in a sensitivity analysis assuming increasing symptom risk with age. Boxplot whiskers extend to the most extreme value no further than 1.5 × IQR from the nearest hinge.*

We previously estimated the infection-hospitalisation risk (IHR) for human LASV infection to be 0.86% (95% CI: 0.56%–1.12%).^1,16^ This estimate was generated by dividing the annual number of laboratory-confirmed hospitalised Lassa fever cases in the high surveillance states of Edo and Ondo between 2018 and 2021 by the annual number of infections occurring in those states in 2019, as estimated by our geospatial risk map. To estimate age-specific IHRs, 500 draws of total zoonotic infections in Nigeria were matched to 500 draws of the overall IHR and multiplied to generate a distribution of the predicted total number of hospitalisations occurring in Nigeria. The number of NCDC-reported laboratory-confirmed hospitalised Lassa fever cases occurring in each age-sex group in Nigeria from 2019 to 2023 were then extracted and summed.^13^ These data cover all states of Nigeria and were assumed to be representative of the age-sex structure of all hospitalised cases nationally. The proportions of all hospital cases occurring in each age-sex group were then fit to a Dirichlet distribution. A total of 500 stochastic draws from this distribution (**Figure S5 panel A**) were multiplied by the distribution of predicted total hospitalisations to stratify predicted hospitalisations into corresponding age-sex groups. Finally, this estimate was divided by the projected number of infections in each age-sex group in Nigeria to recover age- and sex-specific IHR (**Figure S5 panel B**).

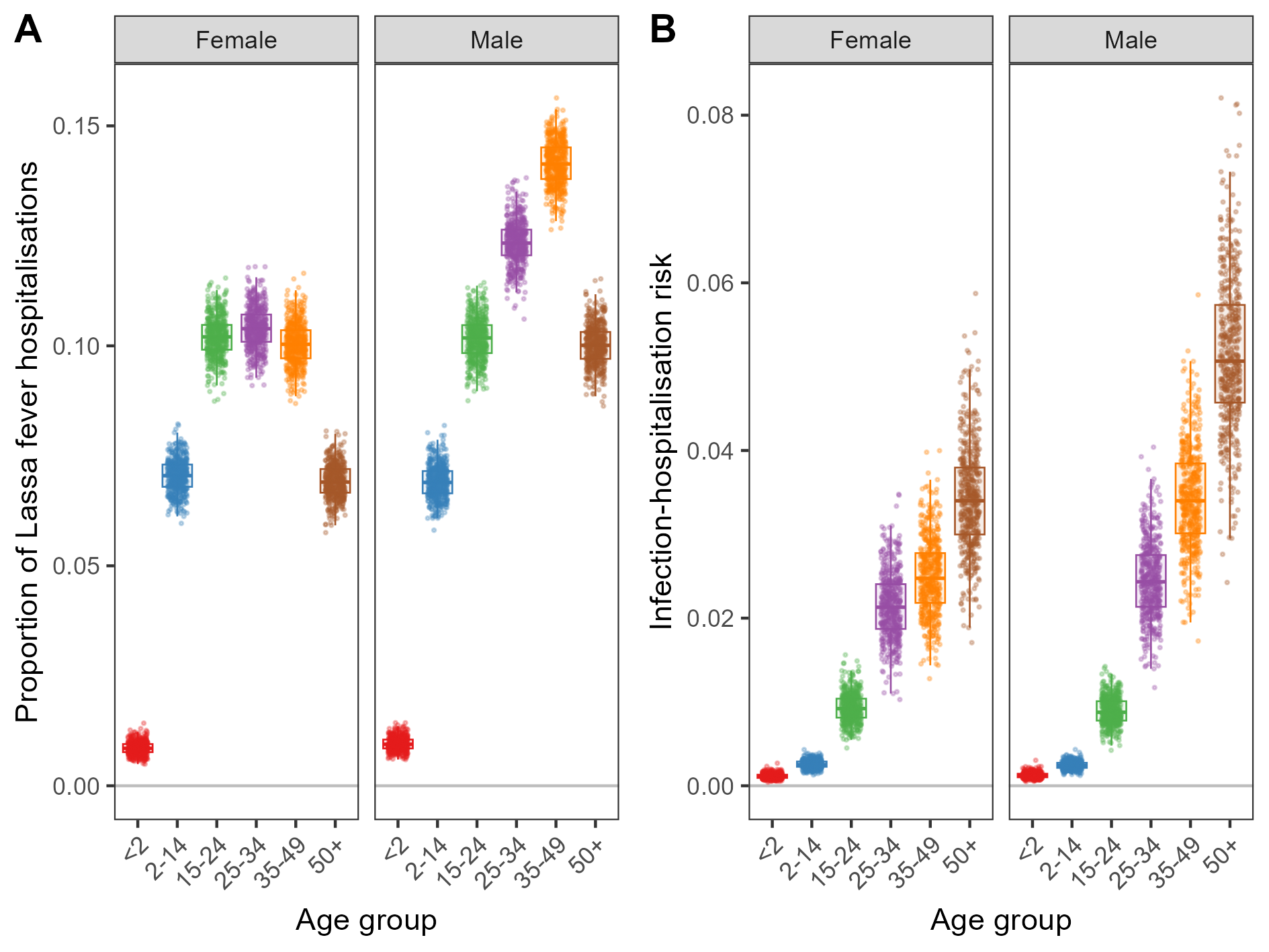

***Figure S5****. Hospitalisation risk model input parameters. (****A****) Each point is a draw (n=500) from a Dirichlet distribution fit to the proportion of all Lassa fever hospitalisations reported by NCDC from 2019 to 2023 occurring in each age-sex group,^13^ after age group transformation. (****B****) Each point is a draw (n=500) of the estimated proportion of infections that result in severe disease and hospitalisation in each age-sex group. Boxplot whiskers extend to the most extreme value no further than 1.5 × IQR from the nearest hinge.*

Data from the LASCOPE study were used to estimate age-specific case-fatality risk (CFR) among hospitalised Lassa fever patients. The LASCOPE study included patients without age restrictions with laboratory-confirmed Lassa fever presenting to the Lassa fever ward of Owo Medical Centre between April 2018 and March 2020.^14^ A follow-up paediatric study including all patients <15 years old presenting to the same ward between April 2018 and February 2023 has since been published.^17^ To increase sample size and improve estimates of mortality risk in children for patients aged <12 years old, data from the original study were replaced with data from the follow-up paediatric study. The resulting data set includes data on 87 children (2 deaths) aged 1 month to 11 years and 460 individuals aged 12+ (59 deaths). Neonates aged <1 month (n=8) were excluded, as risk of neonatal death was estimated separately using results from a systematic review and meta-analysis (see below). Duvignaud *et al.* (2021) reported a greater mortality risk in those aged ≥45 than those <45 but found no association with sex.^14^ Therefore, males and females were pooled together and age-specific CFRs were calculated as the number of deaths in each age group divided by the number of cases. Given the relatively low sample size, patients and deaths in each age group were bootstrap resampled 500 times to generate a distribution of CFR estimates in each age group (**Figure S6**).

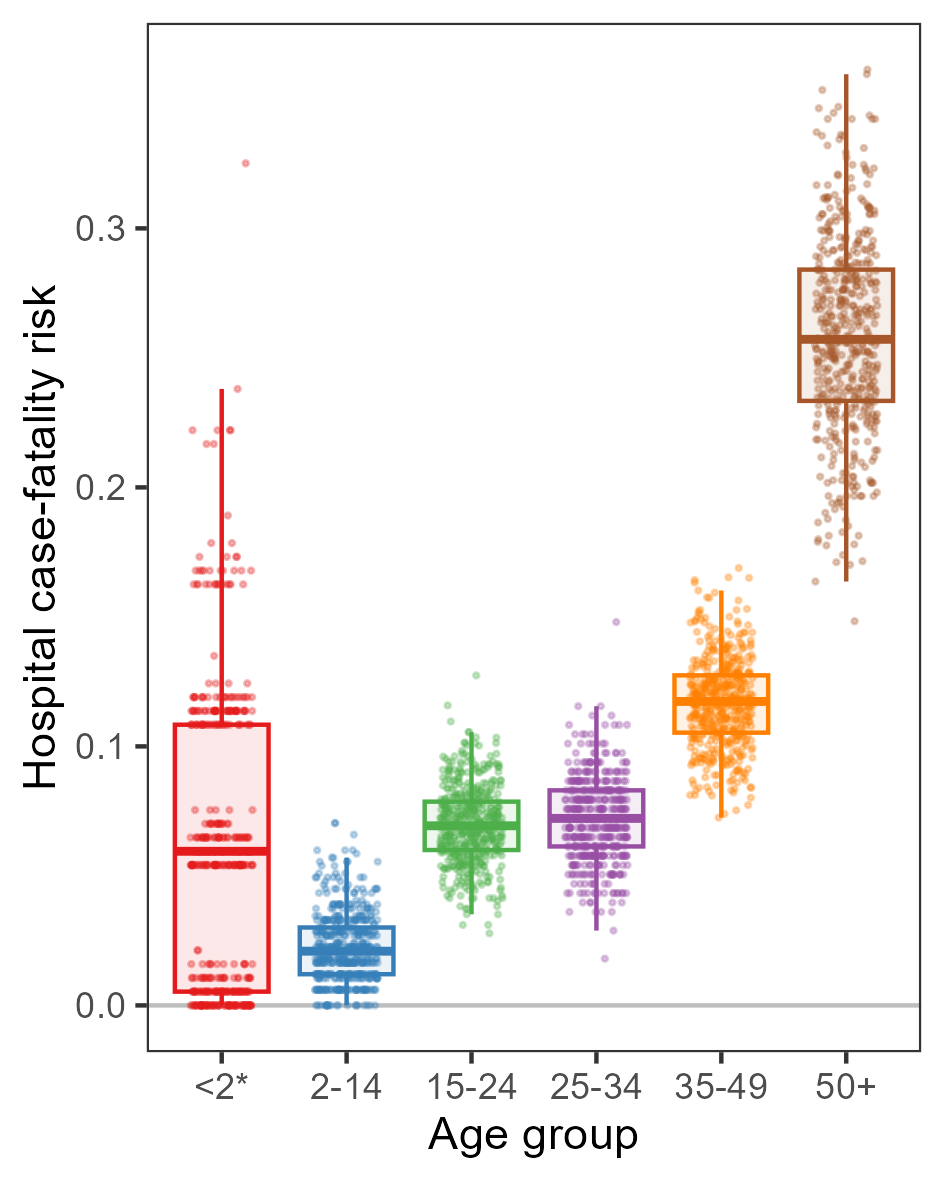

***Figure S6****. The estimated proportion of hospitalised cases that result in death, stratified by age group. Each point is a draw (n=500) from bootstrapped cases and deaths reported in the LASCOPE studies.^14,17^ The age group <2 excludes children <1 month, as neonatal Lassa fever deaths were quantified using data from a systematic review and meta-analysis of neonatal risk. Boxplot whiskers extend to the most extreme value no further than 1.5 × IQR from the nearest hinge.*

Pregnant women with Lassa fever are known to be at increased risk of death relative to non-pregnant women and at high risk of foetal and neonatal demise. Data from a systematic review and meta-analysis from Kayem *et al.* (2020) were used to quantify these risks, including an estimated odds ratio for death in pregnant women relative to non-pregnant women of *OR_P_* = 2.86 (95% CI: 1.77 – 4.63).^18^ This odds ratio was used to estimate age-specific CFR in pregnant women, *CFR_P,a_*, relative to the baseline risk in men and women, *CFR_a_*, as,

$${CFR}_{P,a}=\frac{\left( \frac{{CFR}_{a}}{1-{CFR}_{a}}\cdot OR_{p} \right)}{1+\left( \frac{{CFR}_{a}}{1-{CFR}_{a}}\cdot OR_{p} \right)}$$

By increasing mortality risk among pregnant women, the overall mortality risk among women exceeds that of men. To counteract this and maintain equal overall mortality risk in hospitalised men and women, as reported in the LASCOPE study,^14^ the overall age-specific CFR was defined as the age-specific probability of death given hospitalisation:

$$\mathbb{P}_{a}\left( death | hospital \right)={CFR}_{a}$$

and the CFR in pregnant women as the age-specific probability of death given being pregnant and in hospital, assuming that pregnancy lasts 40 weeks and is distributed uniformly throughout the year:

$$\mathbb{P}_{a}\left( death | pregnant, hospital \right)={CFR}_{P,a}$$

The CFR in non-pregnant women, *CFR_F,a_*, was then derived following probability laws:

$$\mathbb{P}_{a}\left( pregnant | hospital \right)=\mathbb{P}_{a}\left( livebirth in the year \right)\cdot\frac{40}{52}$$

$$\mathbb{P}_{a}\left( not pregnant | hospital \right)=1-\mathbb{P}_{a}\left( pregnant|hospital \right)$$

$$\mathbb{P}_{a}\left( pregnant | death \right)=\mathbb{P}_{a}\left( death|pregnant,hospital \right)\mathbb{\cdot P}_{a}\left( pregnant|hospital \right)$$

$$\mathbb{P}_{a}\left( not pregnant | death \right)=\mathbb{P}_{a}\left( death|hospital \right)\mathbb{-P}_{a}\left( pregnant|death \right)$$

giving

$${CFR}_{F,a}=\frac{\mathbb{P}_{a}\left( not pregnant|death \right)}{\mathbb{P}_{a}\left( pregnant|death \right)}$$

This resulted in CFR estimates in non-pregnant women that are slightly lower than in men of the same age, with a greater effect in female age groups with a greater share of the hospitalised population being pregnant (**Figure S7**). Nigerian data were used for ℙ*_a_(livebirth in the year)* because Nigerian data were also used to estimate baseline values of *CFR_a_*.

For simplicity, it was assumed that pregnant women are not at increased risk of severe disease and hospitalisation but rather only at increased risk of severe outcomes once hospitalised. This assumption has face validity, as the mean share of WCBA with Lassa fever that are pregnant upon hospitalisation in the model (11%) is consistent with the share reported in LASCOPE (10%).^14^

A final summary of acute Lassa fever disease risks stratified by age and sex is provided in **Table S3**.

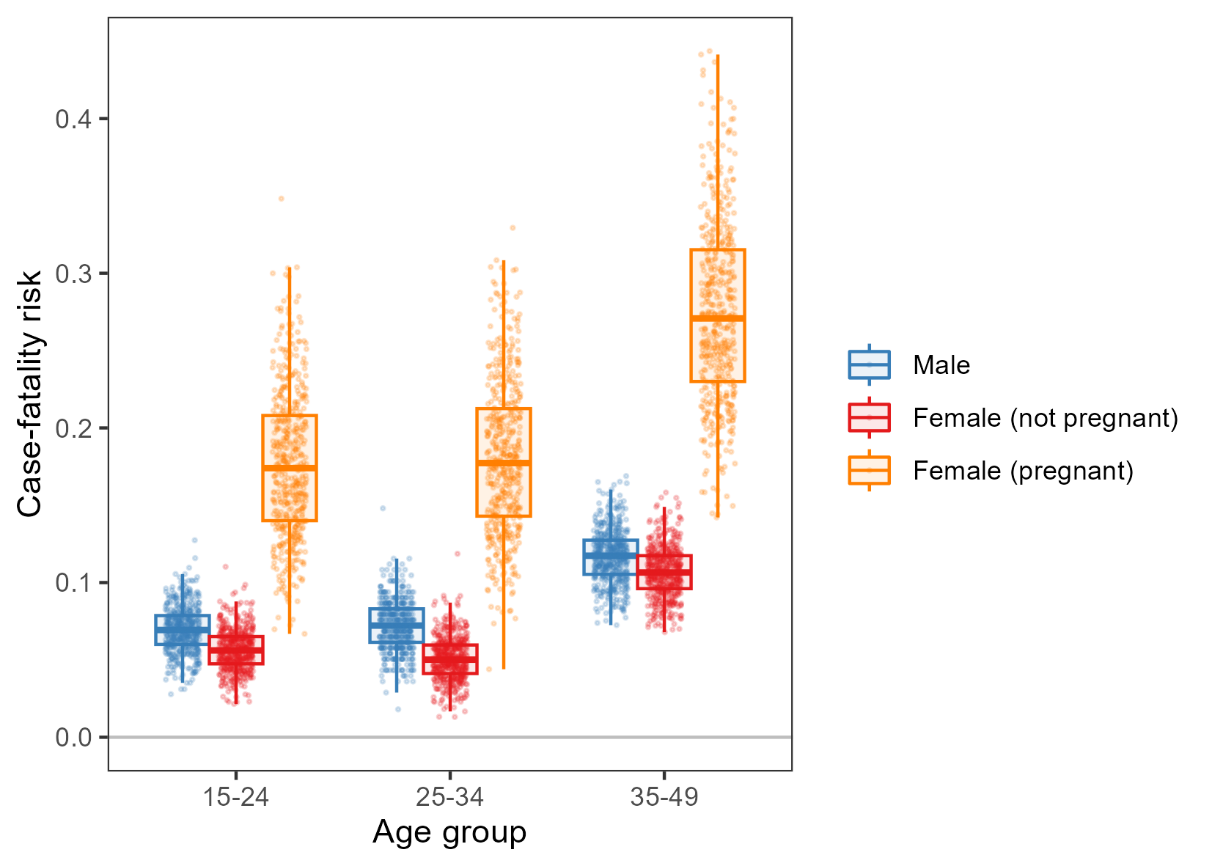

***Figure S7.*** *The estimated proportion of hospitalised cases that result in death in men and women, after adjustment to account for increased odds of mortality in pregnant women relative to non-pregnant women, but maintaining equal overall risk in men and women of the same age group. Each point is a draw (n=500) from bootstrapped cases and deaths reported in the LASCOPE study.^14^ Boxplot whiskers extend to the most extreme value no further than 1.5 × IQR from the nearest hinge.*

***Table S3.*** *Final age- and sex-stratified risks of Lassa fever. For symptom risk, stratification by age is only considered in sensitivity analysis; while the relative difference in risk across age groups is assumed, these values nonetheless recapitulate the population-wide estimate from Smith et al. (2024).^1^ For case-fatality risk, which applies to hospitalised cases, we distinguish between non-pregnant female (F) and pregnant female (P), and the age group <2 includes only those aged 1 month to 23 months, as neonatal mortality risk is estimated separately.*

| **Parameter (unit) *Symbol*** | **Sex** | **Age group** | **Mean (95% UI)** | **Source** |
| --- | --- | --- | --- | --- |
| Infection-symptom risk (%)  $\mathbb{P}_{a}\left( symptoms \right\vert infection)$ | / | / | 19.3 (10.2 – 32.7) | ^1^ |
|  |  | <2 | 7.5 (3.9 – 12.9) | Assumed* |
|  |  | 2-14 | 14.9 (7.8 – 25.9) |  |
|  |  | 15-24 | 22.4 (11.7 – 38.8) |  |
|  |  | 25-34 | 29.9 (15.7 – 51.7) |  |
|  |  | 35-49 | 37.3 (19.6 – 64.7) |  |
|  |  | 50+ | 44.8 (23.5 – 77.6) |  |
| Infection-hospitalisation risk (%)  $\mathbb{P}_{s,a}\left( hospital \right\vert infection)$ | M | <2 | 0.12 (0.07 – 0.20) | Estimated |
|  |  | 2-14 | 0.24 (0.16 – 0.34) |  |
|  |  | 15-24 | 0.89 (0.60 – 1.22) |  |
|  |  | 25-34 | 2.46 (1.64 – 3.39) |  |
|  |  | 35-49 | 3.44 (2.28 – 4.67) |  |
|  |  | 50+ | 5.16 (3.46 – 7.10) |  |
|  | F | <2 | 0.12 (0.06 – 0.18) |  |
|  |  | 2-14 | 0.26 (0.17 – 0.35) |  |
|  |  | 15-24 | 0.93 (0.63 – 1.32) |  |
|  |  | 25-34 | 2.15 (1.47 – 2.93) |  |
|  |  | 35-49 | 2.50 (1.69 – 3.38) |  |
|  |  | 50+ | 3.43 (2.25 – 4.85) |  |
| Case-fatality risk (%)  $\mathbb{P}_{s,a}\left( death \right\vert hospital)$ | M | <2* | 6.1 (0.0 – 17.3) | Estimated |
|  |  | 2-14 | 2.2 (0.0 – 5.5) |  |
|  |  | 15-24 | 7.0 (4.1 – 9.9) |  |
|  |  | 25-34 | 7.1 (4.3 – 10.5) |  |
|  |  | 35-49 | 11.7 (8.4 – 15.2) |  |
|  |  | 50+ | 25.9 (18.4 – 33.6) |  |
|  | F | <2* | 6.1 (0.0 – 17.3) |  |
|  |  | 2-14 | 2.2 (0.0 – 5.5) |  |
|  |  | 15-24 | 5.6 (3.2 – 8.4) |  |
|  |  | 25-34 | 5.1 (2.5 – 7.7) |  |
|  |  | 35-49 | 10.7 (7.7 – 14.1) |  |
|  |  | 50+ | 25.9 (18.4 – 33.6) |  |
|  | P | <2* | / |  |
|  |  | 2-14 | 6.1 (0.0 – 15.3) |  |
|  |  | 15-24 | 17.6 (9.6 – 27.6) |  |
|  |  | 25-34 | 18.0 (9.7 – 28.6) |  |
|  |  | 35-49 | 27.4 (17.3 – 39.8) |  |
|  |  | 50+ | / |  |

The meta-analyses of Kayem *et al.* quantifying risks of Lassa fever-associated foetal loss and neonatal loss were updated to include data from LASCOPE. The initial full-population results from LASCOPE reported 14 documented pregnancy outcomes, among which there were 6 spontaneous miscarriages and 1 intrauterine death.^14^ In the follow-up paediatric study, among 7 neonatal cases of Lassa fever there were 3 deaths.^17^ It was implicitly assumed that these neonatal cases were associated with maternal Lassa fever, though maternal infection history/status was not reported. The other included studies are described in Kayem *et al*.^18^

Proportional meta-analyses implementing the Freeman-Tukey double arcsine transformation were conducted, as this method is well suited to binomial data with extreme proportions. A random effects model was used to calculate a weighted summary estimate and the 95% CI for the proportion of hospitalised maternal Lassa fever cases resulting in foetal loss (**Figure S8**) or neonatal death (**Figure S9**).

To estimate the proportion of foetal losses among hospitalised pregnant Lassa fever patients that are attributable to Lassa fever and not other causes, random draws of the baseline risk of stillbirth in West Africa estimated in 2019 were subtracted from weighted summary estimates from the random effects meta-analysis,^19^ for a final risk estimate of 58.0% (95% UI: 28.4% – 80.9%). To estimate the proportion of neonatal deaths among pregnant Lassa fever patients that are attributable to Lassa fever and not other causes, random draws of the baseline risk of neonatal death estimated in Nigeria in 2020 were subtracted from weighted summary estimates from the random effects meta-analysis,^20^ for a final risk estimate of 33.1% (95% UI: 15.8% – 52.3%).

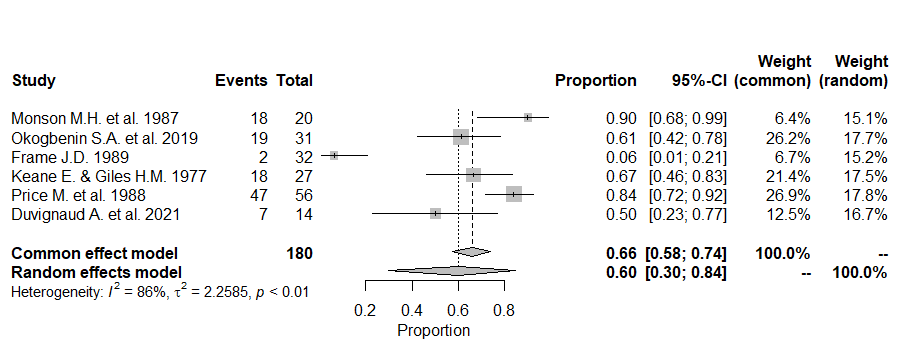

***Figure S8.*** *Proportional meta-analysis of studies reporting foetal loss from Lassa fever in pregnancy.* $I^{2}$*, Higgins statistics;* $\tau^{2}$*, tau squared; p, p-value associated with Cochran’s Q for heterogeneity; Events, number of foetal losses; Total, number of foetuses included in the analysis.*

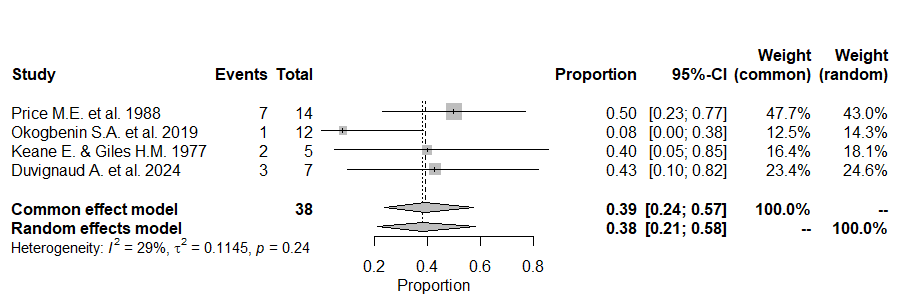

***Figure S9.*** *Proportional meta-analysis of studies reporting neonatal death following maternal Lassa fever.* $I^{2}$*, Higgins statistics;* $\tau^{2}$*, tau squared; p, p-value associated with Cochran’s Q for heterogeneity; Events, number of neonatal deaths; Total, number of neonates included in the analysis.*

Neither neonatal deaths nor foetal losses were counted as Lassa fever cases or hospitalisations, as neonatal deaths attributed to maternal Lassa fever do not necessarily result from infection (all live births in the LASCOPE data were tested and found to be Lassa fever negative).^14^ Further, no additional care costs were included due to many neonatal deaths occurring within a few hours of birth and many not presenting to hospital.^21,22^

***1e. Lassa fever-induced sensorineural hearing loss***

To characterise Lassa fever-induced sensorineural hearing loss (SNHL), 3 relevant systematic literature reviews were consulted and 20 unique research articles describing hearing loss subsequent to Lassa fever were identified.^23-25^ Most of these were case reports or retrospective observational studies. One case-control study by Ficenec *et al.* (2020) quantified risk of hearing loss in Lassa fever survivors (17%) relative to community-matched controls (1%) but did not conduct follow-up or provide individual patient-level audiometry data.^26^ Only one study, a prospective audiometric evaluation conducted by Cummins *et al.* (1990) in Sierra Leone, provides detailed individual patient-level audiometry data at baseline and 1-year follow-up, allowing for quantification of Lassa fever-induced SNHL risk, duration and disability.^27^ They reported SNHL in 14 (29%) of 48 confirmed Lassa fever cases and in 0 of 20 febrile controls, found no obvious association between recovery and severity of the initial hearing deficit, and observed a high pooled prevalence (17.6%) among local villagers and healthcare workers known to be LASV seropositive. These findings suggest that post-acute SNHL is not limited to severe cases or strongly associated with age, so SNHL risk was applied to anyone surviving symptomatic disease, but in sensitivity analysis SNHL risk was limited only to those surviving hospitalisation.

Although some Lassa fever survivors develop lifelong hearing loss, many recover over the months and years following their infection.^23^ To quantify the average duration of SNHL, mean auditory threshold (MAT) estimates were extracted for all patients in both ears at baseline and at 1-year follow-up. Three patients lacking follow-up data were excluded, and only data from each patient’s better ear at both time points were carried forward for analysis (**Figure S10 panel A**), as the Global Burden of Disease (GBD) study on hearing loss defines hearing loss according to the quietest sound an individual can hear in their better ear (GBD Hearing Loss Collaborators, 2021).^28^ These patients were bootstrap resampled 10,000 times and, for each sample, mean MAT in decibels (dB) was calculated at baseline, *M*, and at follow-up, *m*, at *t*=1 year. Assuming an exponential decay in deafness severity (MAT), the MAT decay rate, *d*, was calculated from,

$$m=M e^{-d\cdot t}$$

solving for *d* in each sample:

$d=-\frac{\log\left( \frac{m}{M} \right)}{\left( t-t_{0} \right)}$

resulting in a distribution of 10,000 exponential decay curves corresponding to each bootstrapped sample of patient audiometry data (**Figure S10, Panel B**).

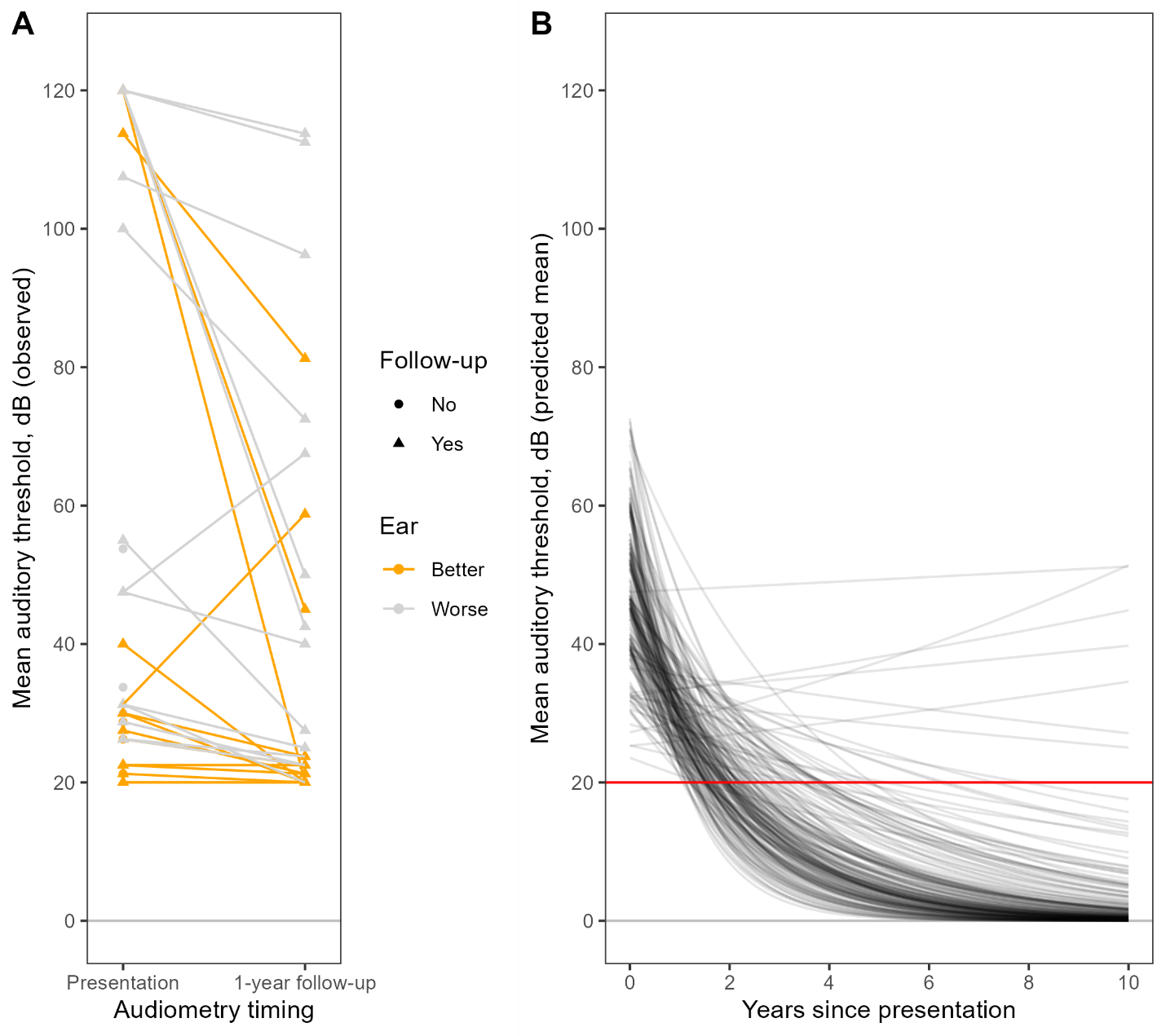
***Figure S10****. Resolution of Lassa fever-induced hearing loss. (****A****) Patient audiometry data from Cummins et al. (1990),^27^ highlighting in gold the data carried forward in analysis: measures of MAT in dB at presentation and 1-year follow-up in patients’ better ear at each time point (not necessarily the same ear in the case of asymmetric improvement in bilateral SNHL). (****B****) Modelled average trajectories of SNHL recovery over time, estimated by fitting bootstrap resampled audiometry data to exponential decay curves. The red horizontal line indicates the threshold at which patients are defined as having any deafness according to the GBD study on hearing loss.*

Mean SNHL duration was estimated by calculating when each decay curve crossed 20 dB, which in the GBD study is the minimum threshold at which patients are classified as having normal hearing and hence no hearing-related disability:

$${dur}^{SNHL}=-\frac{\log\left( \frac{20}{M} \right)}{d}$$

This resulted in a final distribution of SNHL durations with a mean of 2.1 years (**Figure S11**).

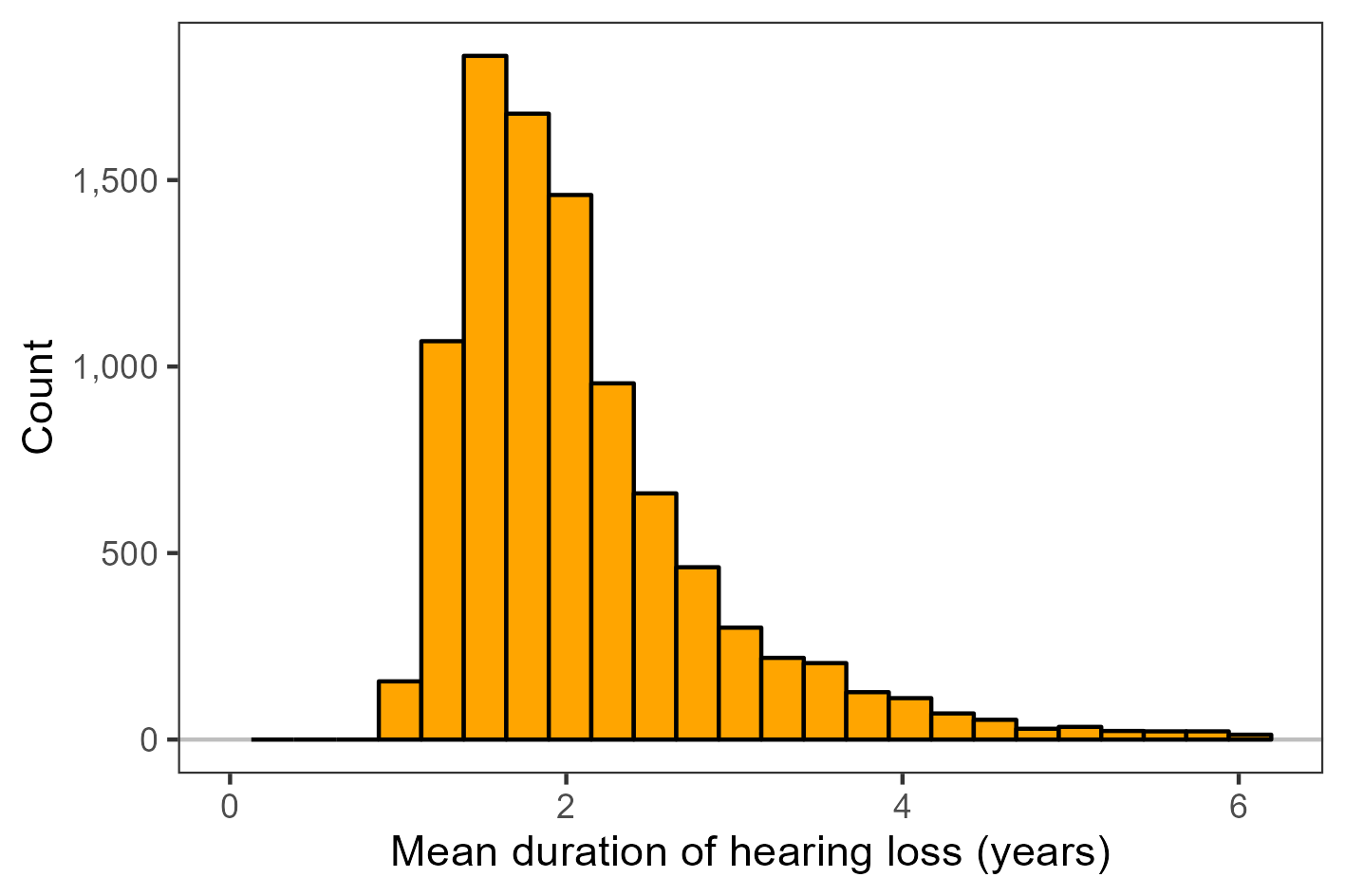

***Figure S11****. Hearing loss duration. Histogram showing the 95% probability distribution (trimmed at 2.5% and 97.5%) of the mean duration of Lassa fever-induced SNHL, estimated from bootstrap resampled audiometry data.*

To quantify disability associated with SNHL, mean MAT measures were taken at baseline from the bootstrapped sample and the severity of average hearing loss was classified according to GBD definitions (mild, 20 dB ≤ MAT < 35 dB; moderate, 35 dB ≤ MAT < 50 dB; moderately severe, 50 dB ≤ MAT < 65 dB; severe, 65 dB ≤ MAT < 80 dB; profound, 80 dB ≤ MAT < 95 dB; and complete, MAT ≥ 95 dB).^28^ Based on these classifications, for each baseline MAT value, a disability weight associated with the corresponding level of hearing loss severity was stochastically drawn, ranging from a mean of 0.01 in mild hearing loss with ringing to 0.316 given complete hearing loss with ringing and assuming normally distributed confidence intervals. The GBD study also reports estimates of hearing loss disability weights without ringing.^28^ However, since Lassa fever-induced SNHL is commonly associated with tinnitus, and since tinnitus is itself a common sequela of Lassa fever even in the absence of SNHL,^25,26^ disability weights associated with ringing were selected.

The binning of disability estimates according to severity categories results in stepwise increases in hearing loss-associated disability with increasing baseline MAT. To generate a more plausible distribution, these data were fitted to a Gamma-distributed generalised additive model (GAM) with a log link function and a smooth term with k=3 basis dimensions (**Figure S12 panel A**). Finally, using the distribution of bootstrap resampled mean MAT from Cummins *et al.* (1990) at baseline as input data, this GAM was used to generate a final distribution of disability values associated with Lassa fever-induced SNHL (**Figure S12 panel B**).

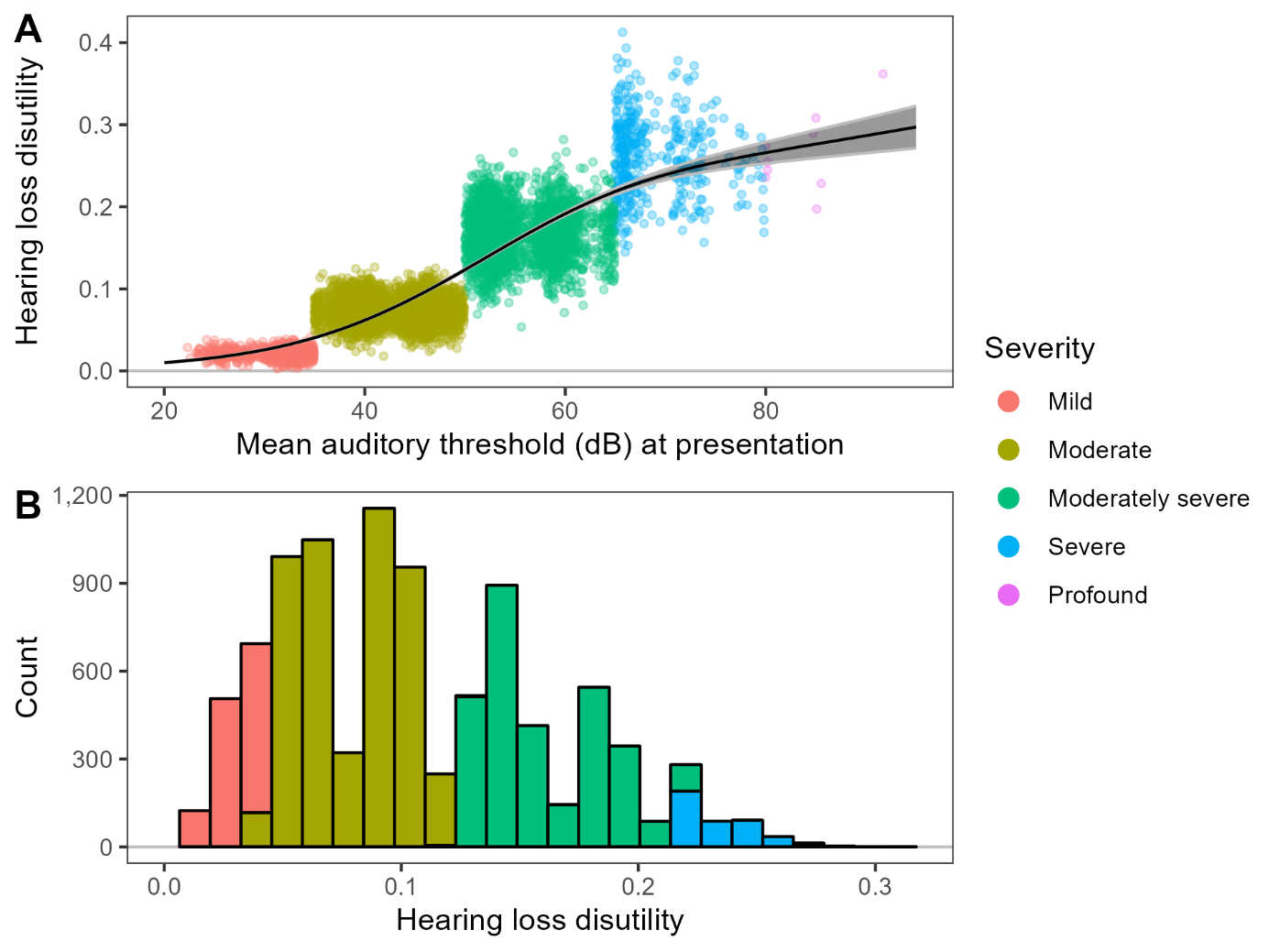

***Figure S12****. Hearing loss disability. (****A****) Disability associated with sensorineural hearing loss (SNHL), simulated from estimates of disability weights for hearing loss with ringing from the Global Burden of Disease (GBD) study on hearing loss (2021) corresponding to mean auditory threshold (MAT) values specific to Lassa fever-induced SNHL. Each point is a value drawn randomly as a function of the MAT in decibels (dB) at baseline from audiometry measurements in patients followed-up in a prospective cohort study of Lassa fever-induced SNHL by Cummins et al. (1990),^27^ and subsequently binned according to the corresponding severity category defined in GBD (2021).^28^ The smoothed curve shows the mean fit (line) and 95% confidence interval (shading) of a generalised additive model fit to these data. (****B****) Histogram of the final distribution of disability predicted by the GAM as a function of the bootstrapped sample of baseline MAT values from Cummins et al. (1990).*

***1f. Vaccination Campaigns***

Vaccination campaigns were included in the model to estimate the projected health-economic benefits of administering a hypothetical single-dose Lassa fever vaccine preventively to different population groups in endemic areas of West Africa. Vaccination campaigns included in the model were conducted only in areas classified as endemic (≥5 Lassa fever cases reported annually) following WHO’s 2024 Lassa fever risk map.^5^ Therefore, vaccine doses were administered to populations in 19 areas spread across Nigeria (14), Liberia (3), Guinea (1) and Sierra Leone (1). Any area not classified as endemic (<5 cases reported annually) did not receive any vaccine doses. Within each area, vaccination campaigns were designed to target 75% of the entire population aged ≥2 years (“untargeted vaccination”) or 75% of one of four target groups defined based on age and sex (“risk-targeted vaccination”). The four target groups considered were children aged 2 to 14 years, women of childbearing age (WCBA) aged 15 to 49 years, all adolescents and adults aged 15 to 49 years, and older adults aged ≥50 years. The 75% total coverage target was based on targets for other mass vaccination campaigns and a previously published Lassa fever vaccine demand forecast.^29-31^

The 3-year campaign was intended to reduce the annual demand after introduction by spreading the campaign over multiple years (**Table S4**). The total demand for each target age group assumed 10% wastage per year and accounted for the changing size of each group over time (per population projections). Any campaign in Nigeria requires significantly more doses than those operated in the other endemic countries, due to its much more populous endemic areas. These demand numbers are driven by target coverage, target population and wastage and are therefore not constrained by supply. The final number of vaccine doses administered under each vaccination campaign was assumed to be equal to vaccine demand.

***Table S4.*** *Total vaccine demand for target groups in endemic areas grouped by year. The demand figures assume 75% population coverage achieved over a 3-year campaign, a 1-dose schedule and 10% wastage, and account for projected population change over the vaccination period. WCBA = women of childbearing age; M = million.*

| **Target group  (age in years)** | **2025** | **2026** | **2027** | **Total** |
| --- | --- | --- | --- | --- |
| Children (2-14) | 8.34 M | 8.40 M | 8.46 M | 25.20 M |
| WCBA (15-49) | 5.86 M | 6.02 M | 6.19 M | 18.06 M |
| Adolescents and adults (15-49) | 11.90 M | 12.23 M | 12.58 M | 36.70 M |
| Older adults (50+) | 2.51 M | 2.59 M | 2.68 M | 7.78 M |
| All (2+) | 22.7 M | 23.23 M | 23.72 M | 69.68 M |

To simulate vaccination cohorts, immunisation was implemented using an algorithm that accounts for aging, such that the age groups in which outcomes are prevented represent individuals’ ages when those outcomes would have occurred, and not necessarily their age when vaccine was received. The age-specific benefits of targeted vaccination thus depend not only on the burden of disease in the targeted group and the vaccine’s efficacy against disease, but also on how much time has passed since vaccine administration. The acquisition and loss of immunity through time when targeting different age groups is visualised in **Figure S13**.

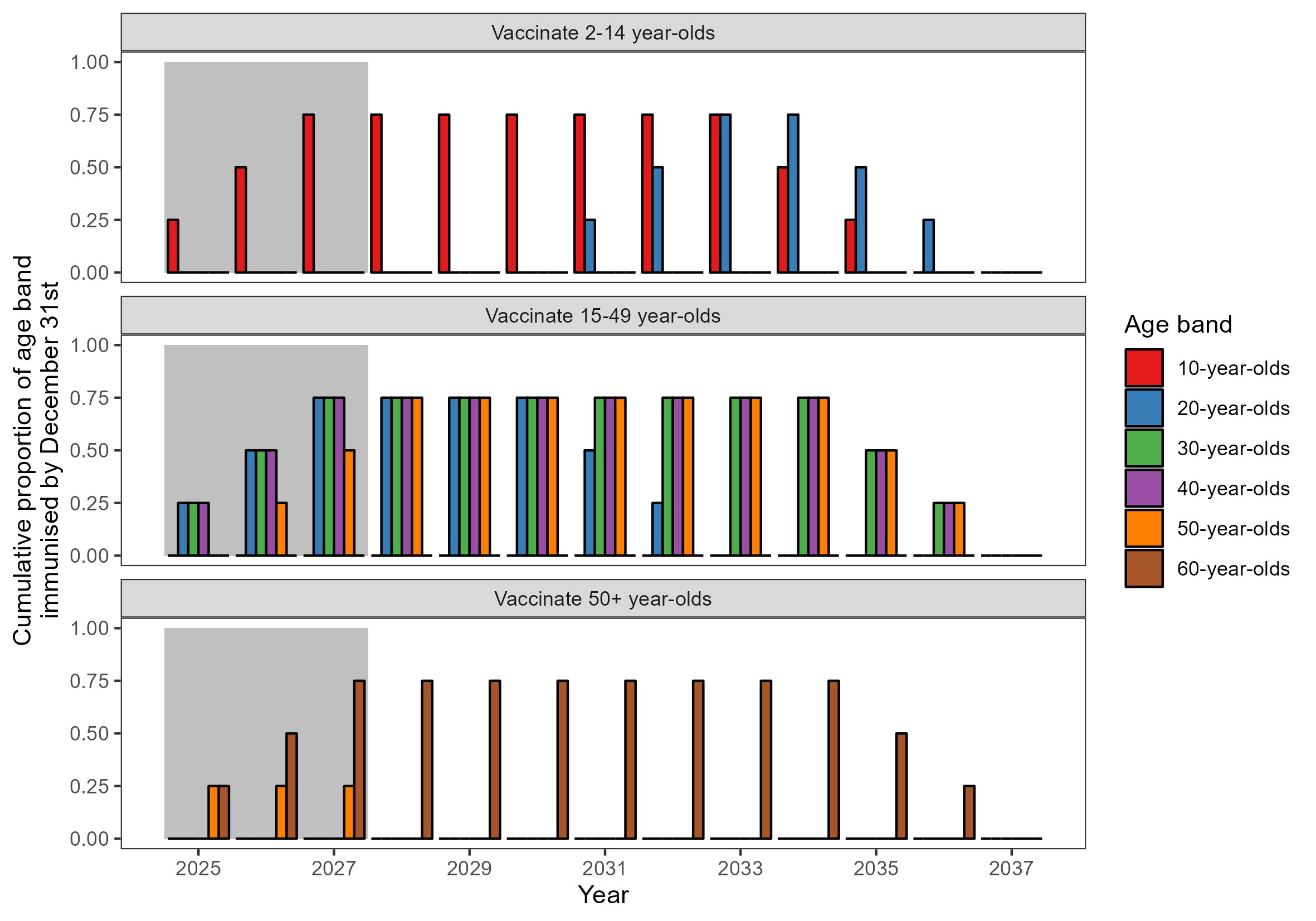

***Figure S13.*** *Levels of vaccine-induced immunity at year’s end (31st December) across selected one-year age bands (colours) for different vaccine population target groups (panels). Changes in the proportion immunised after the end of the vaccine programme (grey shaded area) reflect vaccinated individuals aging into older age groups, and eventual immune waning ten years after vaccine administration. These immunisation proportions therefore demonstrate potential immunogenicity due to prior vaccine exposure and do not account for imperfect vaccine efficacy or natural immunity (e.g. through prior LASV infection).*

***1g. Other health state parameters***

Other health state input parameters in the model remain unchanged since previous publication, including the probability of seeking outpatient care for mild or moderate symptoms, as well as the durations and disability weights of various health states. See Smith *et al*. (2024) for details on derivation of these parameters.^1^ **Table S5** lists all health state input parameters not stratified by age and sex.

***Table S5.*** *Model input parameters not varied by age or sex. The mean and 95% uncertainty interval were calculated from the final vector of 500 stochastic draws for each parameter used as inputs in Monte Carlo simulation. No unit is provided for disability weights, which represent decrements of health-related quality of life on a scale from 0 (death) to 1 (perfect health). CI = confidence interval.*

| **Parameter (unit) *Symbol*** | **Mean  [95% CI]** | **Distribution [parameters]** | **Source** |
| --- | --- | --- | --- |
| ***Health state probabilities*** |  |  |  |
| Probability of foetal loss in pregnant women hospitalised with Lassa fever (%)  $\mathbb{P}\left( FL\vert hospital \right)$ | 60.3 [30.8, 83.3] | Beta  [*α*=6.913, *β*=4.533] | Estimated |
| Probability of all-cause foetal loss (%)  $\mathbb{P}\left( FL\vert baseline \right)$ | 2.28 [1.98, 2.77] | Log-normal  [*μ_log_*=-3.78, *σ_log_*=0.086] | ^19^ |
| Probability of neonatal death in pregnant women hospitalised with Lassa fever (%)  $\mathbb{P}\left( NND\vert hospital \right)$ | 38.1 [21.8, 58.3] | Beta  [*α*=9.89, *β*=16.08] | Estimated |
| Probability of all-cause neonatal death (%)  $\mathbb{P}\left( NND\vert baseline \right)$ | 4.97 [4.45, 5.51] | Log-normal  [*μ_log_*=-3.00, *σ_log_*=0.055] | ^20^ |
| Probability of developing SNHL during Lassa fever convalescence (%)  $\mathbb{P}\left( SNHL \vert discharged \right)$ | 28.0 [16.3, 42.9] | Binomial,  [*n*=49, *p*=0.286] | ^27^ |
| Probability of seeking outpatient care for mild/moderate symptoms (%)  $\mathbb{P}\left( outpatient \vert mild \right)$ | 59.7 [54.5, 65.3] | Normal,  [*μ*=0.598, *σ*=0.028] | ^32^ |
| Probability of outpatient care costs being reimbursed by government (%)  $\mathbb{P}\left( gvt \vert outpatient \right)$ | 48.8 [43.7, 54.5] | Normal,  [*μ*=0.489, *σ*=0.028] | ^32^ |
| ***Health state durations*** |  |  |  |
| Duration of mild/moderate symptoms (days)  ${dur}^{mild}$ | 3.53 [3.29, 3.78] | Normal,  [*μ*=3.53, *σ*=0.123] | ^33^ |
| Duration of severe symptoms prior to hospital (days)  ${dur}_{pre-hospital}^{severe}$ | 9.33 [8.95, 9.73] | Normal,  [*μ*=9.33, *σ*=0.197] | ^14^ |
| Hospital length of stay for survivors (days)  ${dur}_{hospital_{survived}}^{severe}$ | 12.00 [11.66, 12.35] | Normal,  [*μ*=12, *σ*=0.175] | ^14^ |
| Hospital length of stay for fatalities (days)  ${dur}_{hospital_{died}}^{severe}$ | 3.32 [2.42, 4.29] | Normal,  [*μ*=3.33, *σ*=0.475] | ^14^ |
| Duration of Lassa fever-induced SNHL (years)  ${dur}^{SNHL}$ | 2.10 [1.07, 6.07] | Non-parametric | Estimated |
| ***Health state disability weights*** |  |  |  |
| Disability due to mild/moderate symptoms  ${dw}^{mild}$ | 0.0508 [0.0304, 0.0725] | Normal  [*μ*=0.051, *σ*=0.011] | ^34^ |
| Disability due to severe symptoms  ${dw}^{severe}$ | 0.337 [0.219, 0.429] | Non-parametric | ^1^ |
| Disability due to Lassa fever-induced SNHL  ${dw}^{SNHL}$ | 0.104 [0.032, 0.215] | Non-parametric | Estimated |

***1h. Economic parameters***

All economic parameters included in our model were estimated previously (see Smith *et al.* (2024) for details), but were reviewed and updated to 2023 value here using most recent available economic data.^1^ These updated parameters are summarised in **Table S6**. The 2023 World Bank GDP deflator was used to update country-specific outpatient and inpatient treatment costs, including the proportion of costs paid out-of-pocket (OOP) by patients or their families, based on modelled estimates of all-cause outpatient costs and data from an inpatient Lassa fever costing study in Nigeria in 2016.^35-37^ These estimates of OOP healthcare expenditure informed updated estimates of the probability of OOP healthcare expenditure resulting in: (i) catastrophic healthcare expenditure (CHE), i.e. healthcare expenditure exceeding 10% of annual income; and (ii) impoverishing healthcare expenditure (IHE), estimated using updated country-specific poverty lines from 2017 (USD$3.40 in Guinea, USD$3.24 in Sierra Leone, USD$3.13 in Liberia and USD$2.52 in Nigeria).^38^ Finally, 2023 World Bank estimates of per-capita GDP were used to update country-specific estimates of (i) willingness-to-pay per DALY from Ochalek *et al*., and (ii) the value of a statistical life.^36,39-41^

***Table S6.*** *Economic model input parameters. All parameters were estimated previously in Smith et al. (2024) and were updated here to values reflecting International dollars in 2023 (I$ 2023).*

| **Parameter (Unit) *Symbol*** | **Guinea** | **Liberia** | **Nigeria** | **Sierra Leone** |
| --- | --- | --- | --- | --- |
| Outpatient unit cost (I$ 2023)  ${Unit}_{c}^{outpatient}$ | 6.11 | 15.88 | 23.21 | 9.77 |
| Inpatient unit cost, reimbursed (I$ 2023)  ${Unit}_{c}^{inpatient,gvt}$ | 1,723 | 2,114 | 1,414 | 1,751 |
| Inpatient unit cost, paid OOP (I$ 2023)  ${Unit}_{c}^{inpatient,OOP}$ | 725 | 334 | 1,034 | 697 |
| Probability of CHE (%)  $\mathbb{P}_{c}\left( catastrophic\vert hospital \right)$ | 99.3 | 94.9 | 99.9 | 98.3 |
| Probability of IHE (%)  $\mathbb{P}_{c}\left( impoverishing\vert hospital \right)$ | 55.6 | 44.0 | 59.8 | 41.5 |
| Willingness-to-pay per DALY (I$ 2023)  $m_{c}$ | 449 | 168 | 178 | 91 |
| Value of statistical life (I$ 2023)  ${vsl}_{c}$ | 138,544 | 39,266 | 273,485 | 55,070 |

***APPENDIX 2: OUTCOME CALCULATIONS***

***2a. Disease states***

The total number of cases of symptomatic infection in each year *y*, area *d*, 1-year age band *a* and sex *s* (including males, non-pregnant females and pregnant females) is calculated as,

$$N_{y,d,s,a}^{symptoms}=L_{y,d,s,a}\cdot\mathbb{P}_{a}(symptoms|infection)$$

Severe Lassa fever is defined as symptomatic disease that is severe enough to result in hospitalisation, quantified using age- and sex-specific IHR estimates, such that

$$N_{y,d,s,a}^{hospital}=N_{y,d,s,a}^{severe}=L_{y,d,s,a}\cdot\mathbb{P}_{s,a}(hospital|infection)$$

The number of cases of mild or moderate symptomatic disease is therefore,

$$N_{y,d,s,a}^{mild}=N_{y,d,s,a}^{symptoms}-N_{y,d,s,a}^{severe}$$

and the number of outpatient healthcare visits resulting from mild or moderate disease is

$$N_{y,d,s,a}^{outpatient}=N_{y,d,s,a}^{mild}\mathbb{\cdot P(}outpatient|mild)$$

among which a proportion are in government-run facilities,

$$N_{y,d,s,a}^{outpatient,gvt}=N_{y,d,s,a}^{outpatient}\mathbb{\cdot P(}gvt|outpatient)$$

Among hospitalised cases, the number of deaths is calculated using age- and sex-adjusted CFRs,

$$N_{y,d,s,a}^{death}=N_{y,d,s,a}^{hospital}\cdot\mathbb{P}_{s,a}(death|hospital)$$

and, among pregnant women (*s*=*P*), the number of foetal losses due to Lassa fever is calculated as

$$N_{y,d,s=P,a}^{FL}=N_{y,d,s=P,a}^{hospital}\cdot\left( \mathbb{P}\left( FL|hospital \right)\mathbb{-P}\left( FL|baseline \right) \right)$$

and the number of neonatal deaths due to Lassa fever as

$$N_{y,d,s=P,a}^{NND}=N_{y,d,s=P,a}^{hospital}\cdot\left( \mathbb{P}\left( NND|hospital \right)\mathbb{-P}\left( NND|baseline \right) \right)$$

where neonatal deaths and foetal losses were attributed to the youngest age (*a* = 0), with equal proportions male and female, for appropriate calculation of DALYs. Foetal loss is presented as a distinct outcome separate from deaths, but the total number of deaths is presented as including both direct Lassa fever deaths and neonatal deaths following maternal Lassa fever,

$$N_{y,d,s,a}^{deathTotal}=N_{y,d,s,a}^{death}+N_{y,d,s,a}^{NND}$$

Finally, among survivors, the number that develop SNHL during convalescence was calculated,

$$N_{y,d,s,a}^{SNHL}=N_{y,d,s,a}^{mild}\mathbb{\cdot P}\left( SNHL | mild \right)+\left( N_{y,d,s,a}^{hospital}-N_{y,d,s,a}^{death} \right)\mathbb{\cdot P}\left( SNHL | discharged \right)$$

where in the base case it is assumed that

$$\mathbb{P}\left( SNHL | mild \right)\mathbb{=P}\left( SNHL | discharged \right)$$

and in sensitivity analysis it is assumed that

$$\mathbb{P}\left( SNHL | mild \right)=0$$

***2b. Disability-adjusted life years***

As in Smith *et al.* (2024),^1^ disability-adjusted life years (DALYs) due to mild or moderate symptomatic disease were calculated as the disability weight associated with fever multiplied by its cumulative duration,

$${DALY}^{mild}=\sum_{y} \sum_{d} \sum_{s} \sum_{a} N_{y,d,s,a}^{mild}\cdot\frac{{dw}^{mild}}{365}\cdot{dur}^{mild}$$

DALYs due to severe symptomatic disease were calculated as the disability weight associated with hospitalised Lassa fever multiplied by its cumulative duration, accounting for different illness durations among those who die and those who survive,

$${DALY}^{severe}=\sum_{y} \sum_{d} \sum_{s} \sum_{a} \left( \left( N_{y,d,s,a}^{hospital}-N_{y,d,s,a}^{death} \right)\cdot\frac{{dw}^{severe}}{365}\cdot\left( {dur}_{pre-hospital}^{severe}+{dur}_{hospital_{survived}}^{severe} \right)+N_{y,d,s,a}^{death}\cdot\frac{{dw}^{severe}}{365}\cdot\left( {dur}_{pre-hospital}^{severe}+{dur}_{hospital_{died}}^{severe} \right) \right)$$

DALYs due to SNHL were calculated using estimates of the average disability associated with Lassa fever-induced hearing loss multiplied by estimates of its average cumulative duration,

$${DALY}^{SNHL}=\sum_{y} \sum_{d} \sum_{s} \sum_{a} N_{y,d,s,a}^{SNHL}\cdot{dw}^{SNHL}\cdot{dur}^{SNHL}$$

DALYs due to death are equal to each deceased individual’s average remaining life expectancy. For a death occurring at age *a*, annual national projections of age- and sex-specific remaining life expectancy from 2024 WHO Population Prospects, *x_y,c,s,a_*, were used to account for changing life expectancy over the study horizon,^6^

$${DALY}^{deathTotal}=\sum_{y} \sum_{d} \sum_{s} \sum_{a} N_{y,d,s,a}^{deathTotal}\cdot x_{y,c,s,a}$$

The same holds for DALYs due to foetal loss, although this outcome was only included in sensitivity analysis,

$${DALY}^{FL}=\sum_{y} \sum_{d} \sum_{s} \sum_{a} N_{y,d,s,a}^{FL}\cdot x_{y,c,s,a}$$

The total number of DALYs due to Lassa fever is therefore,

$${DALY}^{total}={DALY}^{mild}+{DALY}^{severe}+{DALY}^{SNHL}+{DALY}^{deathTotal}+{DALY}^{FL}$$

***2c. Treatment costs***

As with all monetary costs, outpatient care costs among patients with mild or moderate symptomatic disease were calculated using a discounting rate, *r*, applied in discrete time to future costs to estimate their present value in 2025, i.e. at the start of the study time horizon,

$${Cost}_{r}^{outpatient,gvt}=\sum_{y} \sum_{d} \sum_{s} \sum_{a} \frac{N_{y,d,s,a}^{outpatient,gvt}\cdot{Unit}_{c}^{outpatient}}{\left( 1+r \right)^{y-2025}}$$

$${Cost}_{r}^{outpatient,OOP}=\sum_{y} \sum_{d} \sum_{s} \sum_{a} \frac{\left( N_{y,d,s,a}^{outpatient}-N_{y,d,s,a}^{outpatient,gvt} \right)\cdot{Unit}_{c}^{outpatient}}{\left( 1+r \right)^{y-2025}}$$

$${Cost}_{r}^{outpatient}={Cost}_{r}^{outpatient,gvt}+{Cost}_{r}^{outpatient,OOP}$$

And inpatient costs for patients hospitalised with Lassa fever were calculated as,

$${Cost}_{r}^{hospital,gvt}=\sum_{y} \sum_{d} \sum_{s} \sum_{a} \frac{N_{y,d,s,a}^{hospital}\cdot{Unit}_{c}^{inpatient,gvt}}{\left( 1+r \right)^{y-2025}}$$

$${Cost}_{r}^{hospital,OOP}=\sum_{y} \sum_{d} \sum_{s} \sum_{a} \frac{N_{y,d,s,a}^{hospital}\cdot{Unit}_{c}^{inpatient,OOP}}{\left( 1+r \right)^{y-2025}}$$

$${Cost}_{r}^{hospital}={Cost}_{r}^{hospital,gvt}+{Cost}_{r}^{hospital,OOP}$$

Total Lassa fever healthcare costs are therefore,

$${Cost}_{r}^{care}={Cost}_{r}^{outpatient}+{Cost}_{r}^{hospital}$$

Related to the cost of hospitalisation costs paid OOP, the number of instances of Lassa fever hospitalisation costs resulting in CHE or IHE were calculated,

$$N^{catastrophic}=\sum_{y} \sum_{d} \sum_{s} \sum_{a} N_{y,d,s,a}^{hospital}\mathbb{\cdot P(}catastrophic|hospital)$$

$$N^{impoverishing}=\sum_{y} \sum_{d} \sum_{s} \sum_{a} N_{y,d,s,a}^{hospital}\mathbb{\cdot P(}impoverishing|hospital)$$

Outpatient costs did not contribute to catastrophic or impoverishing expenditure, under the assumption that the probability of attending a government-run facility is associated with having less ability to pay OOP for medical expenses.

***2d. Monetised DALYs***

When monetising DALYs, it is necessary to discount future years of life lost or lived with disability. Following Larson,^42^ the total years of life lost (YLL) due to Lassa fever mortality were calculated assuming continuous time discounting at rate *r* as,

$${YLL}_{r,y,d}=\sum_{s} \sum_{a} N_{y,d,s,a}^{death}\frac{1-e^{-r\cdot x_{y,d,s,a}}}{r}+\left( N_{y,d,s,a}^{NND}+N_{y,d,s,a}^{FL} \right)\frac{1-e^{-r\cdot x_{y,d,s,a=0}}}{r}$$

where (monetised) DALYs due to foetal losses were only included in sensitivity analysis. Half-cycle correction was not applied due to the majority of Lassa fever hospitalisations occurring in January and February of each year.

Years of life lived with disability (YLD) due to Lassa fever-induced SNHL of average duration *dur^SNHL^* years were calculated as,

$${YLD}_{r,y,d}=\sum_{s} \sum_{a} N_{y,d,s,a}^{SNHL}\frac{1-e^{-r\cdot{dur}^{SNHL}}}{r}$$

It was necessary to truncate SNHL duration in 1 of 500 (0.2%) Monte Carlo simulations in which *x_y,d,s,a_* > *dur^SNHL^* in those aged 80+.

In turn, monetised DALYs for Lassa fever cases occurring in year *y* were calculated using country-specific willingness-to-pay thresholds, *m_c_*, as

$${MDALY}_{r}=\sum_{y} \sum_{d} \frac{{DALY}_{r,y,d}^{mild}+{DALY}_{r,y,d}^{severe}+{YLL}_{r,y,d}+{YLD}_{r,y,d}}{\left( 1+r \right)^{y-2025}}\cdot m_{c}$$

***2e. Productivity losses***

To estimate future years of potential productive life lost (YPPLL) due to Lassa fever, most recent age group- and sex-specific estimates of the proportion of the population participating in the labour force in each country, *lfp_c,a,s_*, compiled by the International Labour Organisation (ILO) were used,^43^

$${YPPLL}_{r,y,d}^{mild}=\sum_{s} \sum_{a} N_{y,d,s,a}^{mild}\cdot{dur}^{mild}\cdot\frac{{lpf}_{c,s,a}}{365}$$

$${YPPLL}_{r,y,d}^{severe}=\sum_{s} \sum_{a} \left( N_{y,d,s,a}^{hospital}-N_{y,d,s,a}^{death} \right)\cdot\left( {dur}_{pre-hospital}^{severe}+{dur}_{hospital_{survived}}^{severe} \right)\cdot\frac{{lpf}_{c,s,a}}{365}+N_{y,d,s,a}^{death}\cdot\left( {dur}_{pre-hospital}^{severe}+{dur}_{hospital_{died}}^{severe} \right)\cdot\frac{{lpf}_{c,s,a}}{365}$$

For YPPLL associated with death and disability, discrete time discounting was applied over future years of lost productivity because of variable *lfp_c,a,s_* over individuals’ remaining years of life (death) or years of life lived with disability (SNHL),

$${YPPLL}_{r,y,d}^{death}=\sum_{s} \sum_{a} N_{y,d,s,a}^{death}\cdot\left( \sum_{i=0}^{\left\lfloor x_{y,d,s,a} \right\rfloor-1} \frac{{lpf}_{c,s,a+i}}{\left( 1+r \right)^{i}} \right)+\frac{\vartheta_{y,d,s,a}^{death}\cdot{lpf}_{c,s,a+\left\lfloor x_{y,d,s,a} \right\rfloor}}{\left( 1+r \right)^{\left\lfloor x_{y,d,s,a} \right\rfloor}}$$

$${YPPLL}_{r,y,d}^{SNHL}=\sum_{s} \sum_{a} N_{y,d,s,a}^{SNHL}\cdot\left( \sum_{i=0}^{\left\lfloor{dur}^{SNHL} \right\rfloor-1} \frac{{lpf}_{c,s,a+i}\cdot\left( 1-{pl}^{SNHL} \right)}{\left( 1+r \right)^{i}} \right)+\frac{\vartheta^{SNHL}\cdot{lpf}_{c,s,a+\left\lfloor{dur}^{SNHL} \right\rfloor}\cdot\left( 1-{pl}^{SNHL} \right)}{\left( 1+r \right)^{\left\lfloor{dur}^{SNHL} \right\rfloor}}$$

where non-integer durations of remaining life expectancy and SNHL are given by

$$x_{y,d,s,a}\equiv\vartheta_{y,d,s,a}^{death} \left( \mathrm{mod} x_{y,d,s,a} \right)$$

and

$${dur}^{SNHL}\equiv\vartheta^{SNHL} \left( \mathrm{mod} {dur}^{SNHL} \right)$$

The calculation for *YPPLL^SNHL^* includes *pl^SNHL^*, a proportional reduction in labour force participation due to SNHL. Estimates specific to Lassa fever could not be found, but a 2017 WHO report has highlighted the global costs of unaddressed hearing loss, including lost economic productivity among those with hearing loss due to isolation, communication difficulties, stigma and other factors (WHO, 2017a).^44^ Although this report highlights that information on productivity losses related to hearing loss remains scarcely available even in high-income settings, the authors highlighted a conservative estimate that there is an 18% gap in employment rates between working-age people not classified as disabled relative to working-age disabled people reporting hearing loss as their main health problem. This estimate of 18% has also been used in a study estimating the global costs of hearing loss.^45^ It was therefore assumed that *pl^SNHL^* = 0.18 among those experiencing SNHL for the duration of their disability.

Total productivity losses were then calculated by summing annual national YPPLL multiplied by per-capita gross national income (GNI) and discounting annually,

$${PL}_{r}=\sum_{y} \sum_{d} \frac{{YPPLL}_{r,y,d}^{mild}+{YPPLL}_{r,y,d}^{severe}+{YPPLL}_{r,y,d}^{death}+{YPPLL}_{r,y,d}^{SNHL}}{\left( 1+r \right)^{y-2025}}\cdot{GNI}_{c}$$

Based on these calculations, age- and sex-stratified per-person productivity losses due to death (*YPPLL^death^*) vary across included countries (**Figure S14**), over time (**Figure S15**) and depending on the discount rate assumed (**Figure S16**). Per-person productivity losses due to SNHL (*YPPLL^SNHL^*) also vary based on these factors, as well as on draws of the average duration of SNHL in Monte Carlo simulation (**Figure S17**).

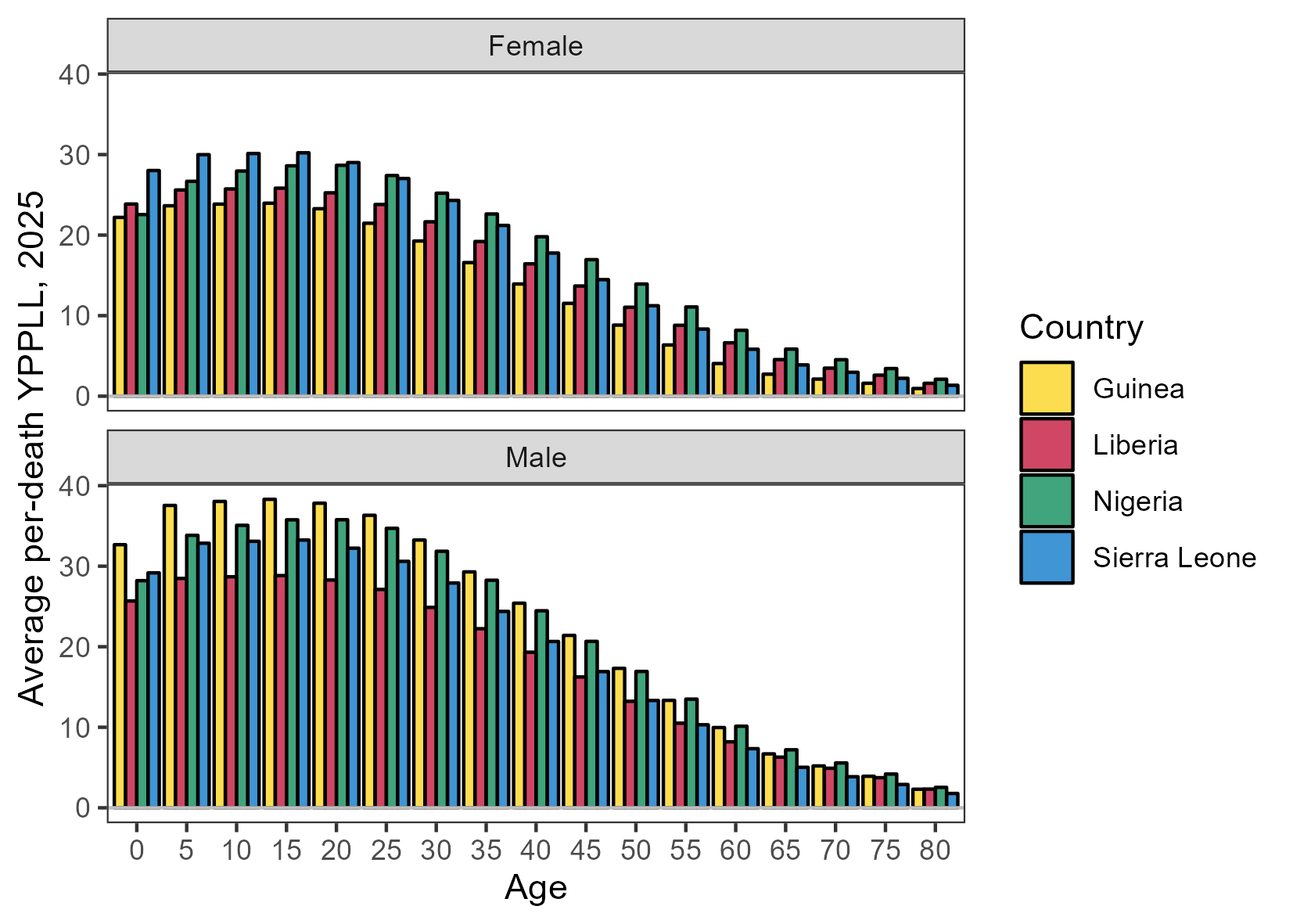

***Figure S14.*** *The average YPPLL per Lassa fever death across countries (colours), stratified by sex (panels) and age (x-axis), showing a selection one-year age bands from 0 to 80+ years by increments of 5 years and assuming no discounting of future YPPLL (r=0%).*

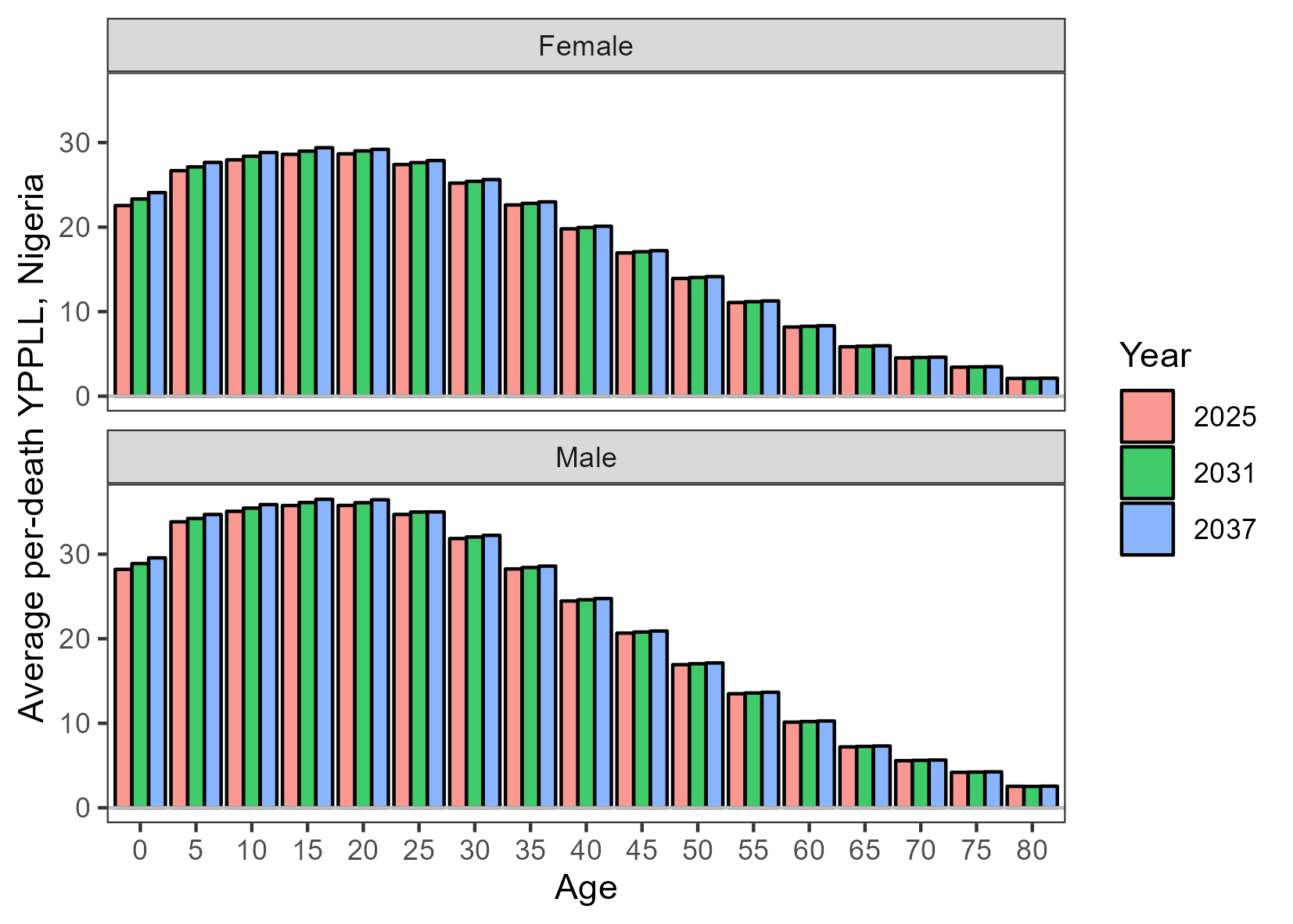

***Figure S15.*** *The average YPPLL per Lassa fever death across a selection of 3 years from the beginning, middle and end of the modelled time horizon (colours), stratified by sex (panels) and age (x-axis), showing a selection of one-year age bands from 0 to 80+ years by increments of 5 years and assuming no discounting of future YPPLL (r=0%).*

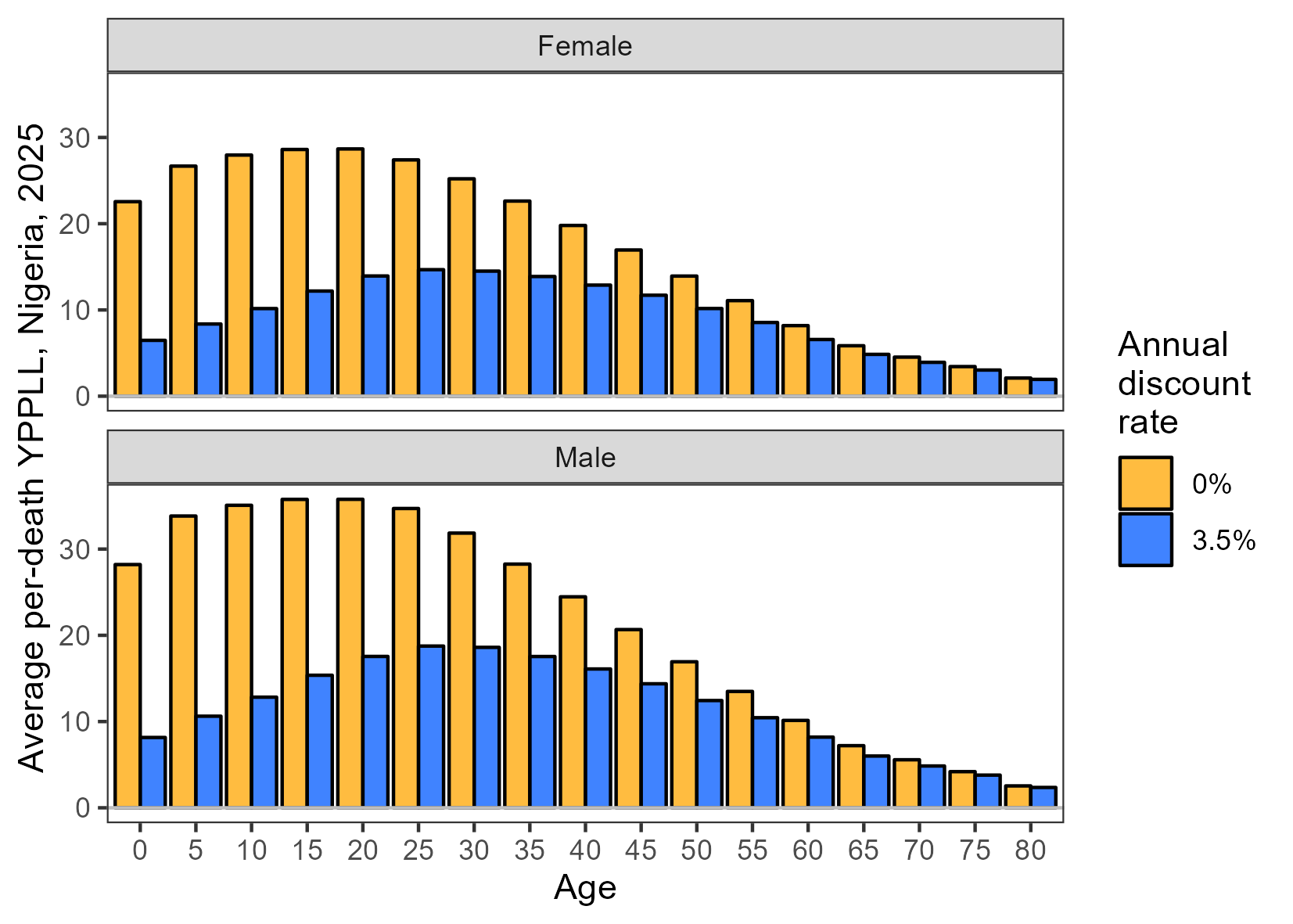

***Figure S16.*** *The average YPPLL per Lassa fever death with and without discounting of future life-years (colours), stratified by sex (panels) and age (x-axis), showing a selection of one-year age bands from 0 to 80+ years by increments of 5 years.*

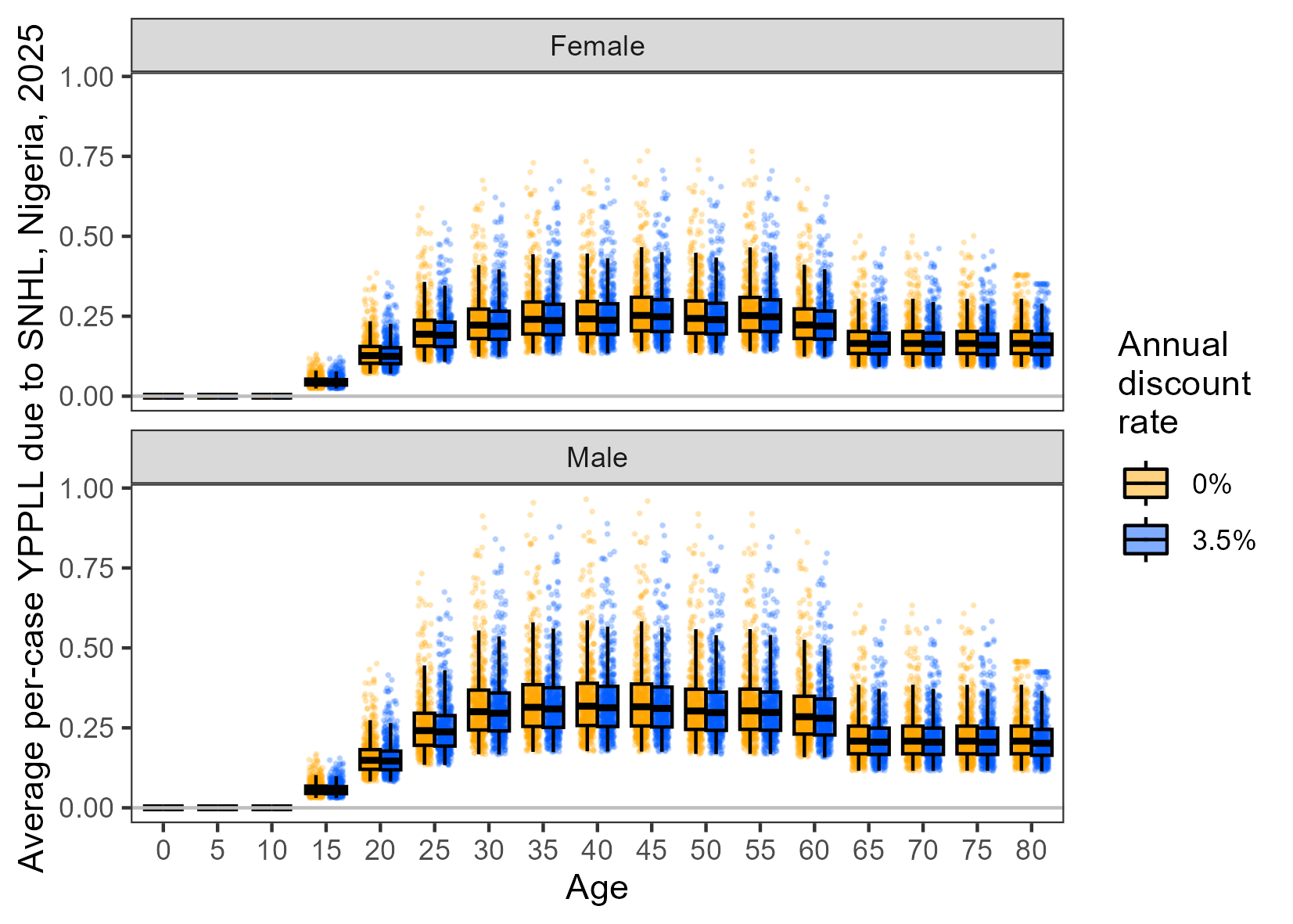

***Figure S17.*** *The average YPPLL per case of Lassa fever-induced SNHL with and without discounting of future life-years (colours), stratified by sex (panels) and age (x-axis), showing a selection of one-year age bands from 0 to 80+ years by increments of 5 years. Each point (n=500) corresponds to a stochastic draw of SNHL duration used in Monte Carlo simulation. Boxplot whiskers extend to the most extreme value no further than 1.5 × IQR from the nearest hinge.*

***2f. Societal costs***

The economic burden of disease from a societal perspective includes both direct costs of disease (e.g. healthcare costs), as well as indirect costs (e.g. productivity losses) and intangible costs (e.g. reduced quality of life). Societal costs of Lassa fever were therefore calculated as the sum of these costs,

$${SC}_{r}={Cost}_{r}^{care}+{MDALY}_{r}+{PL}_{r}$$

***2g. Value of statistical life-years***

In addition to the value of statistical life (VSL), as calculated in Smith *et al*. (2024),^1^ the value of statistical life-years (VSLY) was also calculated. VSLY divides VSL by a given population’s life expectancy at birth, *x_y,c,s,a=0_*, and spreads the estimated per-year value across the number of expected remaining years of life left to live (equivalent to undiscounted DALYs due to death),

$$VSLY=\sum_{y} \sum_{d} \sum_{s} \sum_{a} \frac{{DALY}_{y,d,s,a}^{deathTotal}\cdot{vsl}_{c}}{x_{y,d,s,a=0}}$$

VSLY thus places greater value on younger people, who have more years left to live and hence greater total VSLY.

***2h. Threshold vaccine costs***

The threshold vaccine cost (TVC) is the price per vaccine dose at which the benefit-to-cost ratio of vaccination exceeds 1. This was calculated as the value of an economic burden outcome *b* averted due to vaccination divided by *v_y_^g^*, the number of vaccine doses allocated in year *y* when targeting risk group *g*, including wasted doses (10%) and discounting future vaccine doses at 3% per year.

$${TVC}_{r}^{b,g}=\frac{b_{r}}{\sum_{y} \frac{v_{y}^{g}}{{(1+r)}^{y-2025}}}$$

TVCs were calculated separately for each of *b* in *SC* (base case analysis) and *VSLY* (sensitivity analysis).

***APPENDIX 3: GATHER CHECKLIST***

**Table S7.** Guidelines for Accurate and Transparent Health Estimates Reporting (GATHER) checklist.^46^

| **Item** | **Checklist item** | **Reporting** |
| --- | --- | --- |
| **Objectives and funding** | | |
| **1** | Define the indicator(s), populations (including age, sex, and geographic entities), and time period(s) for which estimates were made. | Methods [*Model outcomes*] |
| **2** | List the funding sources for the work. | Methods [*Role of the funder*] |
| **Data inputs** | | |
| *For all data inputs from multiple sources that are synthesized as part of the study:* | | |
| **3** | Describe how the data were identified and how the data were accessed. | Methods [*Model overview*]; Supplementary appendix 1 |
| **4** | Specify the inclusion and exclusion criteria. Identify all ad‐hoc exclusions. | Supplementary appendix 1 |
| **5** | Provide information on all included data sources and their main characteristics. For each data source used, report reference information or contact name/institution, population represented, data collection method, year(s) of data collection, sex and age range, diagnostic criteria or measurement method, and sample size, as relevant. | Tables S1 through S4; Supplementary appendix 1 |
| **6** | Identify and describe any categories of input data that have potentially important biases (e.g., based on characteristics listed in item 5). | Supplementary appendix 1 |
| *For data inputs that contribute to the analysis but were not synthesized as part of the study:* | | |
| **7** | Describe and give sources for any other data inputs. | Tables S5 and S6 |
| *For all data inputs:* | | |
| **8** | Provide all data inputs in a file format from which data can be efficiently extracted (e.g., a spreadsheet rather than a PDF), including all relevant meta‐data listed in item 5. For any data inputs that cannot be shared because of ethical or legal reasons, such as third‐party ownership, provide a contact name or the name of the institution that retains the right to the data. | Methods [*Code and data sharing*] |
| **Data analysis** | | |
| **9** | Provide a conceptual overview of the data analysis method. A diagram may be helpful. | Figure 2 |
| **10** | Provide a detailed description of all steps of the analysis, including mathematical formulae. This description should cover, as relevant, data cleaning, data pre‐processing, data adjustments and weighting of data sources, and mathematical or statistical model(s). | Supplementary appendix 2 |
| **11** | Describe how candidate models were evaluated and how the final model(s) were selected. | Methods [*Simulation and statistical reporting*]; Supplementary appendix 1 |
| **12** | Provide the results of an evaluation of model performance, if done, as well as the results of any relevant sensitivity analysis. | Methods [*Simulation and statistical reporting*]; Figure S30 |
| **13** | Describe methods for calculating uncertainty of the estimates. State which sources of uncertainty were, and were not, accounted for in the uncertainty analysis. | Methods [*Simulation and statistical reporting*] |
| **14** | State how analytic or statistical source code used to generate estimates can be accessed. | Methods [*Code and data sharing*] |
| **Results and Discussion** | | |
| **15** | Provide published estimates in a file format from which data can be efficiently extracted. | Results tables |
| **16** | Report a quantitative measure of the uncertainty of the estimates (e.g. uncertainty intervals). | Reported throughout |
| **17** | Interpret results in light of existing evidence. If updating a previous set of estimates, describe the reasons for changes in estimates. | Discussion |
| **18** | Discuss limitations of the estimates. Include a discussion of any modelling assumptions or data limitations that affect interpretation of the estimates. | Discussion |

***APPENDIX 4: SUPPLEMENTARY RESULTS***

***4a. Projected Lassa fever burden in the absence of vaccination***

***Table S8.*** *Projected cumulative burden of Lassa fever health outcomes from 2025 to 2037 in the absence of vaccination, stratified by age group. DALYs do not include DALYs due to foetal loss. Hearing loss refers to sensorineural hearing loss, DALY = disability-adjusted life-year.*

| **Age group** | **LASV infection** | **Symptomatic cases** | **Hospitalisation** | **Hearing loss** | **Death** | **Foetal loss** | **DALYs** |
| --- | --- | --- | --- | --- | --- | --- | --- |
| *Mean (95% uncertainty interval) cumulative totals* | | | | | | | |
| <2 | 1.10 M (942 K - 1.27 M) | 212 K (115 K - 363 K) | 1.31 K (835 - 1.95 K) | 60.7 K (25.7 K - 118 K) | 995 (402 - 1.71 K) | 1.60 K (710 - 2.56 K) | 69.7 K (30.2 K - 122 K) |
| 2-14 | 4.00 M (3.76 M - 4.24 M) | 770 K (410 K - 1.29 M) | 10.0 K (6.72 K - 13.5 K) | 221 K (93.6 K - 434 K) | 223 (0 - 559) | 0 (0 - 0) | 60.4 K (15.6 K - 157 K) |
| 15-24 | 1.76 M (1.60 M - 1.91 M) | 339 K (175 K - 577 K) | 16.0 K (10.6 K - 21.9 K) | 96.9 K (40.2 K - 188 K) | 1.12 K (594 - 1.94 K) | 0 (0 - 0) | 72.8 K (35.6 K - 132 K) |
| 25-34 | 850 K (761 K - 935 K) | 164 K (85.5 K - 281 K) | 19.6 K (13.1 K - 26.4 K) | 46.6 K (19.9 K - 92.1 K) | 1.40 K (729 - 2.36 K) | 0 (0 - 0) | 64.0 K (32.6 K - 108 K) |
| 35-49 | 639 K (576 K - 702 K) | 123 K (63.5 K - 212 K) | 19.0 K (12.9 K - 26.3 K) | 34.7 K (14.4 K - 67.9 K) | 2.22 K (1.35 K - 3.39 K) | 0 (0 - 0) | 72.3 K (43.0 K - 111 K) |
| 50+ | 336 K (301 K - 369 K) | 64.7 K (33.6 K - 111 K) | 14.3 K (9.71 K - 19.7 K) | 17.5 K (7.08 K - 35.1 K) | 3.71 K (2.22 K - 5.53 K) | 0 (0 - 0) | 61.7 K (37.0 K - 91.9 K) |
| All | 8.68 M (8.19 M - 9.15 M) | 1.67 M (884 K - 2.86 M) | 80.3 K (54.2 K - 108 K) | 477 K (204 K - 933 K) | 9.66 K (6.22 K - 14.2 K) | 1.60 K (710 - 2.56 K) | 401 K (228 K - 672 K) |
| *Mean (95% uncertainty interval) cumulative totals per 100,000 person-years* | | | | | | | |
| <2 | 1.54 K (1.32 K - 1.77 K) | 296 (161 - 507) | 1.83 (1.17 - 2.72) | 84.9 (35.9 - 165) | 1.39 (0.56 - 2.39) | 2.24 (0.99 - 3.57) | 97.4 (42.2 - 170) |
| 2-14 | 968 (911 - 1.03 K) | 186 (99.3 - 313) | 2.43 (1.63 - 3.28) | 53.5 (22.7 - 105) | 0.05 (0 - 0.14) | 0 (0 - 0) | 14.6 (3.77 - 38.1) |
| 15-24 | 641 (584 - 696) | 123 (63.6 - 210) | 5.84 (3.87 - 7.99) | 35.3 (14.7 - 68.6) | 0.41 (0.22 - 0.70) | 0 (0 - 0) | 26.5 (13.0 - 47.9) |
| 25-34 | 425 (381 - 468) | 82.0 (42.8 - 141) | 9.78 (6.54 - 13.2) | 23.3 (9.96 - 46.1) | 0.70 (0.36 - 1.18) | 0 (0 - 0) | 32.0 (16.3 - 53.9) |
| 35-49 | 348 (313 - 382) | 67.0 (34.5 - 115) | 10.3 (7.01 - 14.3) | 18.9 (7.81 - 36.9) | 1.21 (0.73 - 1.84) | 0 (0 - 0) | 39.3 (23.4 - 60.4) |
| 50+ | 232 (207 - 255) | 44.7 (23.2 - 76.3) | 9.90 (6.70 - 13.6) | 12.1 (4.89 - 24.2) | 2.56 (1.53 - 3.82) | 0 (0 - 0) | 42.6 (25.5 - 63.4) |
| All | 674 (636 - 711) | 130 (68.6 - 222) | 6.23 (4.21 - 8.42) | 37.1 (15.8 - 72.4) | 0.75 (0.48 - 1.10) | 0.12 (0.06 - 0.20) | 31.1 (17.7 - 52.2) |

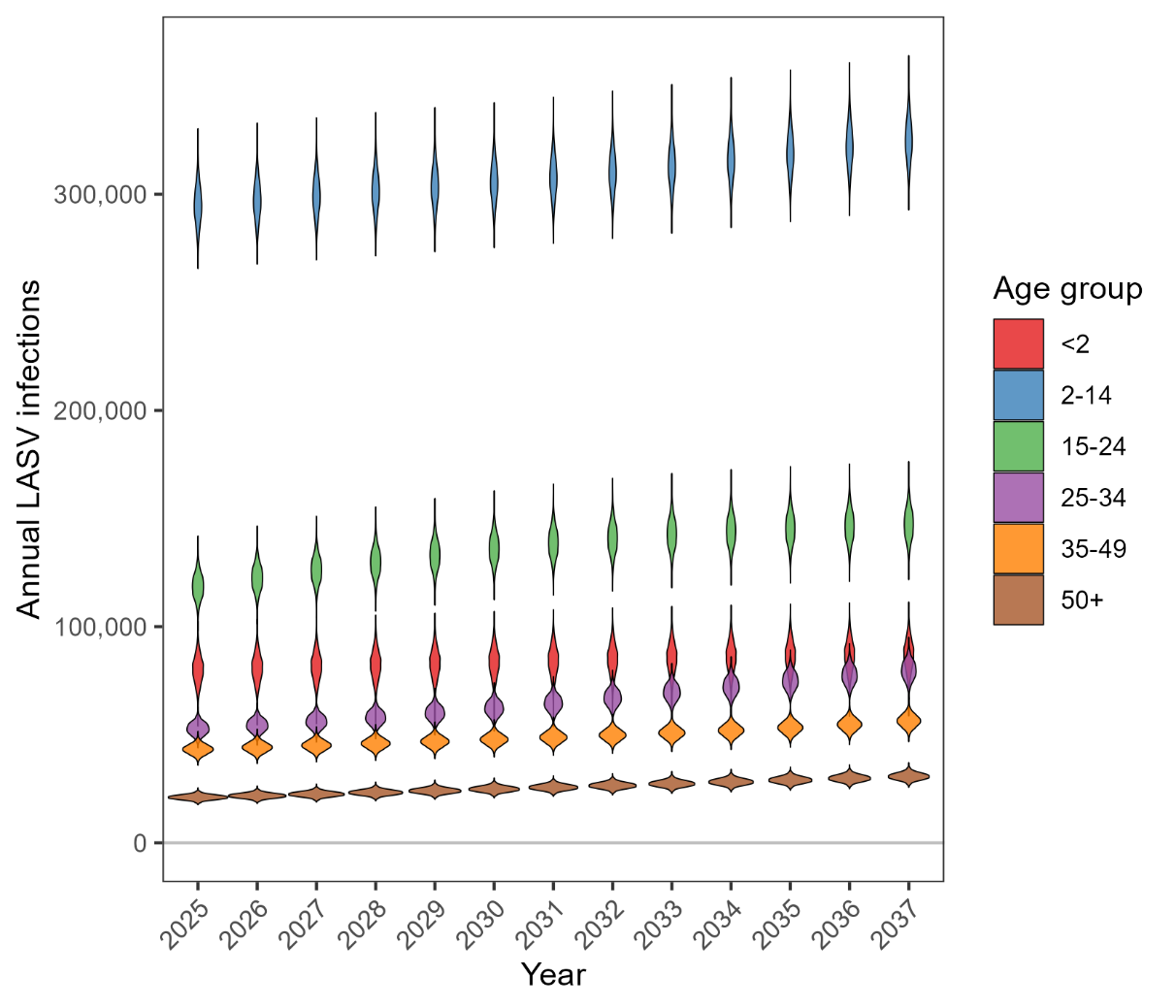

***Figure S18.*** *Density distributions (violins) of 500 Monte Carlo draws of the projected annual number of human LASV infections occurring in each age group, summed across all 19 areas.*

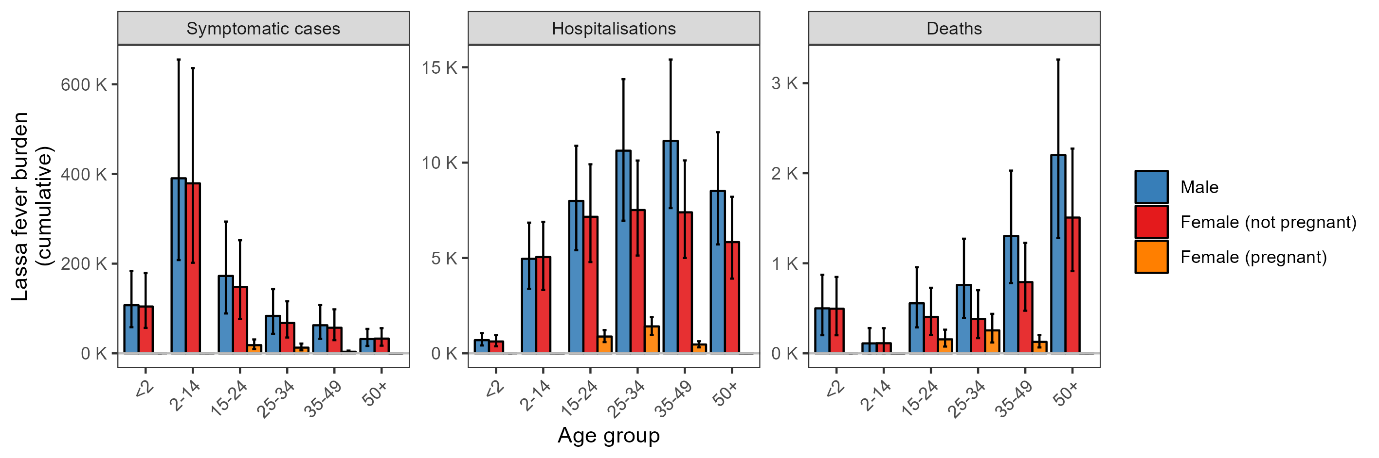

***Figure S19.*** *Cumulative Lassa fever cases, hospitalisations and deaths from 2025-2037, stratified by age group, sex and pregnancy status upon outcome onset. Error bars represent 95% uncertainty intervals.*

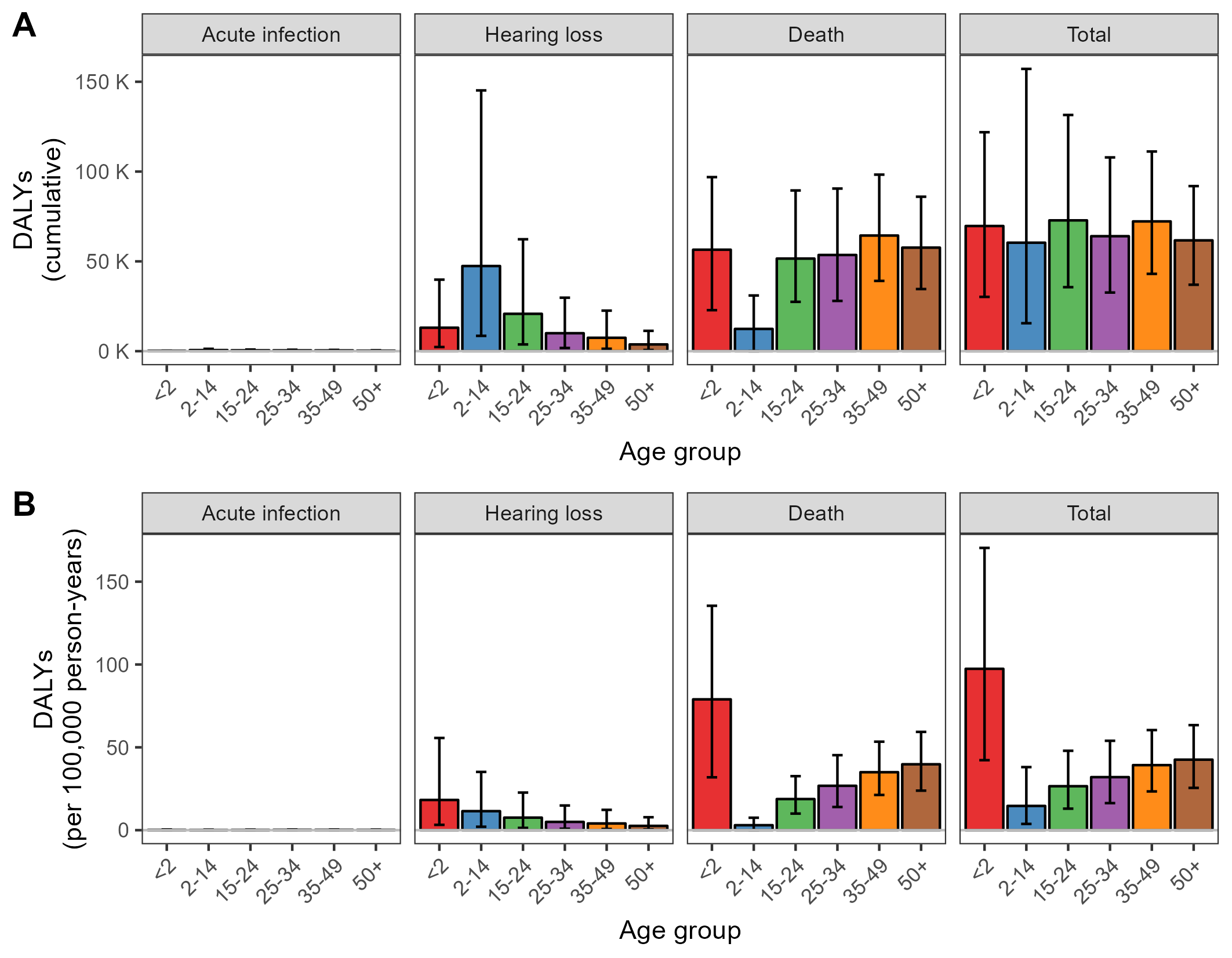

***Figure S20.*** *Cumulative DALYs due to Lassa fever from 2025-2037, including DALYs due to acute infection (including both mild/moderate disease and severe disease), DALYs due to sensorineural hearing loss and DALYs due to death. (****A****) Cumulative totals, stratified by the age of the individual when the infection occurs. (****B****) Cumulative totals per 100,000 person-years, stratified by the age of the individual when the infection occurs. Error bars represent 95% uncertainty intervals. DALY = disability-adjusted life-year.*

***Table S9.*** *Projected cumulative total societal costs of Lassa fever from 2025 to 2037 in the absence of vaccination, disaggregated by types of cost, and comparing estimates with (left) and without (right) discounting of future costs and life-years. DALYs do not include DALYs due to foetal loss. DALY = disability-adjusted life-year, K = thousand, M = million, B = billion.*

|  | **Mean (95% uncertainty interval) cumulative totals, discounted at 3.5%/year** | **Mean (95% uncertainty interval) cumulative totals, not discounted** |
| --- | --- | --- |
| **Healthcare costs (I$ 2023)** | | |
| Outpatient (reimbursed) | 3.44 M (575 K - 8.89 M) | 4.22 M (706 K - 10.9 M) |
| Outpatient (out-of-pocket) | 14.2 M (7.21 M - 23.9 M) | 17.4 M (8.84 M - 29.4 M) |
| Inpatient (reimbursed) | 65.7 M (44.4 M - 88.8 M) | 81.0 M (54.7 M - 109 M) |
| Inpatient (out-of-pocket) | 93.6 M (63.2 M - 126 M) | 115 M (77.9 M - 156 M) |
| Total | 177 M (116 M - 247 M) | 218 M (143 M - 305 M) |
| **Productivity losses (I$ 2023)** | | |
| Mild/moderate disease | 14.1 M (6.41 M - 26.2 M) | 17.5 M (7.91 M - 32.3 M) |
| Severe disease | 11.6 M (7.55 M - 16.4 M) | 14.4 M (9.33 M - 20.3 M) |
| Hearing loss | 178 M (53.8 M - 433 M) | 226 M (67.2 M - 556 M) |
| Death | 502 M (306 M - 767 M) | 1.02 B (615 M - 1.58 B) |
| Total | 706 M (402 M - 1.14 B) | 1.28 B (736 M - 1.99 B) |
| **Monetised DALYs (I$ 2023)** | | |
| Mild/moderate disease | 126 K (33.9 K - 313 K) | 155 K (41.5 K - 385 K) |
| Severe disease | 225 K (126 K - 360 K) | 278 K (155 K - 444 K) |
| Hearing loss | 14.6 M (2.69 M - 43.9 M) | 18.7 M (3.39 M - 56.5 M) |
| Death | 24.9 M (15.5 M - 36.6 M) | 54.1 M (33.3 M - 79.2 M) |
| Total | 39.9 M (21.9 M - 74.2 M) | 73.3 M (41.5 M - 123 M) |

***4b. Vaccine impact***

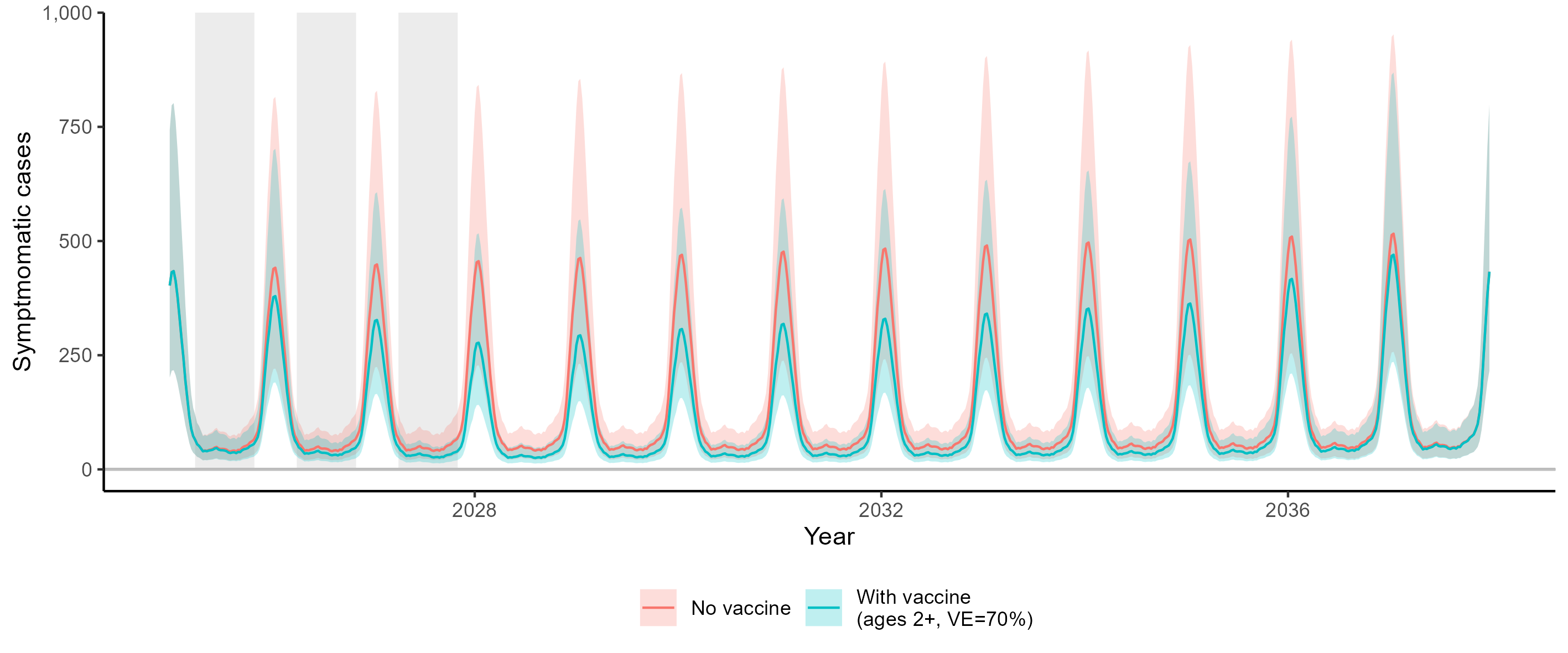

***Figure S21.*** *Total weekly symptomatic Lassa fever cases in an exemplar area (Edo, Nigeria) from 1 January 2025 to 31 December 2037, with (blue) and without (red) an untargeted 3-year vaccination campaign (i.e. vaccinating the whole population aged 2+) and assuming 70% vaccine efficacy against all symptomatic disease. Curved lines represent means, shading represents 95% uncertainty intervals, and grey vertical bars represent periods of vaccine distribution. VE = vaccine efficacy.*

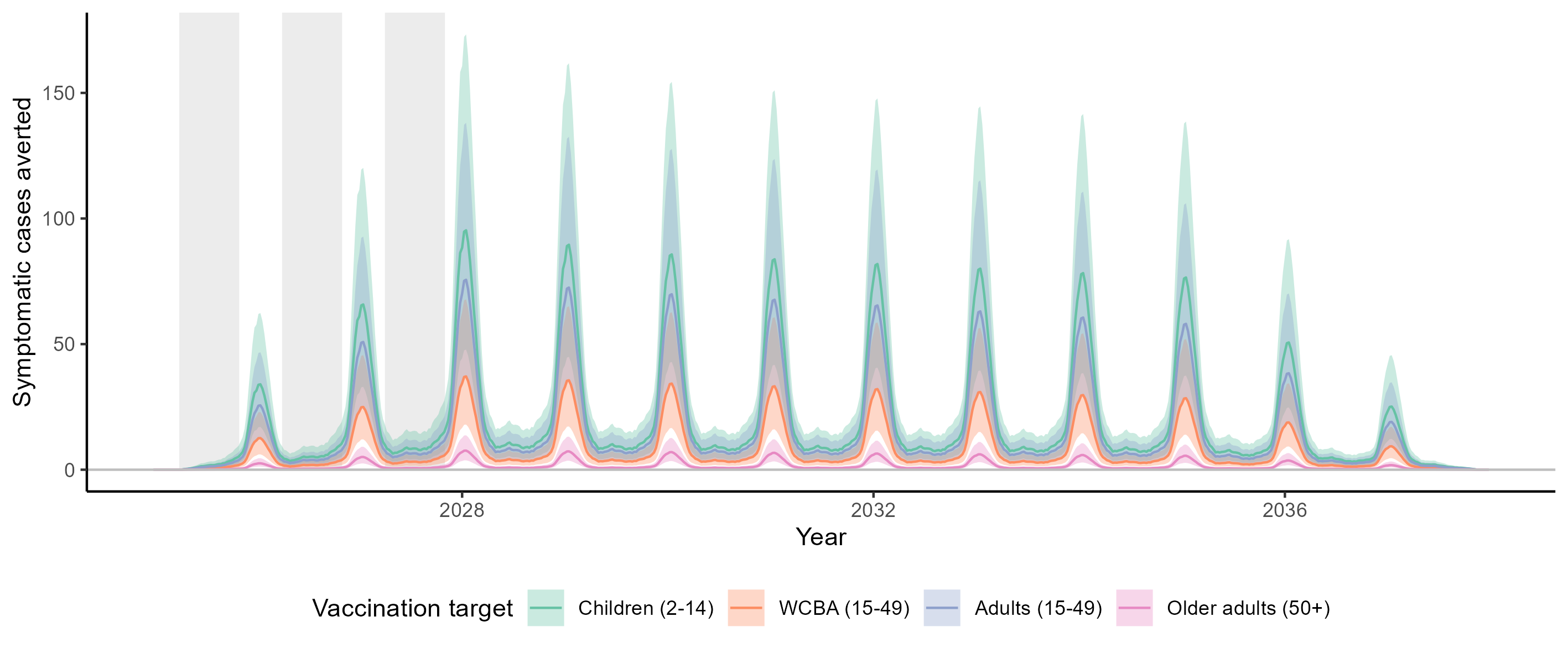

***Figure S22.*** *Total weekly symptomatic Lassa fever cases averted by a 3-year vaccination campaign in an exemplar area (Edo, Nigeria) from 1 January 2025 to 31 December 2037, varying the group targeted by vaccination (colours) and assuming 70% vaccine efficacy against all symptomatic disease. Curved lines represent means, shading represents 95% uncertainty intervals, and grey vertical bars represent periods of vaccine distribution. The target group “adults” includes adolescents and adults. WCBA = women of childbearing age.*

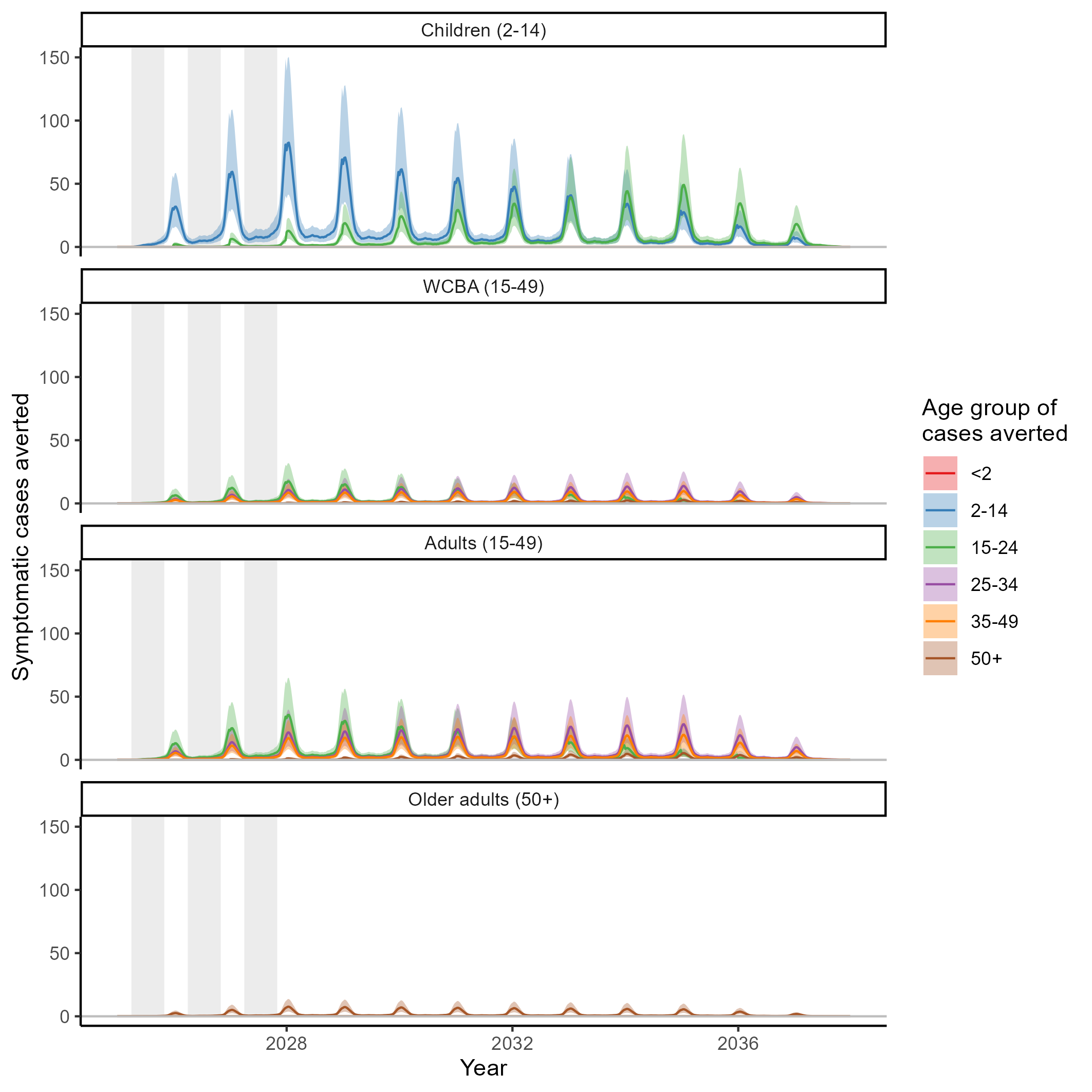

***Figure S23.*** *Weekly symptomatic Lassa fever cases in different age groups (colours) that are averted by vaccination in an exemplar area (Edo, Nigeria) from 1 January 2025 to 31 December 2037, varying the group targeted by vaccination (panels) and assuming 70% vaccine efficacy against all symptomatic disease. Curved lines represent means, shading represents 95% uncertainty intervals, and grey vertical bars represent periods of vaccine distribution. The target group “adults” includes adolescents and adults.*

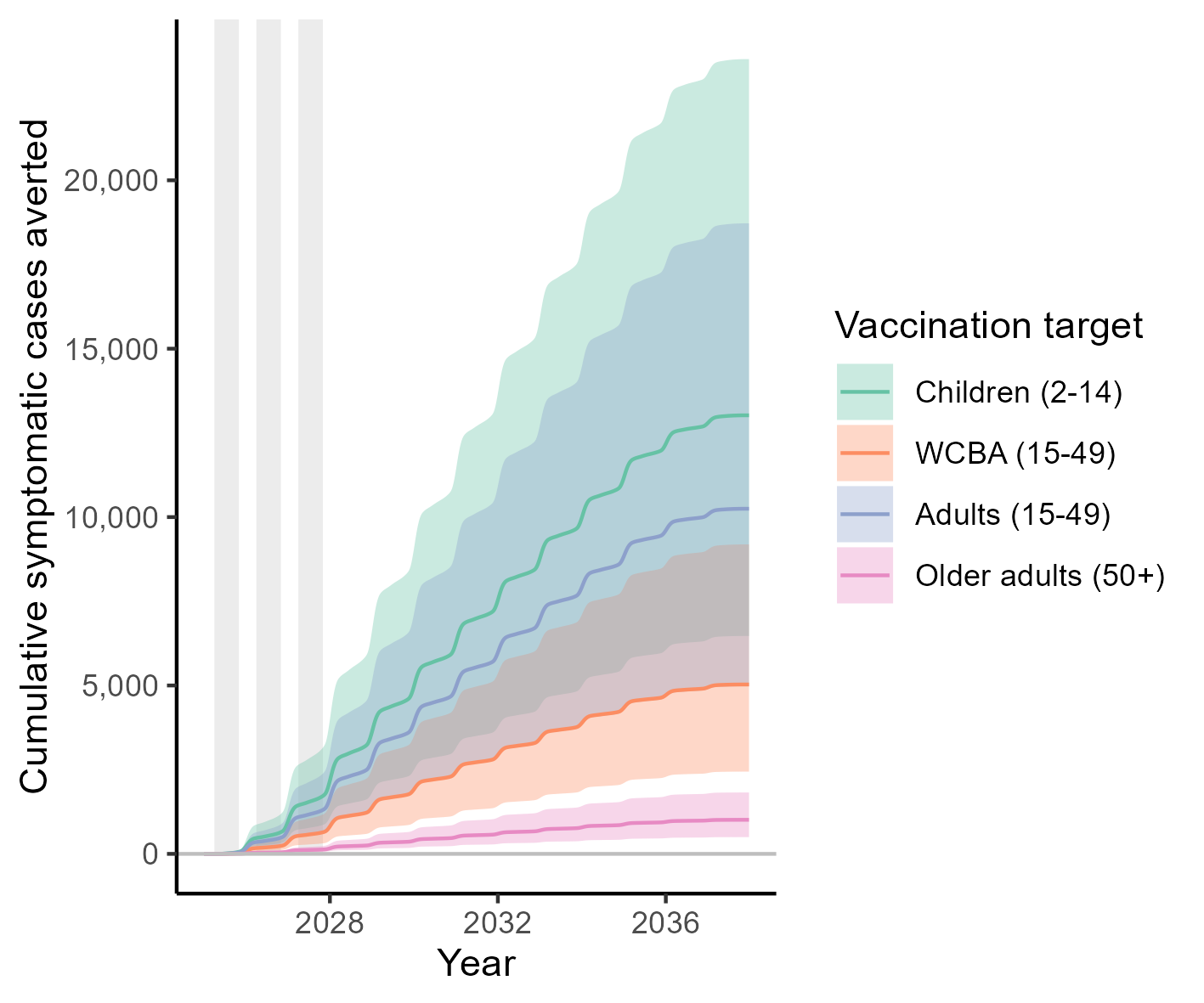

***Figure S24.*** *The weekly total cumulative number of symptomatic Lassa fever cases averted due to vaccination in one exemplar area (Edo, Nigeria) from 1 January 2025 to 31 December 2037 when targeting different groups for vaccination (colours). Grey vertical bars indicate periods of Lassa vaccine distribution. Curved lines and shaded areas represent means and 95% uncertainty intervals, respectively, for a vaccine 70% effective against all symptomatic disease. The target group “adults” includes adolescents and adults. WCBA = women of childbearing age.*

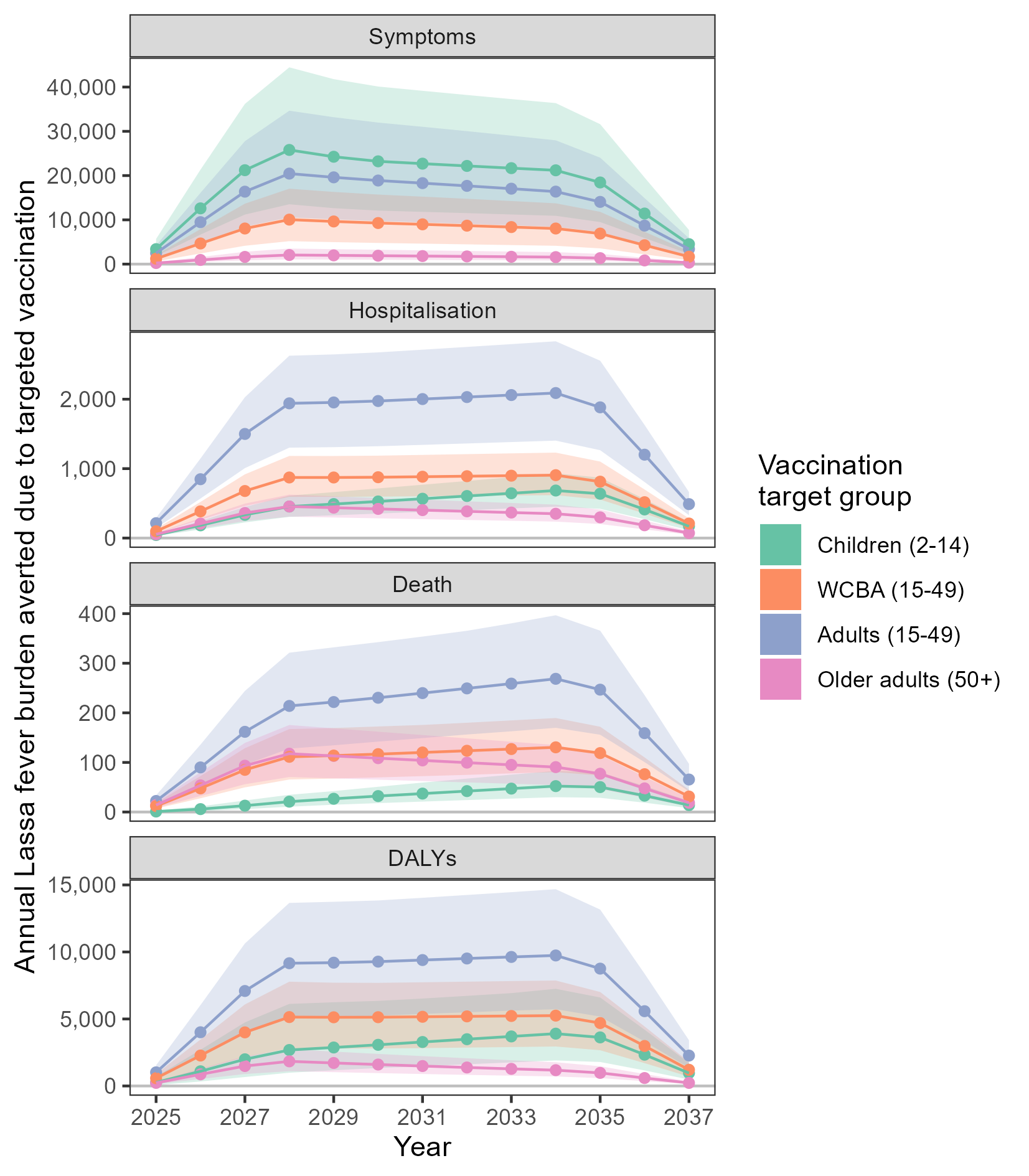

***Figure S25.*** *Change through time in annual projections of the total number of Lassa fever cases, hospitalisations, deaths and DALYs averted when vaccinating different target groups (colours). These numbers represent total outcomes averted each year summed across all 19 areas when assuming 70% vaccine efficacy against all symptomatic disease. Lines represent means, shading represents 95% uncertainty intervals. WCBA = women of childbearing age.*

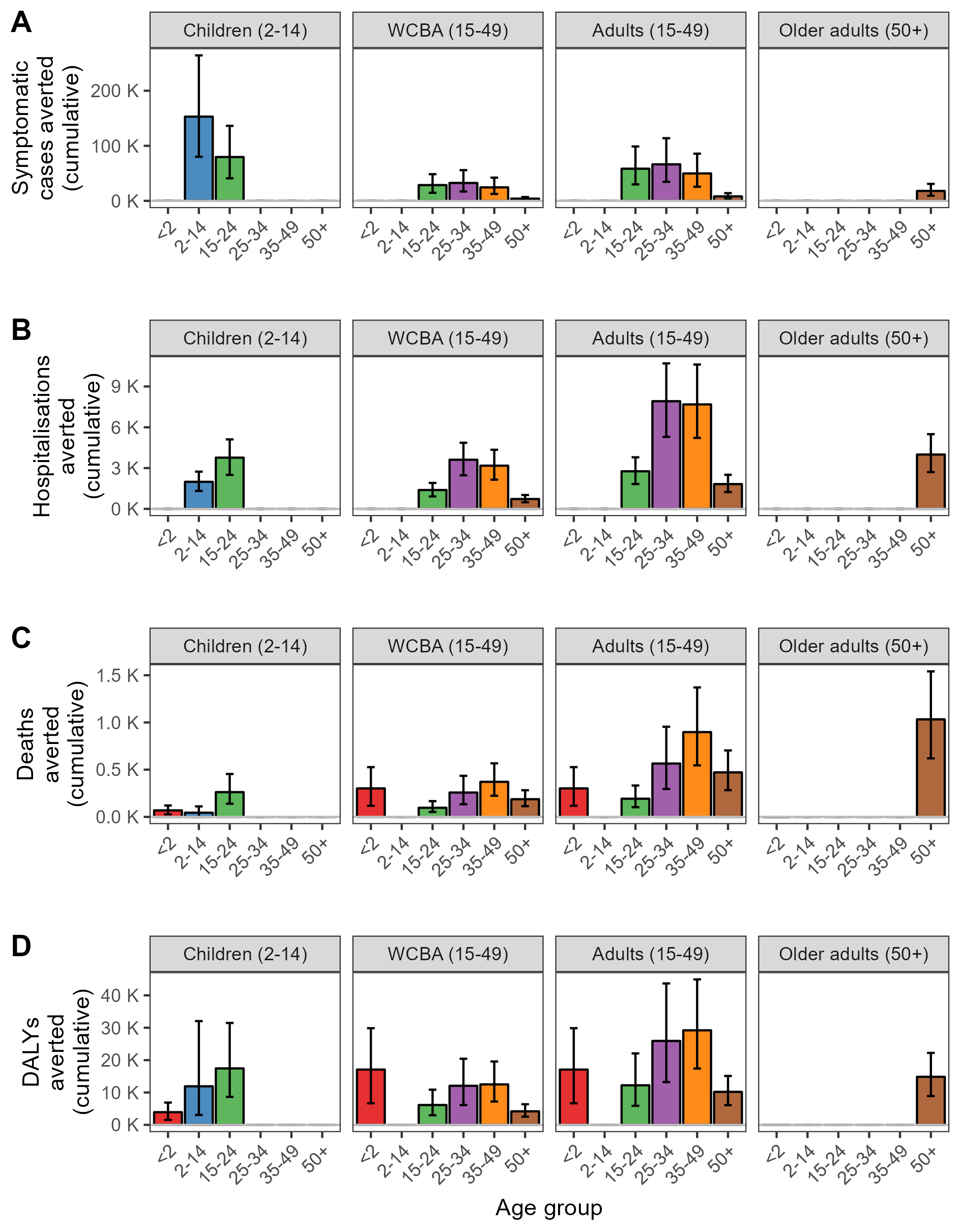

***Figure S26.*** *Age distributions of cumulative Lassa fever health outcomes averted due to targeted Lassa vaccination for a vaccine having 70% efficacy against all symptomatic disease. Cumulative symptomatic Lassa fever cases (****A****), hospitalisations (****B****), deaths (****C****) and DALYs (****D****) averted from 2025-2037 when vaccinating different target groups (panels), where results are stratified by the age groups in which the outcomes are averted (x-axis, colours). DALYs do not include foetal loss DALYs. Error bars represent 95% uncertainty intervals. DALY = disability-adjusted life-year, WCBA = women of childbearing age.*

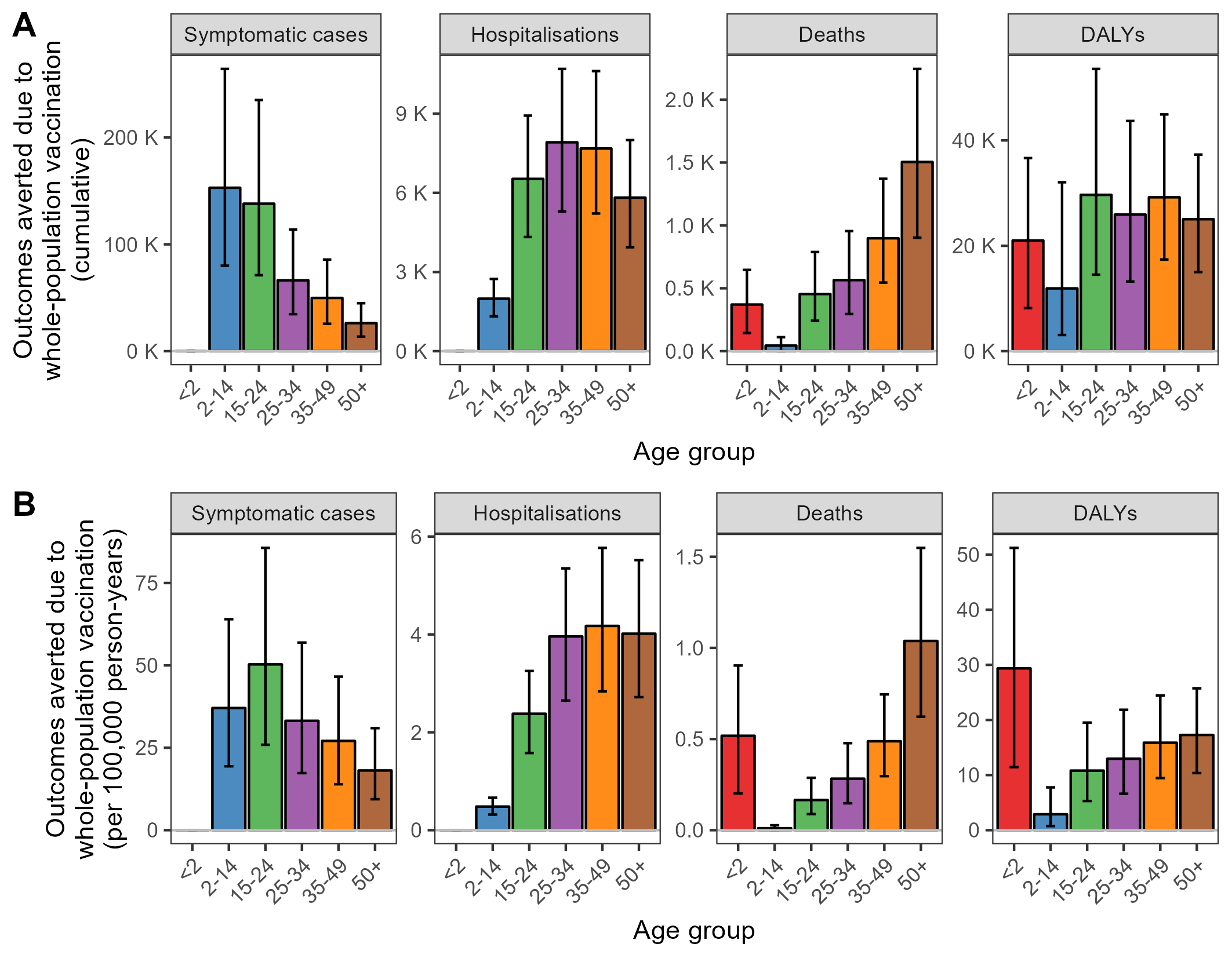

***Figure S27.*** *Age distributions of cumulative Lassa fever health outcomes averted due to untargeted whole-population vaccination (all individuals aged 2+) with a 1-dose Lassa fever vaccine having 70% efficacy against all symptomatic disease. (****A****) Impact: cumulative total health outcomes averted due to vaccination summed across all 19 areas from 2025 through to 2037. (****B****) Efficiency: cumulative total health outcomes averted per 100,000 doses of vaccine administered. DALYs exclude DALYs due to foetal loss. Error bars represent 95% uncertainty intervals. WCBA, women of childbearing age.*

***Table S10.*** *Projected cumulative total Lassa fever health outcomes averted due to Lassa vaccination from 2025 to 2037, depending on the group targeted for vaccination and the vaccine’s efficacy against mild/moderate and severe disease. These projections correspond to base case assumptions: DALYs do not include foetal loss DALYs, symptom risk is not associated with age, and all survivors of symptomatic infection are at risk of developing sensorineural hearing less, regardless of symptom severity. DALY = disability-adjusted life-year, WCBA = women of childbearing age.*

| **Outcome averted** | **Vaccine target group** | **Vaccine efficacy** | | | | |
| --- | --- | --- | --- | --- | --- | --- |
|  |  | **50% mild/moderate, 50% severe** | **50% mild/moderate, 70% severe** | **70% mild/moderate, 70% severe** | **70% mild/moderate, 90% severe** | **90% mild/moderate, 90% severe** |
| Symptomatic Lassa fever | Children (2-14) | 166 K (86.1 K - 287 K) | 168 K (87.2 K - 289 K) | 233 K (121 K - 401 K) | 234 K (122 K - 404 K) | 299 K (155 K - 516 K) |
|  | WCBA (15-49) | 64.2 K (33.0 K - 109 K) | 66.7 K (34.7 K - 112 K) | 89.8 K (46.2 K - 153 K) | 92.4 K (47.9 K - 156 K) | 115 K (59.4 K - 196 K) |
|  | Adolescents and adults (15-49) | 131 K (67.2 K - 222 K) | 136 K (71.1 K - 230 K) | 183 K (94.1 K - 310 K) | 189 K (98.0 K - 318 K) | 235 K (121 K - 399 K) |
|  | Older adults (50+) | 12.9 K (6.69 K - 22.0 K) | 14 K (7.49 K - 23.6 K) | 18.1 K (9.36 K - 30.9 K) | 19.2 K (10.2 K - 32.4 K) | 23.2 K (12.0 K - 39.7 K) |
|  | All (2+) | 310 K (160 K - 531 K) | 318 K (166 K - 543 K) | 433 K (224 K - 743 K) | 442 K (230 K - 756 K) | 557 K (288 K - 956 K) |
| Hospitalisation | Children (2-14) | 4.11 K (2.74 K - 5.57 K) | 5.76 K (3.83 K - 7.80 K) | 5.76 K (3.83 K - 7.80 K) | 7.4 K (4.92 K - 10.0 K) | 7.4 K (4.92 K - 10.0 K) |
|  | WCBA (15-49) | 6.36 K (4.34 K - 8.63 K) | 8.90 K (6.07 K - 12.1 K) | 8.90 K (6.07 K - 12.1 K) | 11.4 K (7.81 K - 15.5 K) | 11.4 K (7.81 K - 15.5 K) |
|  | Adolescents and adults (15-49) | 14.4 K (9.67 K - 19.5 K) | 20.2 K (13.5 K - 27.4 K) | 20.2 K (13.5 K - 27.4 K) | 26.0 K (17.4 K - 35.2 K) | 26.0 K (17.4 K - 35.2 K) |
|  | Older adults (50+) | 2.85 K (1.93 K - 3.92 K) | 4.00 K (2.71 K - 5.49 K) | 4.00 K (2.71 K - 5.49 K) | 5.14 K (3.48 K - 7.06 K) | 5.14 K (3.48 K - 7.06 K) |
|  | All (2+) | 21.4 K (14.4 K - 28.9 K) | 29.9 K (20.2 K - 40.4 K) | 29.9 K (20.2 K - 40.4 K) | 38.5 K (25.9 K - 51.9 K) | 38.5 K (25.9 K - 51.9 K) |
| Sensorineural hearing loss | Children (2-14) | 47.6 K (20.1 K - 91.2 K) | 48.0 K (20.3 K - 91.9 K) | 66.6 K (28.1 K - 128 K) | 67.1 K (28.4 K - 128 K) | 85.7 K (36.2 K - 164 K) |
|  | WCBA (15-49) | 18.2 K (7.62 K - 36.3 K) | 18.9 K (7.98 K - 37.3 K) | 25.5 K (10.7 K - 50.8 K) | 26.2 K (11.0 K - 51.8 K) | 32.8 K (13.7 K - 65.4 K) |
|  | Adolescents and adults (15-49) | 37.0 K (15.5 K - 73.8 K) | 38.5 K (16.3 K - 76.0 K) | 51.8 K (21.7 K - 103 K) | 53.3 K (22.5 K - 105 K) | 66.6 K (27.9 K - 133 K) |
|  | Older adults (50+) | 3.49 K (1.41 K – 7.00 K) | 3.73 K (1.54 K - 7.37 K) | 4.89 K (1.98 K - 9.80 K) | 5.13 K (2.10 K - 10.2 K) | 6.28 K (2.54 K - 12.6 K) |
|  | All (2+) | 88.1 K (37.2 K - 172 K) | 90.3 K (38.3 K - 175 K) | 123 K (52.0 K - 241 K) | 126 K (53.2 K - 244 K) | 159 K (66.9 K - 310 K) |
| Death | Children (2-14) | 268 (152 - 427) | 375 (212 - 598) | 375 (212 - 598) | 483 (273 - 769) | 483 (273 - 769) |
|  | WCBA (15-49) | 867 (523 - 1.27 K) | 1.21 K (732 - 1.77 K) | 1.21 K (732 - 1.77 K) | 1.56 K (941 - 2.28 K) | 1.56 K (941 - 2.28 K) |
|  | Adolescents and adults (15-49) | 1.73 K (1.08 K - 2.56 K) | 2.43 K (1.51 K - 3.58 K) | 2.43 K (1.51 K - 3.58 K) | 3.12 K (1.94 K - 4.60 K) | 3.12 K (1.94 K - 4.60 K) |
|  | Older adults (50+) | 738 (443 - 1.10 K) | 1.03 K (620 - 1.54 K) | 1.03 K (620 - 1.54 K) | 1.33 K (797 - 1.98 K) | 1.33 K (797 - 1.98 K) |
|  | All (2+) | 2.74 K (1.75 K - 4.07 K) | 3.84 K (2.46 K - 5.69 K) | 3.84 K (2.46 K - 5.69 K) | 4.93 K (3.16 K - 7.32 K) | 4.93 K (3.16 K - 7.32 K) |
| Foetal loss | Children (2-14) | 86.1 (38.5 - 138) | 121 (53.9 - 193) | 121 (53.9 - 193) | 155 (69.3 - 249) | 155 (69.3 - 249) |
|  | WCBA (15-49) | 378 (168 - 604) | 529 (236 - 845) | 529 (236 - 845) | 680 (303 - 1.09 K) | 680 (303 - 1.09 K) |
|  | Adolescents and adults (15-49) | 378 (168 - 604) | 529 (236 - 845) | 529 (236 - 845) | 680 (303 - 1.09 K) | 680 (303 - 1.09 K) |
|  | Older adults (50+) | 0.21 (0.10 - 0.34) | 0.30 (0.13 - 0.47) | 0.30 (0.13 - 0.47) | 0.38 (0.17 - 0.60) | 0.38 (0.17 - 0.60) |
|  | All (2+) | 464 (206 - 740) | 650 (288 - 1.04 K) | 650 (288 - 1.04 K) | 835 (370 - 1.33 K) | 835 (370 - 1.33 K) |
| DALYs | Children (2-14) | 23.8 K (10.6 K - 47.5 K) | 29.2 K (14.1 K - 54.3 K) | 33.3 K (14.8 K - 66.5 K) | 38.8 K (18.3 K - 73.1 K) | 42.8 K (19.1 K - 85.6 K) |
|  | WCBA (15-49) | 37.1 K (20.3 K - 55.7 K) | 50.5 K (28.1 K - 75.8 K) | 52 K (28.5 K - 77.9 K) | 65.4 K (36.2 K - 97.9 K) | 66.8 K (36.6 K - 100 K) |
|  | Adolescents and adults (15-49) | 67.6 K (38.7 K - 101 K) | 91.7 K (53.1 K - 138 K) | 94.6 K (54.2 K - 141 K) | 119 K (68.5 K - 179 K) | 122 K (69.7 K - 182 K) |
|  | Older adults (50+) | 10.6 K (6.37 K - 15.9 K) | 14.6 K (8.77 K - 21.7 K) | 14.9 K (8.92 K - 22.2 K) | 18.8 K (11.3 K - 28 K) | 19.1 K (11.5 K - 28.6 K) |
|  | All (2+) | 102 K (58.5 K - 162 K) | 136 K (78 K - 203 K) | 143 K (81.8 K - 227 K) | 176 K (101 K - 268 K) | 184 K (105 K - 292 K) |

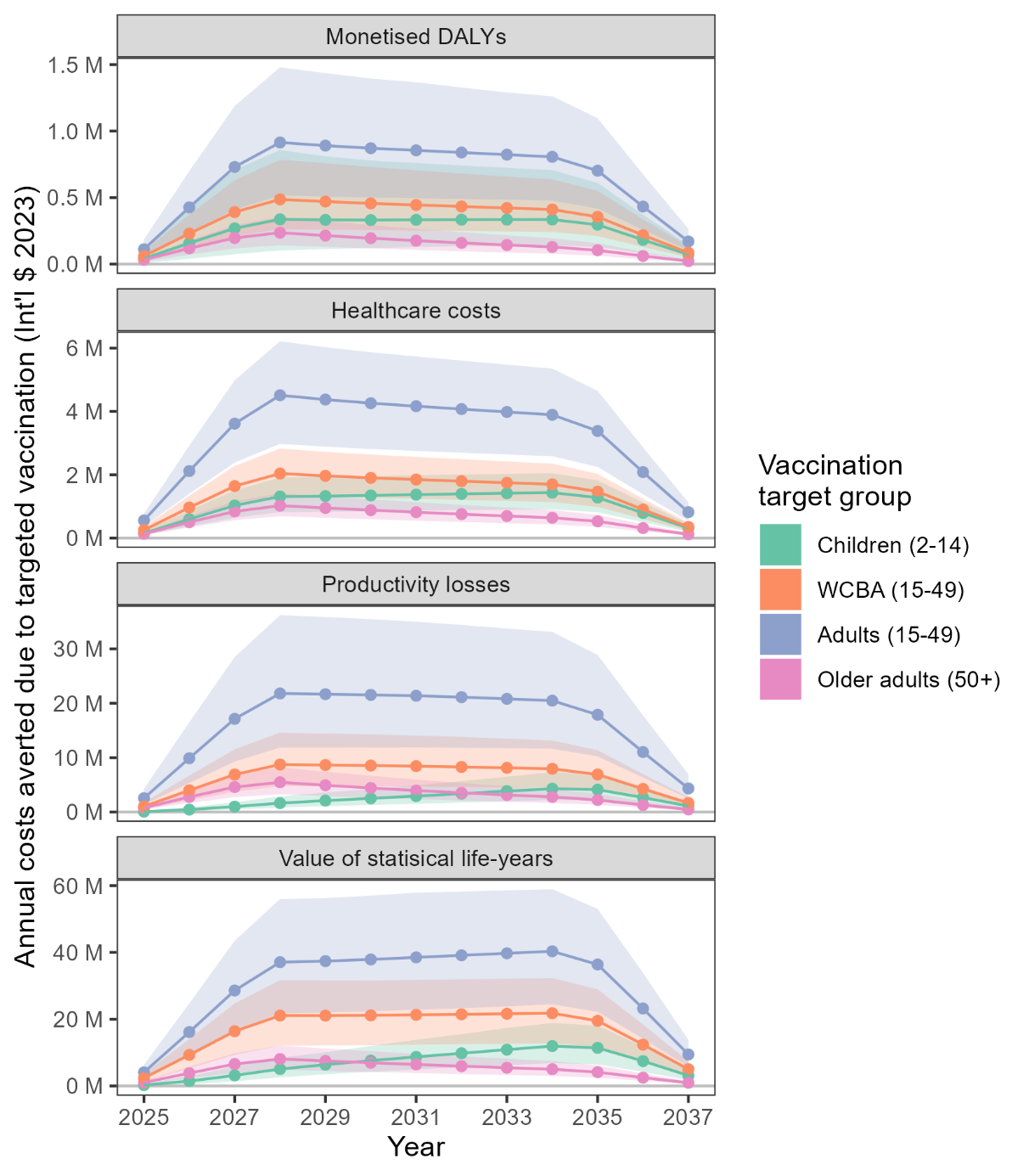

***Figure S28.*** *Change through time in annual projections of monetary costs averted (panels) when vaccinating different target groups (colours). These numbers represent total costs averted each year combined across all 19 areas when assuming 70% vaccine efficacy against all symptomatic disease. Monetised DALYs do not include foetal loss DALYs. Future costs and life-years are discounted at 3.5%. Lines represent means, shading represents 95% uncertainty intervals. DALY = disability-adjusted life-year, WCBA = women of childbearing age.*

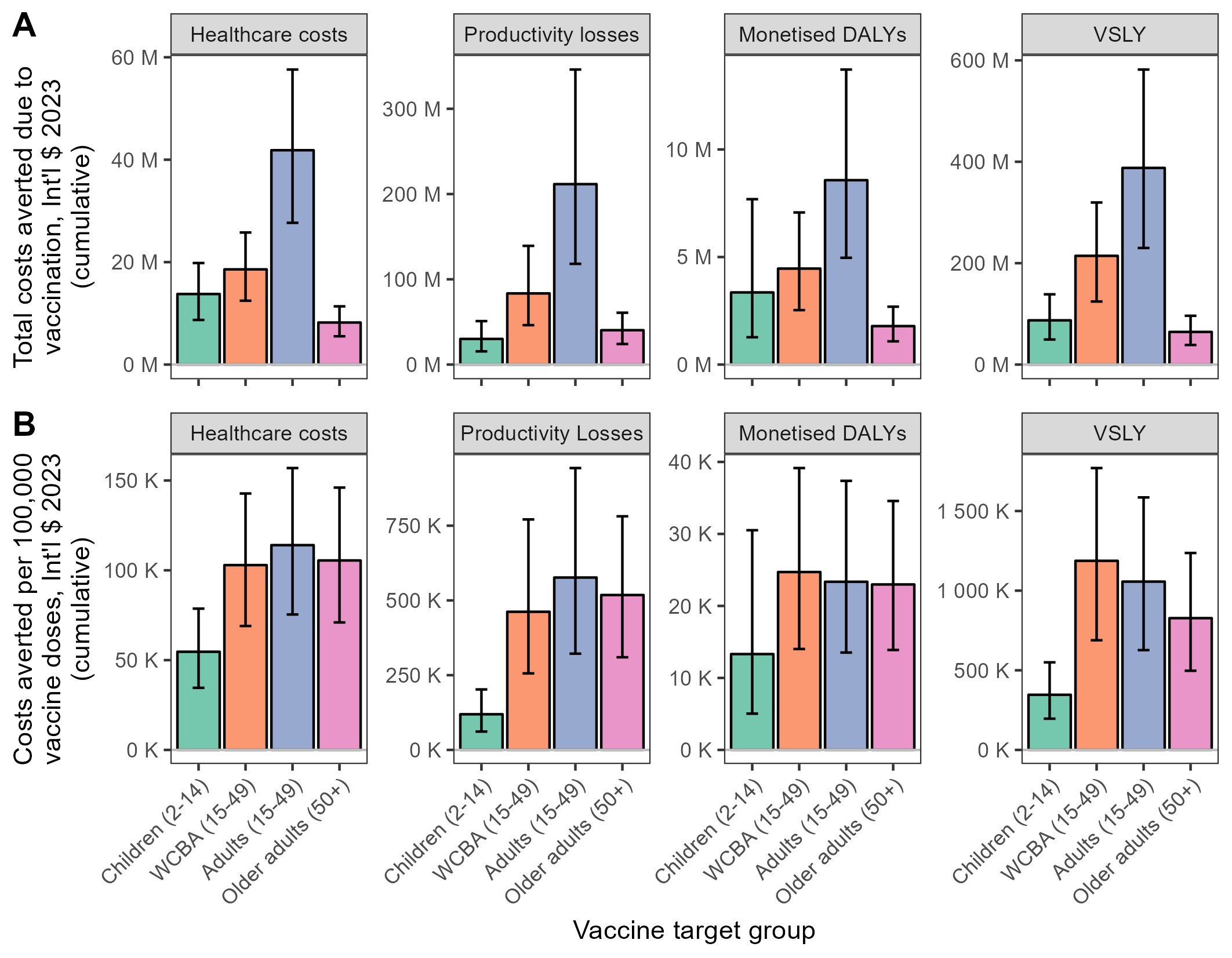

***Figure S29.*** *Comparing the cumulative economic impacts and efficiencies of targeting different risk groups for vaccination with a 1-dose Lassa fever vaccine having 70% efficacy against both mild and severe symptomatic disease. (****A****) Impact: cumulative total economic costs averted due to vaccination across all 19 areas from 2025 through to 2037. (****B****) Efficiency: cumulative total economic costs averted per 100,000 doses of vaccine administered. Monetised DALYs do not include foetal loss DALYs. Future costs and life-years are discounted at 3.5%. Error bars represent 95% uncertainty intervals. WCBA = women of childbearing age.*

***Table S11****. Projected cumulative total healthcare costs averted due to Lassa vaccination from 2025 to 2037, depending on the group targeted for vaccination and the vaccine’s efficacy against disease. Future costs and life-years are discounted annually at 3.5%/year. WCBA = women of childbearing age, I$ = International dollar, K = thousand, M = million.*

| **Costs averted** | **Vaccine target group** | **Vaccine efficacy** | | | | |
| --- | --- | --- | --- | --- | --- | --- |
|  |  | **50% mild/moderate, 50% severe** | **50% mild/moderate, 70% severe** | **70% mild/moderate, 70% severe** | **70% mild/moderate, 90% severe** | **90% mild/moderate, 90% severe** |
| Outpatient healthcare costs, reimbursed (I$ 2023) | Children (2-14) | 1.46 M (731 K - 2.43 M) | 1.46 M (731 K - 2.43 M) | 2.04 M (1.02 M - 3.41 M) | 2.04 M (1.02 M - 3.41 M) | 2.62 M (1.32 M - 4.38 M) |
|  | WCBA (15-49) | 519 K (256 K - 886 K) | 519 K (256 K - 886 K) | 727 K (358 K - 1.24 M) | 727 K (358 K - 1.24 M) | 935 K (461 K - 1.59 M) |
|  | Adolescents and adults (15-49) | 1.04 M (513 K - 1.78 M) | 1.04 M (513 K - 1.78 M) | 1.46 M (718 K - 2.50 M) | 1.46 M (718 K - 2.50 M) | 1.88 M (924 K - 3.21 M) |
|  | Older adults (50+) | 90.2 K (40.9 K - 159 K) | 90.2 K (40.9 K - 159 K) | 126 K (57.3 K - 223 K) | 126 K (57.3 K - 223 K) | 162 K (73.7 K - 286 K) |
|  | All (2+) | 2.59 M (1.29 M - 4.37 M) | 2.59 M (1.29 M - 4.37 M) | 3.63 M (1.80 M - 6.12 M) | 3.63 M (1.80 M - 6.12 M) | 4.67 M (2.32 M - 7.87 M) |
| Outpatient healthcare costs, out-of-pocket  (I$ 2023) | Children (2-14) | 353 K (58.9 K - 938 K) | 353 K (58.9 K - 938 K) | 494 K (82.5 K - 1.31 M) | 494 K (82.5 K - 1.31 M) | 636 K (106 K - 1.69 M) |
|  | WCBA (15-49) | 126 K (21.1 K - 341 K) | 126 K (21.1 K - 341 K) | 177 K (29.5 K - 478 K) | 177 K (29.5 K - 478 K) | 227 K (37.9 K - 614 K) |
|  | Adolescents and adults (15-49) | 254 K (42.3 K - 688 K) | 254 K (42.3 K - 688 K) | 356 K (59.2 K - 963 K) | 356 K (59.2 K - 963 K) | 458 K (76.1 K - 1.24 M) |
|  | Older adults (50+) | 22.1 K (3.36 K - 58.7 K) | 22.1 K (3.36 K - 58.7 K) | 30.9 K (4.71 K - 82.2 K) | 30.9 K (4.71 K - 82.2 K) | 39.7 K (6.06 K - 106 K) |
|  | All (2+) | 629 K (105 K - 1.69 M) | 629 K (105 K - 1.69 M) | 881 K (147 K - 2.37 M) | 881 K (147 K - 2.37 M) | 1.13 M (189 K - 3.04 M) |
| Inpatient healthcare costs, reimbursed (I$ 2023) | Children (2-14) | 4.71 M (3.14 M - 6.38 M) | 6.60 M (4.40 M - 8.94 M) | 6.60 M (4.40 M - 8.94 M) | 8.48 M (5.65 M - 11.5 M) | 8.48 M (5.65 M - 11.5 M) |
|  | WCBA (15-49) | 7.43 M (5.06 M - 10.1 M) | 10.4 M (7.09 M - 14.1 M) | 10.4 M (7.09 M - 14.1 M) | 13.4 M (9.12 M - 18.1 M) | 13.4 M (9.12 M - 18.1 M) |
|  | Adolescents and adults (15-49) | 16.8 M (11.3 M - 22.8 M) | 23.5 M (15.8 M - 31.9 M) | 23.5 M (15.8 M - 31.9 M) | 30.3 M (20.3 M – 41.0 M) | 30.3 M (20.3 M – 41.0 M) |
|  | Older adults (50+) | 3.38 M (2.29 M - 4.65 M) | 4.74 M (3.21 M - 6.51 M) | 4.74 M (3.21 M - 6.51 M) | 6.09 M (4.12 M - 8.37 M) | 6.09 M (4.12 M - 8.37 M) |
|  | All (2+) | 24.9 M (16.8 M - 33.6 M) | 34.9 M (23.5 M - 47.1 M) | 34.9 M (23.5 M - 47.1 M) | 44.8 M (30.2 M - 60.5 M) | 44.8 M (30.2 M - 60.5 M) |
| Inpatient healthcare costs, out-of-pocket (I$ 2023) | Children (2-14) | 3.31 M (2.21 M - 4.49 M) | 4.64 M (3.09 M - 6.28 M) | 4.64 M (3.09 M - 6.28 M) | 5.96 M (3.98 M - 8.08 M) | 5.96 M (3.98 M - 8.08 M) |
|  | WCBA (15-49) | 5.20 M (3.55 M - 7.08 M) | 7.28 M (4.97 M - 9.91 M) | 7.28 M (4.97 M - 9.91 M) | 9.37 M (6.39 M - 12.7 M) | 9.37 M (6.39 M - 12.7 M) |
|  | Adolescents and adults (15-49) | 11.8 M (7.91 M – 16.0 M) | 16.5 M (11.1 M - 22.4 M) | 16.5 M (11.1 M - 22.4 M) | 21.2 M (14.2 M - 28.7 M) | 21.2 M (14.2 M - 28.7 M) |
|  | Older adults (50+) | 2.37 M (1.60 M - 3.26 M) | 3.32 M (2.24 M - 4.56 M) | 3.32 M (2.24 M - 4.56 M) | 4.26 M (2.89 M - 5.86 M) | 4.26 M (2.89 M - 5.86 M) |
|  | All (2+) | 17.5 M (11.8 M - 23.6 M) | 24.5 M (16.5 M – 33.0 M) | 24.5 M (16.5 M – 33.0 M) | 31.4 M (21.2 M - 42.4 M) | 31.4 M (21.2 M - 42.4 M) |

***Table S12****. Projected cumulative total productivity losses averted due to Lassa vaccination from 2025 to 2037, depending on the group targeted for vaccination and the vaccine’s efficacy against disease. Future costs and life-years are discounted annually at 3.5%/year. WCBA = women of childbearing age, I$ = International dollar, K = thousand, M = million, B = billion.*

|  | **Vaccine target group** | **Vaccine efficacy** | | | | |
| --- | --- | --- | --- | --- | --- | --- |
|  |  | **50% mild/moderate, 50% severe** | **50% mild/moderate, 70% severe** | **70% mild/moderate, 70% severe** | **70% mild/moderate, 90% severe** | **90% mild/moderate, 90% severe** |
| Productivity lost due to mild and moderate disease (I$ 2023) | Children (2-14) | 478 K (224 K - 869 K) | 478 K (224 K - 869 K) | 669 K (314 K - 1.22 M) | 669 K (314 K - 1.22 M) | 861 K (403 K - 1.56 M) |
|  | WCBA (15-49) | 1.44 M (661 K - 2.68 M) | 1.44 M (661 K - 2.68 M) | 2.02 M (925 K - 3.75 M) | 2.02 M (925 K - 3.75 M) | 2.60 M (1.19 M - 4.82 M) |
|  | Adolescents and adults (15-49) | 3.28 M (1.49 M - 6.11 M) | 3.28 M (1.49 M - 6.11 M) | 4.59 M (2.09 M - 8.55 M) | 4.59 M (2.09 M - 8.55 M) | 5.91 M (2.68 M – 11.0 M) |
|  | Older adults (50+) | 319 K (138 K - 611 K) | 319 K (138 K - 611 K) | 447 K (193 K - 855 K) | 447 K (193 K - 855 K) | 575 K (248 K - 1.10 M) |
|  | All (2+) | 4.08 M (1.85 M - 7.55 M) | 4.08 M (1.85 M - 7.55 M) | 5.71 M (2.59 M - 10.6 M) | 5.71 M (2.59 M - 10.6 M) | 7.34 M (3.33 M - 13.6 M) |
| Productivity lost due to severe disease  (I$ 2023) | Children (2-14) | 138 K (87.7 K - 195 K) | 193 K (123 K - 273 K) | 193 K (123 K - 273 K) | 249 K (158 K - 351 K) | 249 K (158 K - 351 K) |
|  | WCBA (15-49) | 1.02 M (669 K - 1.44 M) | 1.43 M (936 K - 2.01 M) | 1.43 M (936 K - 2.01 M) | 1.83 M (1.20 M - 2.58 M) | 1.83 M (1.20 M - 2.58 M) |
|  | Adolescents and adults (15-49) | 2.72 M (1.76 M - 3.81 M) | 3.80 M (2.46 M - 5.34 M) | 3.80 M (2.46 M - 5.34 M) | 4.89 M (3.17 M - 6.87 M) | 4.89 M (3.17 M - 6.87 M) |
|  | Older adults (50+) | 502 K (323 K - 729 K) | 703 K (452 K - 1.02 M) | 703 K (452 K - 1.02 M) | 904 K (581 K - 1.31 M) | 904 K (581 K - 1.31 M) |
|  | All (2+) | 3.36 M (2.18 M - 4.74 M) | 4.70 M (3.05 M - 6.63 M) | 4.70 M (3.05 M - 6.63 M) | 6.04 M (3.92 M - 8.52 M) | 6.04 M (3.92 M - 8.52 M) |
| Productivity lost due to hearing loss (I$ 2023) | Children (2-14) | 6.25 M (1.68 M – 17.0 M) | 6.35 M (1.72 M - 17.3 M) | 8.75 M (2.35 M - 23.9 M) | 8.85 M (2.39 M - 24.1 M) | 11.2 M (3.03 M - 30.7 M) |
|  | WCBA (15-49) | 17.7 M (5.43 M - 42.6 M) | 18.4 M (5.69 M - 44.2 M) | 24.8 M (7.60 M - 59.6 M) | 25.5 M (7.86 M - 61.2 M) | 31.9 M (9.77 M - 76.7 M) |
|  | Adolescents and adults (15-49) | 41.0 M (12.5 M - 98.5 M) | 42.8 M (13.2 M - 103 M) | 57.4 M (17.6 M - 138 M) | 59.2 M (18.2 M - 142 M) | 73.7 M (22.6 M - 177 M) |
|  | Older adults (50+) | 4.17 M (1.27 M - 9.89 M) | 4.47 M (1.38 M - 10.4 M) | 5.83 M (1.78 M - 13.8 M) | 6.13 M (1.89 M - 14.4 M) | 7.50 M (2.29 M - 17.8 M) |
|  | All (2+) | 51.4 M (15.5 M - 125 M) | 53.6 M (16.4 M - 130 M) | 71.9 M (21.7 M - 175 M) | 74.2 M (22.6 M - 180 M) | 92.5 M (27.9 M - 225 M) |
| Productivity lost due to death (I$ 2023) | Children (2-14) | 14.6 M (7.68 M - 24.6 M) | 20.5 M (10.8 M - 34.4 M) | 20.5 M (10.8 M - 34.4 M) | 26.3 M (13.8 M - 44.3 M) | 26.3 M (13.8 M - 44.3 M) |
|  | WCBA (15-49) | 39.4 M (23.1 M – 61.0 M) | 55.2 M (32.3 M - 85.5 M) | 55.2 M (32.3 M - 85.5 M) | 71.0 M (41.5 M - 110 M) | 71.0 M (41.5 M - 110 M) |
|  | Adolescents and adults (15-49) | 104 M (61.8 M - 162 M) | 146 M (86.6 M - 227 M) | 146 M (86.6 M - 227 M) | 188 M (111 M - 291 M) | 188 M (111 M - 291 M) |
|  | Older adults (50+) | 23.8 M (14.2 M - 35.5 M) | 33.3 M (19.9 M - 49.7 M) | 33.3 M (19.9 M - 49.7 M) | 42.9 M (25.6 M - 63.9 M) | 42.9 M (25.6 M - 63.9 M) |
|  | All (2+) | 143 M (87.0 M - 218 M) | 200 M (122 M - 305 M) | 200 M (122 M - 305 M) | 257 M (157 M - 392 M) | 257 M (157 M - 392 M) |

***4c. Partial rank correlation coefficients***

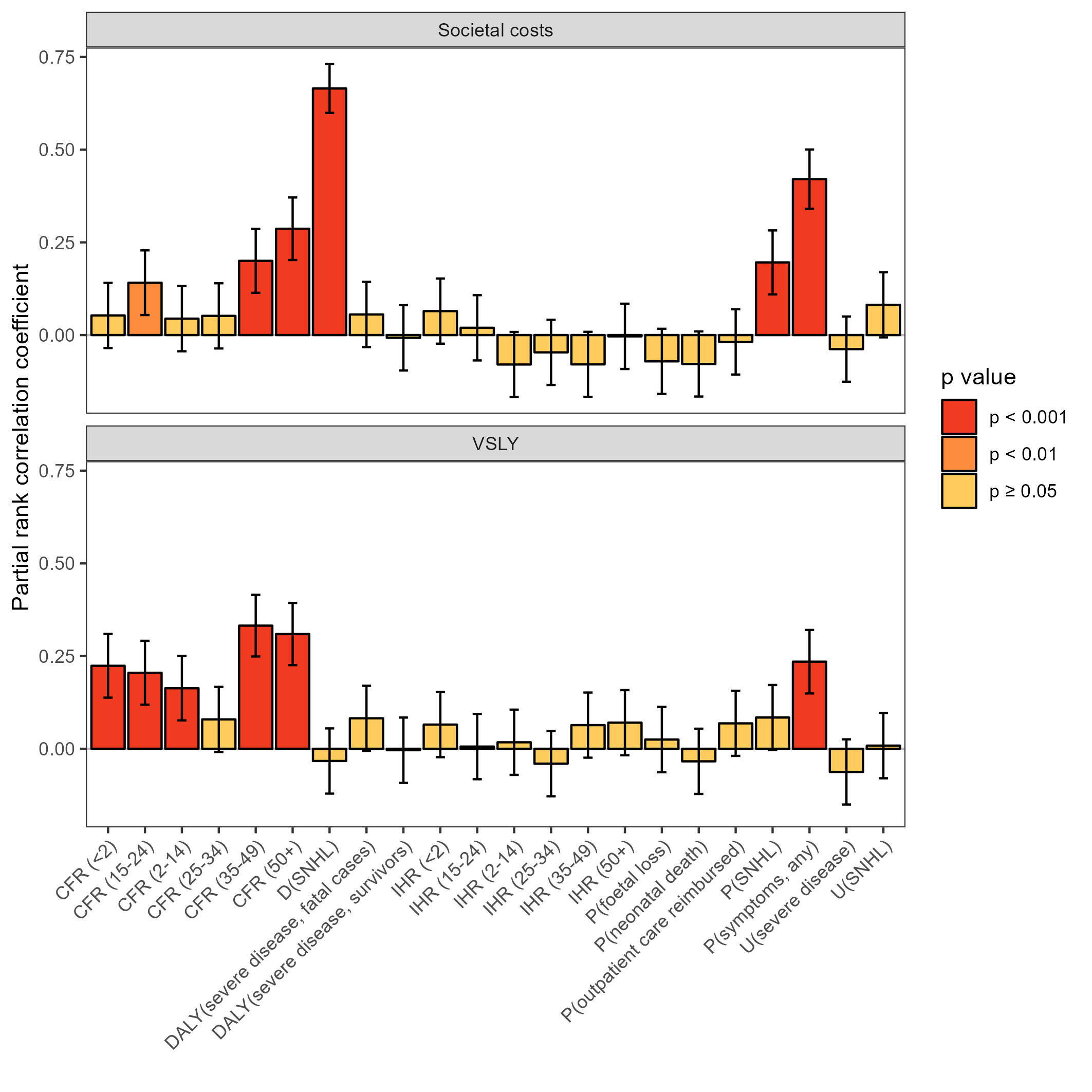

***Figure S30.*** *Partial rank correlation coefficients between model input parameters (x-axis) and two estimates of the cumulative total economic burden of Lassa fever in the absence of vaccination: societal costs (top panel) and VSLY (bottom panel). From left to right, input parameters varied along the x-axis include age-specific CFRs, the duration of SNHL, average DALYs per case for severe disease (the product of health state duration and corresponding disability), age-specific IHRs, probabilities of health state outcomes and health state disability weight estimates. The following input parameters were removed due to extreme multicollinearity and/or negligible variance: the durations of fever (mild/moderate symptoms), severe symptoms prior to hospitalisation and hospitalisation among survivors; disability weight for fever; per-case DALYs due to fever; and the probability of seeking any outpatient care. Although IHRs vary slightly between sexes, only male IHRs were included to reduce dimensionality. CFR = case-fatality risk, SNHL = sensorineural hearing loss, DALY = disability-adjusted life-year, IHR = infection-hospitalisation risk.*

6.	UN Department of Economic and Social Affairs Population Division. Data from: Population by single age – both sexes. 2024.

7.	UN Department of Economic and Social Affairs Population Division. Data from: Births, by age of mother. 2024.

8.	WorldPop. Age and sex structures / Constrained individual countries 2020. Accessed 20/02/2023, <https://hub.worldpop.org/geodata/listing?id=87>

9.	Basinski AJ, Fichet-Calvet E, Sjodin AR, et al. Bridging the gap: Using reservoir ecology and human serosurveys to estimate Lassa virus spillover in West Africa. *PLoS Comput Biol*. Mar 2021;17(3):e1008811. doi:10.1371/journal.pcbi.1008811

10.	Camacho A, Ndiaye A, Amevoin Y, Mousset M. *Enable interim analysis: preliminary results (data as of 24 October 2022)*. 2022. <https://cdn.who.int/media/docs/default-source/blue-print/day1_session1_7_anton-camacho_lassa-vaccine-meeting_nigeria.pdf?sfvrsn=60534f12_3>

11.	Gelman A, Carlin JB, Stern HS, Dunson DB, Vehtari A, Rubin DB. *Bayesian Data Analysis, Third Edition*. 2021.

43.	International Labour Organization. Data from: Labour force participation rate by sex and age (%). 2024.

44.	World Health Organization. *Global costs of unaddressed hearing loss and cost-effectiveness of interventions*. 2017. Accessed 17/10/2024. <https://iris.who.int/bitstream/handle/10665/254659/9789241512046-eng.pdf>

45.	McDaid D, Park AL, Chadha S. Estimating the global costs of hearing loss. *Int J Audiol*. Mar 2021;60(3):162-170. doi:10.1080/14992027.2021.1883197

46.	Stevens GA, Alkema L, Black RE, et al. Guidelines for Accurate and Transparent Health Estimates Reporting: the GATHER statement. *Lancet*. Dec 10 2016;388(10062):e19-e23. doi:10.1016/S0140-6736(16)30388-9
